# Behaviour change interventions improve maternal and child nutrition in sub-Saharan Africa: a systematic review

**DOI:** 10.1101/2022.03.30.22273189

**Authors:** Daniella Watson, Patience Mushamiri, Paula Beeri, Toussaint Rouamba, Sarah Jenner, Sarah H Kehoe, Kate A Ward, Mary Barker, Wendy Lawrence, the INPreP Study Group

## Abstract

**Background:** Evidence that nutrition-specific and nutrition-sensitive interventions can improve maternal and child nutrition status in sub-Saharan Africa is inconclusive. Using behaviour change theory and techniques in intervention design may increase effectiveness and make outcomes more predictable. This systematic review aimed to determine whether interventions that included behaviour change functions were effective.

**Methods:** Six databases were searched systematically, using MeSH and free-text terms, for articles describing nutrition-specific and nutrition-sensitive behaviour change interventions published in English until January 2022. Titles, abstracts and full-text papers were double-screened. Data extraction and quality assessments followed Centre for Reviews and Dissemination guidelines. Behaviour change functions of interventions were mapped onto the COM-B model and Behaviour Change Wheel. PROSPERO registered (135054).

**Findings:** The search yielded 1149 articles: 71 articles met inclusion criteria, ranging from low (n=30) to high (n=11) risk of bias. Many that applied behaviour change theory, communication or counselling resulted in significant improvements in infant stunting and wasting, household dietary intake and maternal psychosocial measures. Interventions with >2 behaviour change functions (including persuasion, incentivisation, environmental restructuring) were the most effective.

**Interpretation:** We recommend incorporating behaviour change functions in nutrition interventions to improve maternal and child outcomes, specifically drawing on the Behaviour Change Wheel, COM-B model. To enhance the designs of these interventions, and ultimately improve the nutritional and psychosocial outcomes for mothers and infants in sub-Saharan Africa, collaborations are recommended between behaviour change and nutrition experts, intervention designers, policy makers and commissioners to fund and roll-out multicomponent behaviour change interventions.

## Background

The triple burden of malnutrition^(1)^, coexistence of under- and over-nutrition^(2)^ and poor micronutrient status impacts maternal and child health^(3–6)^. In sub-Saharan Africa this occurs at the household level, where family members can be underweight (BMI<18.5kg/m^2^)^(7)^ or overweight (BMI>25kg/m^2^)^(7)^, and at the individual level, where an infant can be stunted (height for age < −2 SD from the median)^(7)^, wasted (weight for age <-2SD from the median)^(7)^ or overweight^(8)^. One meta-analysis found that rural residency, low educational status of partners, multiple pregnancies and poor nutritional indicators were determinants of malnutrition in pregnancy^(9)^. A recent Lancet series^(3–6, 10–17)^ advocated for person-centred health systems and proposed a lifecourse double-duty approach to optimise dietary quality to address both under- and over-nutrition^(11–14)^. They advise using both “nutrition-specific” and “nutrition-sensitive” interventions^(3–6)^. Large-scale nutrition-sensitive interventions (agriculture, clean water, sanitation, education and employment) may enhance nutrition-specific interventions (breastfeeding promotion, fortification of foods, and micronutrient supplementation) by addressing underlying determinants of nutrition^(5, 18)^. Approaches to improve infant nutrition status include 1) promotion of individual behaviour change to address diet quality, 2) food fortification, 3) nutritional supplementation^(18–20)^.

Behavioural functions of interventions are increasingly recognised as crucial, yet expertise in global health behaviour change is lacking^(21)^. Shelton (2013) described behaviour change as the missing block in global health systems, pointing out that the top 20 health risks (e.g. obesity, childhood underweight, vitamin deficiencies) in sub-Saharan Africa are influenced by behaviour (e.g. care seeking, adherence, pro-social behaviours)^(21)^. Public health and health promotion interventions based on social and behavioural science theories are more effective than interventions without such a theoretical base^(22)^. Further, a lack of theory-informed evaluations limits what can be learnt about how interventions work in different contexts, how health impacts are observed, and effects on primary and secondary outcomes^(23–25)^.

Research suggests that intervention strategies must combine behaviour change with access to nutritious food^(26)^, acknowledging that people eat ‘food’ rather than ‘nutrients’^(27)^. Behavioural scientists argue that behaviour change interventions should engage with the target population, understand their motivation to change, and adapt to the contexts that facilitate change including environment and social networks^(26)^. The Behaviour Change Wheel (Figure 1) encapsulates features applicable to intervention design and implementation^(28)^. It outlines nine behavioural intervention functions and seven policy categories. It also includes three underlying human factors – **C**apability (physical and psychological), **O**pportunity (physical and social), and **M**otivation (automatic and reflective) that influence **B**ehaviour (COM-B model) (Figure 2). Both models can be applied to inform the design of interventions to improve maternal and child nutrition in the context into which they are being implemented.

**Figure 1.**
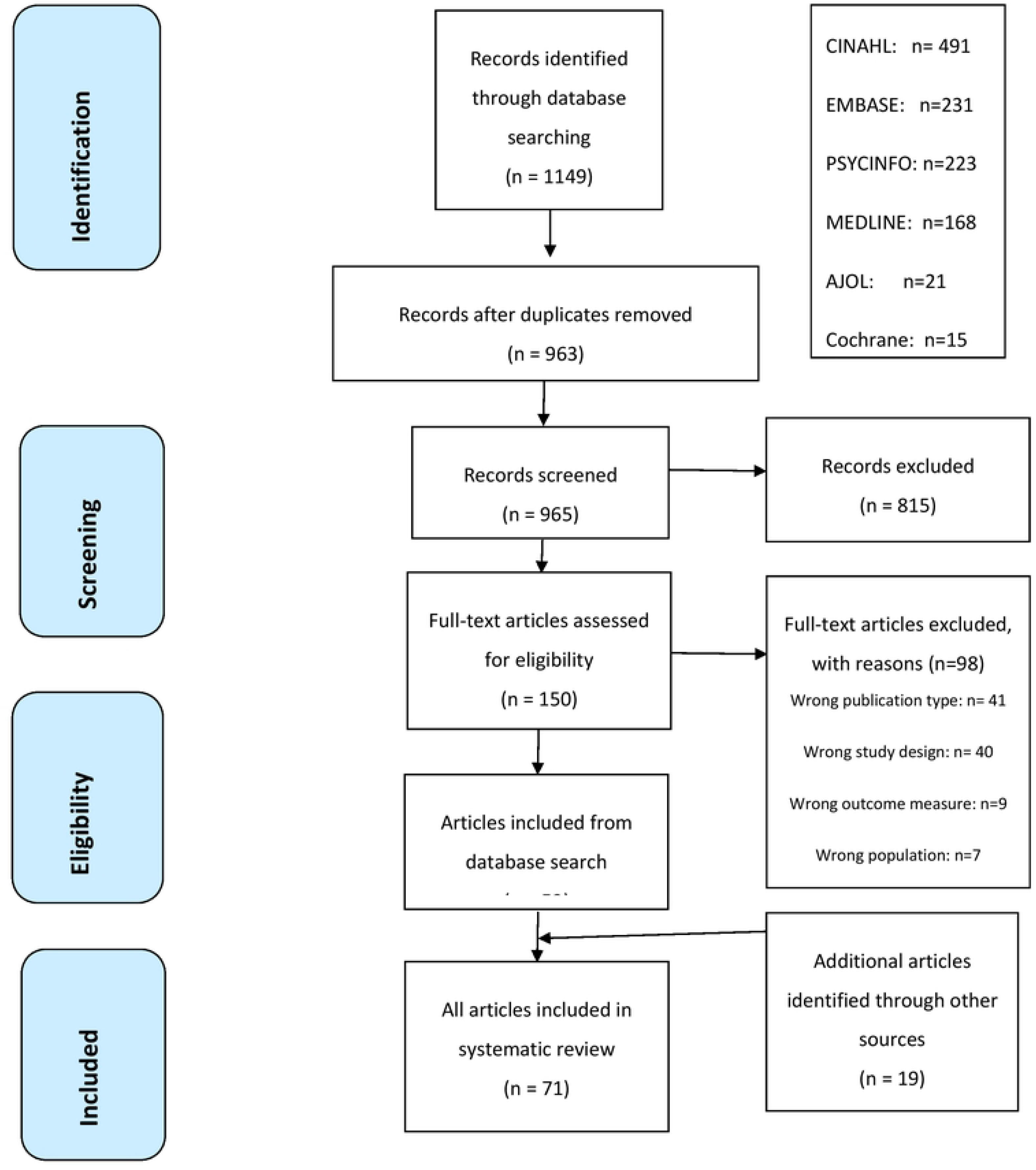
Behaviour Change Wheel (Michie et al, 2011)^(28)^

**Figure 2.**
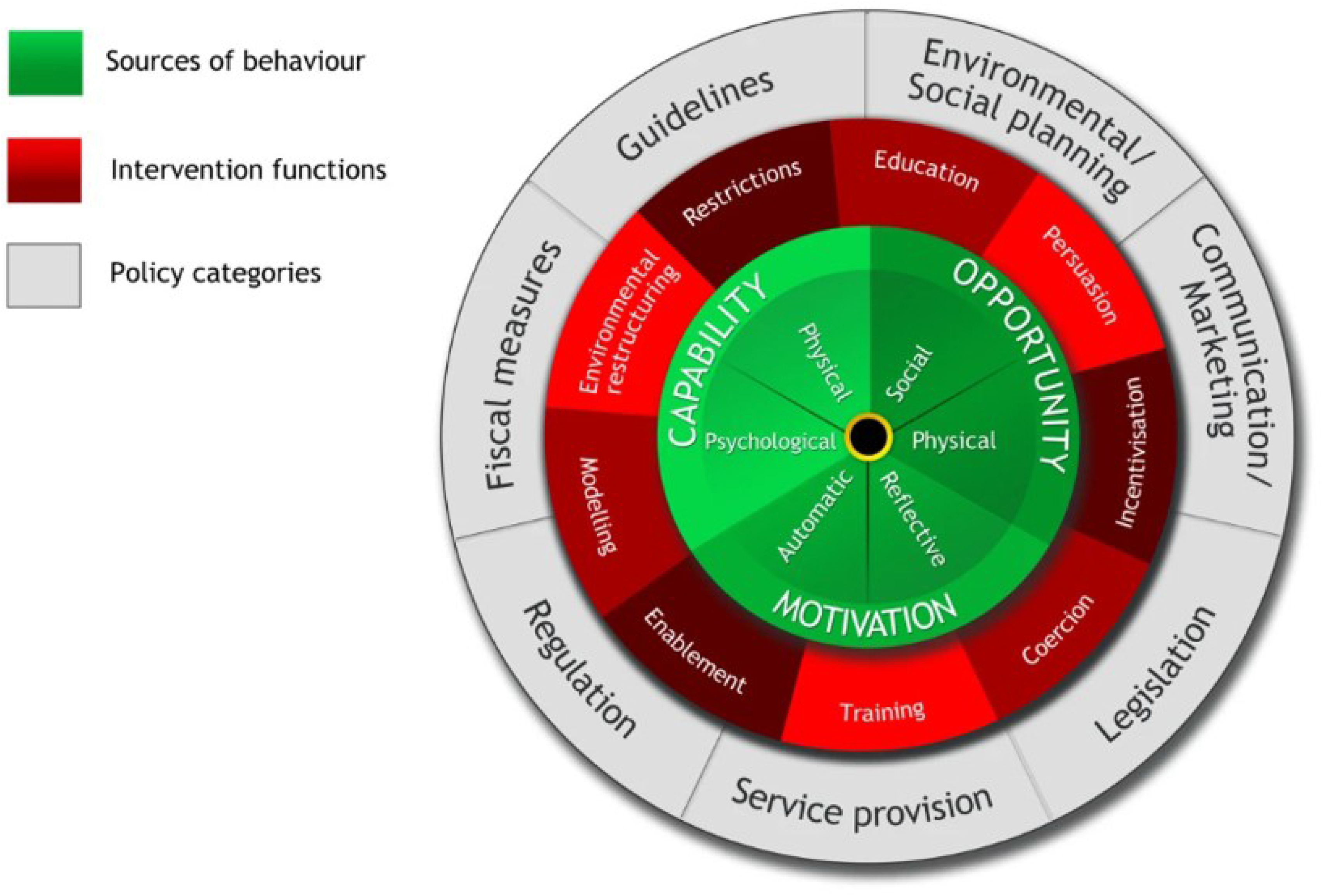
The COM-B model (Capability, Motivation, Opportunity – Behaviour) (Michie et al, 2011)^(28)^

A systematic review was undertaken to collate evidence to answer the following questions: 1) are nutrition-specific and nutrition-sensitive interventions effective in improving maternal and child nutrition in sub-Saharan Africa if they include behaviour change intervention functions? 2) which functions of behaviour change interventions are associated with improvements in maternal and child nutrition outcomes?

## Materials and Method

### Search strategy and study selection

This systematic review follows the Centre for Reviews and Dissemination^(29)^ and PRISMA guidelines^(30, 31)^. It is PROSPERO registered (135054). Articles were systematically searched for on major medical and social science databases including Cochrane library, EMBASE, MEDLINE, PSYCHINFO, CINAHL and African Journal OnLine (AJOL) published up to 24^th^ January 2022. The search strategy was based on the most prevalent behaviour change concepts and theories identified by Davis et al (2015)^(32)^. Nutrition-specific and nutrition-sensitive interventions, and maternal and child nutrition terms were derived from the 2013 Lancet series on Maternal and Child Nutrition^(4, 5)^ and in consultation with experts. The terms were entered into the databases using a combination of Medical Subject Headings (MeSH) and free text to include behaviour change interventions for maternal and child nutrition in sub-Saharan Africa (Appendix A). The terms were based on English and American spelling and were not translated into other languages. French articles were identified from the database search and were reviewed by a French speaking screener working on this review. As French articles were already included in our search, it was decided not to also translate the search terms into French. This may be a limitation as some French and other language articles might not have been identified. Further studies were sought by hand-searching bibliographies of included studies, tracking citations on Google Scholar, and asking experts on the topic. Breastfeeding interventions were excluded as there have been previous systematic reviews on this topic, which incidentally conclude that evidence for including behaviour change in breastfeeding and complimentary feeding interventions is lacking^(33, 34)^.

Publications were stored in Endnote X9.2, duplications were removed and papers were screened on Rayyan qcri online website^(35)^. Paper titles, abstracts and full text were double-screened. Inclusion criteria can be found in Table 1 and Appendix B. Full texts were also screened against the Behaviour Change Wheel intervention functions^(28)^ (Appendix C) to only include interventions that had a behaviour change component. Discrepancies were resolved by team review, which was deemed the final consensus.

**Table 1.** Systematic review inclusion and exclusion criteria

|  | **Inclusion criteria** | **Exclusion criteria** |
| --- | --- | --- |
| **Publication type** | Journal articles | Protocols<br>Commentary articles<br>Conference papers<br>Abstracts<br>Theses |
| **Study design** | Interventional studies with or without control group<br>Experimental or quasi-experimental design<br>Primary data collection<br>Qualitative evaluations of an intervention | Observational studies<br>Qualitative methodologies to explore perceptions of nutrition<br>Demographic surveillance to investigate nutrition status |
| **Population** | At least one of the following populations living in sub-Saharan Africa:<br>- women of child-bearing age (preconception), including adolescents<br>- pregnant women<br>- fathers<br>- infants, children and adolescents | Populations living outside sub-Saharan Africa<br>Expats living in sub-Saharan Africa |
| **Interventions** | Nutrition related behaviour change interventions | Solely breastfeeding or complimentary feeding interventions<br>Non-behaviour change interventions<br>Non-nutrition interventions solely focused on<br>- HIV/AIDS,<br>- malaria<br>- other infectious diseases |
| **Outcomes** | At least one of the following outcomes for:<br>- Body composition, nutrient intake, dietary patterns<br>- Health outcomes including physical and psychological<br>- Behavioural or psychosocial outcomes related to nutrition<br>- Short- and long-term outcomes |  |

### Study quality assessment and data extraction

The interventions identified were mixed methods and heterogeneous and therefore a meta-analysis was not deemed appropriate. A quality assessment tool was adapted from a systematic review on digital interventions using mixed methods and tailored for use in this review (Appendix D & Appendix E)^(36)^, based on the Centre for Reviews and Dissemination quality assessment criteria^(29)^. Papers were assessed for risk of bias by two reviewers independently ^(Table 2)^. Scores between −4 and 4 indicated a medium risk of bias. Scores below - 4 and above 4 indicated a high and low risk of bias respectively.

**Table 2.** Risk of Bias score table

| # | Author, year | Study design | Randomisation method | Blinding | Difference between groups | Selection | Loss to follow-up | Dietary assessment | Behaviour change component | Performance bias | Intention to Treat | Analysis | Confounding | Total | Risk of Bias |
| --- | --- | --- | --- | --- | --- | --- | --- | --- | --- | --- | --- | --- | --- | --- | --- |
| 1 | Abiyu, 2020 | +1 | +1 | 0 | +1 | +1 | +1 | 0 | 0 | 0 | +1 | +1 | +1 | **8** | Low |
| 2 | Antwi, 2020 | +1 | +1 | -1 | 0 | 0 | +1 | 0 | +1 | 0 | -1 | +1 | -1 | **2** | Medium |
| 3 | Aubel, 2004 | 0 | -1 | -1 | -1 | 0 | -1 | 0 | 0 | 0 | N/A | 0 | -1 | **0** | High |
| 4 | Becquey, 2019 | +1 | +1 | -1 | -1 | +1 | 0 | 0 | 0 | 0 | +1 | +1 | +1 | **4** | Medium |
| 5 | Briaux, 2020 | +1 | +1 | -1 | +1 | +1 | 0 | 0 | 0 | -1 | +1 | +1 | 0 | **4** | Medium |
| 6 | Byrd, 2019 | +1 | +1 | 0 | 0 | +1 | 0 | 0 | +1 | +1 | N/A | +1 | +1 | **7** | Low |
| 7 | DeLorme, 2018 | -1 | -1 | N/A | -1 | -1 | +1 | 0 | 0 | 0 | N/A | +1 | -1 | **-3** | Medium |
| 8 | Demilew, 2020 | +1 | +1 | 0 | +1 | +1 | +1 | +1 | +1 | 0 | 0 | +1 | +1 | **9** | Low |
| 9 | Diddana, 2018 | +1 | +1 | 0 | +1 | +1 | 0 | -1 | +1 | 0 | N/A | +1 | +1 | **6** | Low |
| 10 | Dozio, 2016 | -1 | -1 | -1 | -1 | 0 | -1 | 0 | +1 | 0 | N/A | -1 | -1 | **-6** | High |
| 11 | Ezezika, 2018 | -1 | -1 | N/A | 0 | -1 | +1 | 0 | -1 | 0 | N/A | -1 | N/A | **-4** | High |
| 12 | Felegush, 2018 | +1 | +1 | -1 | +1 | +1 | +1 | +1 | +1 | 0 | N/A | +1 | +1 | **8** | Low |
| 13 | Fernandes, 2016 | -1 | +1 | -1 | 0 | +1 | -1 | -1 | +1 | -1 | N/A | -1 | -1 | **-4** | High |
| 14 | Flax, 2021 | +1 | +1 | -1 | 0 | +1 | +1 | 0 | 0 | 0 | -1 | +1 | +1 | **4** | Medium |
| 15 | Galasso, 2009 | 0 | +1 | -1 | 0 | +1 | -1 | +1 | 0 | 0 | +1 | +1 | +1 | **4** | Medium |
| 16 | Galasso, 2019 | +1 | +1 | -1 | +1 | +1 | +1 | +1 | 0 | +1 | +1 | +1 | +1 | **9** | Low |
| 17 | Gelli, 2018 | +1 | +1 | +1 | +1 | +1 | +1 | +1 | +1 | +1 | +1 | +1 | -1 | **10** | Low |
| 18 | Gelli, 2020 | +1 | 0 | +1 | +1 | +1 | +1 | +1 | +1 | +1 | +1 | +1 | -1 | **10** | Low |
| 19 | Han, 2021 | +1 | +1 | -1 | +1 | +1 | 0 | +1 | +1 | 0 | +1 | 0 | +1 | **7** | Low |
| 20 | Hurley | 0 | -1 | -1 | 0 | 0 | +1 | +1 | 0 | 0 | N/A | +1 | +1 | **2** | Medium |
| 21 | Huybregts, 2019 | +1 | +1 | 0 | 0 | +1 | 0 | -1 | +1 | 0 | +1 | +1 | +1 | **6** | Low |
| 22 | Jacobs, 2013 | +1 | +1 | -1 | 0 | +1 | -1 | -1 | +1 | 0 | N/A | +1 | -1 | **1** | Medium |
| 23 | Jemmott, 2011; | +1 | +1 | +1 | +1 | +1 | +1 | 0 | +1 | +1 | +1 | +1 | +1 | **11** | Low |
| 24 | Jemmott, 2019 | +1 | +1 | +1 | +1 | 0 | +1 | 0 | +1 | +1 | +1 | +1 | +1 | **10** | Low |
| 25 | Kang, 2016 | +1 | +1 | -1 | -1 | -1 | +1 | -1 | -1 | +1 | N/A | +1 | -1 | **-1** | Medium |
| 26 | Kang, 2017 | +1 | +1 | -1 | -1 | -1 | +1 | -1 | -1 | +1 | N/A | +1 | -1 | **-1** | Medium |
| 27 | Kang, 2017 | 0 | -1 | -1 | -1 | 0 | -1 | +1 | -1 | +1 | 0 | 0 | -1 | **-4** | Medium |
| 28 | Katenga-Kaunda 2021 | +1 | +1 | -1 | +1 | +1 | 0 | 0 | 0 | 0 | -1 | +1 | +1 | **4** | Medium |
| 29 | Kim, 2019, | +1 | +1 | 0 | 0 | +1 | +1 | 0 | +1 | 0 | +1 | +1 | +1 | **8** | Low |
| 30 | Kim, 2015 | -1 | -1 | -1 | -1 | -1 | 1 | -1 | -1 | 0 | N/A | 0 | -1 | **-7** | High |
| 31 | Kim, 2016 | 0 | 1 | -1 | 1 | 1 | -1 | 1 | 0 | 0 | -1 | 1 | 1 | **3** | Medium |
| 32 | Kumar, 2018 | +1 | +1 | -1 | +1 | 0 | -1 | 0 | +1 | 0 | +1 | +1 | +1 | **5** | Low |
| 33 | Rosenberg, 2018 | +1 | +1 | -1 | +1 | 0 | -1 | 0 | +1 | 0 | +1 | +1 | +1 | **5** | Low |
| 34 | Lagerkvist, 2018 | -1 | +1 | -1 | +1 | +1 | -1 | -1 | +1 | 0 | N/A | +1 | +1 | **2** | Medium |
| 35 | Leroy, 2016 | +1 | +1 | -1 | +1 | 0 | 0 | 0 | 0 | +1 | +1 | +1 | +1 | **6** | Low |
| 36 | Leroy, 2018 | +1 | +1 | -1 | +1 | 0 | 0 | 0 | 0 | +1 | +1 | +1 | +1 | **6** | Low |
| 37 | Leroy, 2020 | +1 | +1 | -1 | +1 | 0 | 0 | 0 | 0 | +1 | +1 | +1 | +1 | **6** | Low |
| 38 | Olney, 2019 | +1 | +1 | -1 | +1 | 0 | 0 | 0 | 0 | +1 | +1 | +1 | +1 | **6** | Low |
| 39 | Heckert, 2020 | 0 | -1 | +1 | +1 | 0 | N/A | N/A | 0 | +1 | N/A | +1 | +1 | **4** | Medium |
| 40 | Lion, 2018 | 0 | +1 | -1 | 0 | +1 | 0 | -1 | +1 | +1 | N/A | +1 | +1 | **4** | Medium |
| 41 | McKune, 2021 | +1 | +1 | +1 | +1 | +1 | -1 | +1 | 0 | +1 | -1 | +1 | +1 | **7** | Medium |
| 42 | Mlinda, 2018 | +1 | +1 | -1 | +1 | +1 | +1 | 0 | 0 | 0 | +1 | +1 | +1 | **7** | Low |
| 43 | Morris, 2012 | -1 | -1 | 0 | +1 | -1 | +1 | 0 | +1 | 0 | +1 | +1 | +1 | **3** | Medium |
| 44 | Muehlhoff, 2016 | 0 | -1 | -1 | -1 | 0 | +1 | -1 | -1 | -1 | N/A | -1 | -1 | **-7** | High |
| 45 | Mushaphi, 2015 | 0 | +1 | -1 | +1 | -1 | -1 | +1 | -1 | 0 | N/A | 0 | -1 | **-2** | Medium |
| 46 | Mushaphi, 2017 | 0 | +1 | -1 | +1 | -1 | -1 | +1 | -1 | 0 | N/A | 0 | -1 | **-2** | Medium |
| 47 | Mutiso, 2018 | -1 | +1 | -1 | -1 | +1 | -1 | 0 | +1 | 0 | N/A | +1 | +1 | **1** | Medium |
| 48 | Ogunsile & Ogunsile, 2016 | 0 | +1 | -1 | 0 | +1 | +1 | 0 | +1 | +1 | n/a | +1 | -1 | **4** | Medium |
| 49 | Olney, 2015 | +1 | +1 | 0 | +1 | +1 | 0 | +1 | +1 | +1 | N/A | +1 | +1 | **9** | Low |
| 50 | Olney, 2016 | +1 | +1 | 0 | +1 | +1 | 0 | +1 | +1 | +1 | N/A | +1 | +1 | **9** | Low |
| 51 | Nielsen, 2019 | 0 | +1 | 0 | +1 | +1 | 0 | 0 | 0 | +1 | N/A | +1 | +1 | **6** | Low |
| 52 | Passarelli, 2020 | +1 | +1 | -1 | 0 | +1 | +1 | 0 | +1 | +1 | -1 | +1 | +1 | **6** | Low |
| 53 | Ruel-Bergeron, 2019a | -1 | -1 | -1 | -1 | +1 | -1 | +1 | 0 | -1 | -1 | -1 | -1 | **-6** | High |
| 54 | Ruel-Bergeron, 2019b | 0 | -1 | -1 | -1 | +1 | -1 | N/A | 0 | 0 | N/A | 0 | N/A | **-3** | Medium |
| 55 | Christian, 2020 | 0 | -1 | -1 | -1 | +1 | -1 | 0 | 0 | -1 | -1 | -1 | -1 | **-7** | High |
| 56 | Saaka, 2011 | 0 | -1 | -1 | -1 | 0 | -1 | -1 | -1 | 0 | N/A | -1 | -1 | **-7** | High |
| 57 | Saaka, 2021 | 0 | +1 | -1 | 0 | +1 | -1 | +1 | 0 | 0 | N/A | +1 | +1 | **3** | Medium |
| 58 | Skar, 2019 | 0 | 0 | -1 | 0 | 0 | -1 | +1 | -1 | 0 | N/A | +1 | -1 | **-2** | Medium |
| 59 | Santoso, 2021 | +1 | +1 | 0 | +1 | -1 | +1 | +1 | -1 | -1 | +1 | +1 | +1 | **5** | Low |
| 60 | Shapu, 2021 | +1 | +1 | 0 | +1 | +1 | +1 | 0 | +1 | +1 | +1 | 0 | +1 | **9** | Low |
| 61 | Steyn, 2015 | +1 | +1 | 0 | 0 | +1 | +1 | 0 | -1 | 0 | N/A | 0 | -1 | **2** | Medium |
| 62 | De Villiers, 2016 | +1 | +1 | 0 | 0 | +1 | +1 | 0 | -1 | 0 | N/A | 0 | -1 | **2** | Medium |
| 63 | Tamiru, 2017 | 0 | +1 | -1 | +1 | 0 | -1 | +1 | +1 | +1 | n/a | 0 | +1 | **4** | Medium |
| 64 | Tariku, 2015; | +1 | +1 | -1 | +1 | +1 | +1 | 0 | +1 | 0 | n/a | +1 | 0 | **6** | Low |
| 65 | Mulualem, 2016 | +1 | +1 | -1 | +1 | +1 | +1 | 0 | +1 | 0 | n/a | +1 | 0 | **6** | Low |
| 66 | Thuita, 2015; | 0 | -1 | -1 | -1 | 0 | -1 | -1 | -1 | -1 | N/A | -1 | -1 | **-9** | High |
| 67 | Mukuria, 2016 | 0 | -1 | -1 | -1 | 0 | -1 | -1 | -1 | -1 | N/A | -1 | -1 | **-9** | High |
| 68 | Tomlinson, 2020 | 0 | +1 | -1 | -1 | +1 | 0 | +1 | +1 | 0 | -1 | +1 | -1 | **1** | Medium |
| 69 | Walton, 2017 | +1 | +1 | +1 | +1 | +1 | +1 | 0 | +1 | -1 | N/A | +1 | +1 | **8** | Low |
| 70 | Workicho,<br>2021 | 0 | -1 | -1 | +1 | -1 | +1 | 0 | 0 | 0 | N/A | 0 | +1 | -1 | Medium |
| 71 | Yetnayet,<br>2017 | 0 | +1 | -1 | +1 | 0 | -1 | 0 | +1 | 0 | N/A | +1 | +1 | 3 | Medium |

### Data analysis

Study designs were coded using an established method^(37)^ based on the functions from the Behaviour Change Wheel and coded to the COM-B model^(28)^. It is a well-evidenced behaviour change framework and this type of analysis has been carried out in other research projects^(28)^. Findings are described using a narrative synthesis approach^(38)^.

## Results

1149 papers were identified leaving 963 studies after duplicates were removed to be screened by titles and abstracts. From this screening, 150 papers were eligible to progress to full text screening. Nineteen further articles, six new interventions studies and 13 papers related to studies already included in this review, were found during the bibliography search and expert consultation. A total of 71 articles were included (Figure 3).

**Figure 3.**
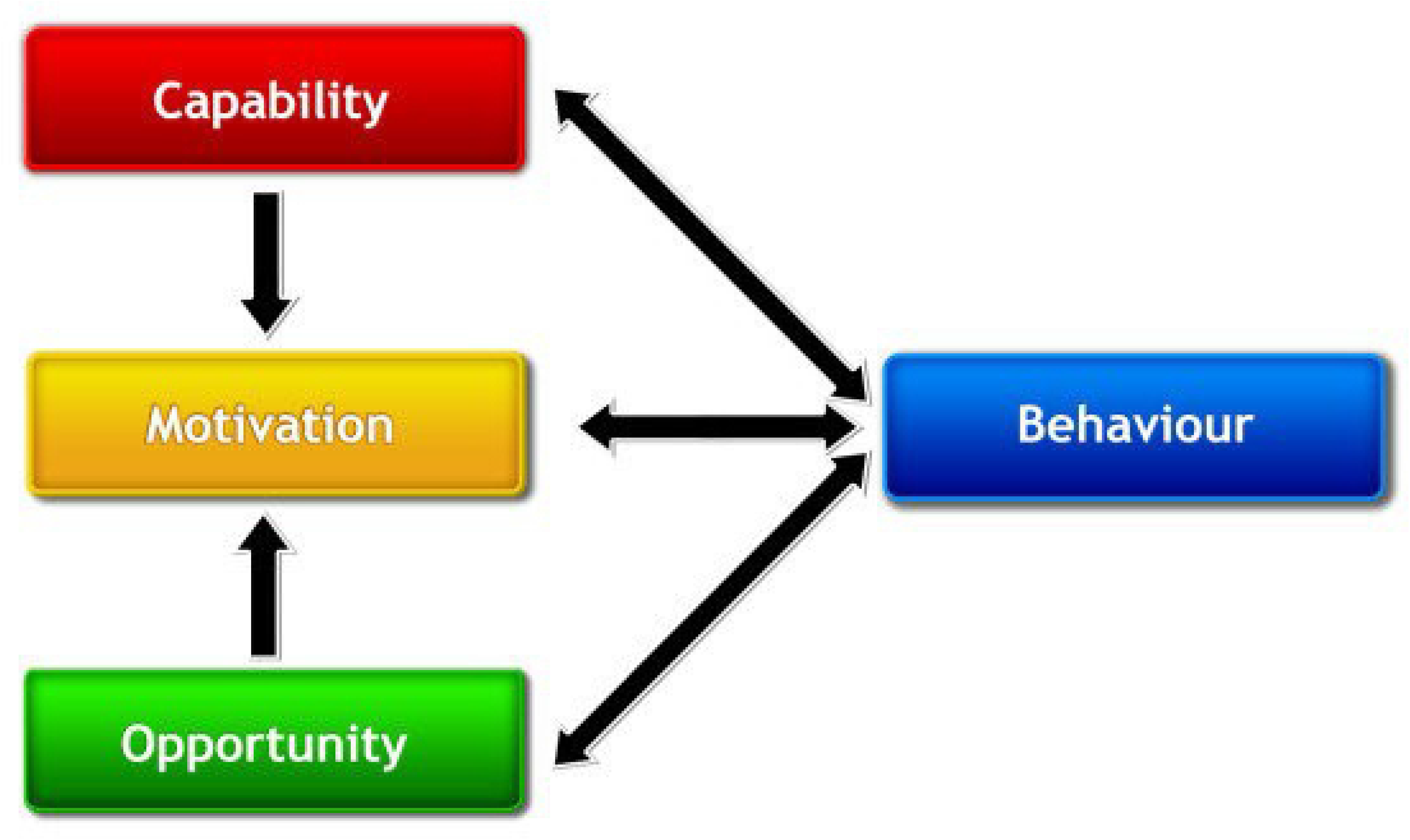
Flowchart of study selection PRISMA diagram

The 71 articles were published between 2004-2022 and included cluster randomised control trials (RCT)(n=38)^(20, 39–75)^, study evaluations(n=13)^(52, 56, 68, 76–86)^, quantitative studies(n=11)^(87–100)^, mixed methods studies(n=5)^(88, 89, 101–103)^, qualitative studies(n=3)^(104–106)^, and national case studies(n=2)^(107, 108)^. Behaviour change interventions were identified from 18 sub-Saharan African countries: Ethiopia(n=18)^(42, 58, 59, 62, 64, 67, 68, 70, 79–81, 92–95, 100)^, South Africa(n=8)^(40, 41, 45, 46, 66, 87–89)^, Malawi(n=8)^(43, 44, 71, 77, 78, 82, 85, 106)^, Kenya(n=5)^(39, 96, 101, 102, 104)^, Burundi(n=5)^(48–52)^, Nigeria(n=5)^(73, 90, 97, 99, 105)^, Burkina Faso(n=5)^(48, 54, 56, 74, 109)^, Ghana(n=4)^(84, 91, 108)^, Madagascar(n=2)^(20, 75)^, Zambia(n=2)^(83, 110)^,Tanzania(n=2)^(47, 72)^ and one each from, Central African Republic^(107)^, Mali^(63)^, Mozambique^(65)^, Rwanda^(69)^, Senegal ^(103)^, Togo^(61)^, and Uganda^(53)^. A total of 30 studies were graded as low risk of bias, 30 as medium, and 11 as high ^(Table 2)^. Studies were not removed if they were deemed low quality as findings can still be informative, even if there might be some bias in the intervention design. However, all findings are presented alongside their risk of bias to highlight which studies’ findings may be less credible. The studies deemed as low quality are not given as much weight in this review due to the possible bias and accuracy of results and interpretation.

Descriptions of intervention design, outcomes and implications are presented in Table 3. The results are summarised below by the impact the interventions had on 1) anthropometric markers, 2) dietary outcomes, 3) psychosocial outcomes and 4) behavioural intervention components.

**Table 3.**
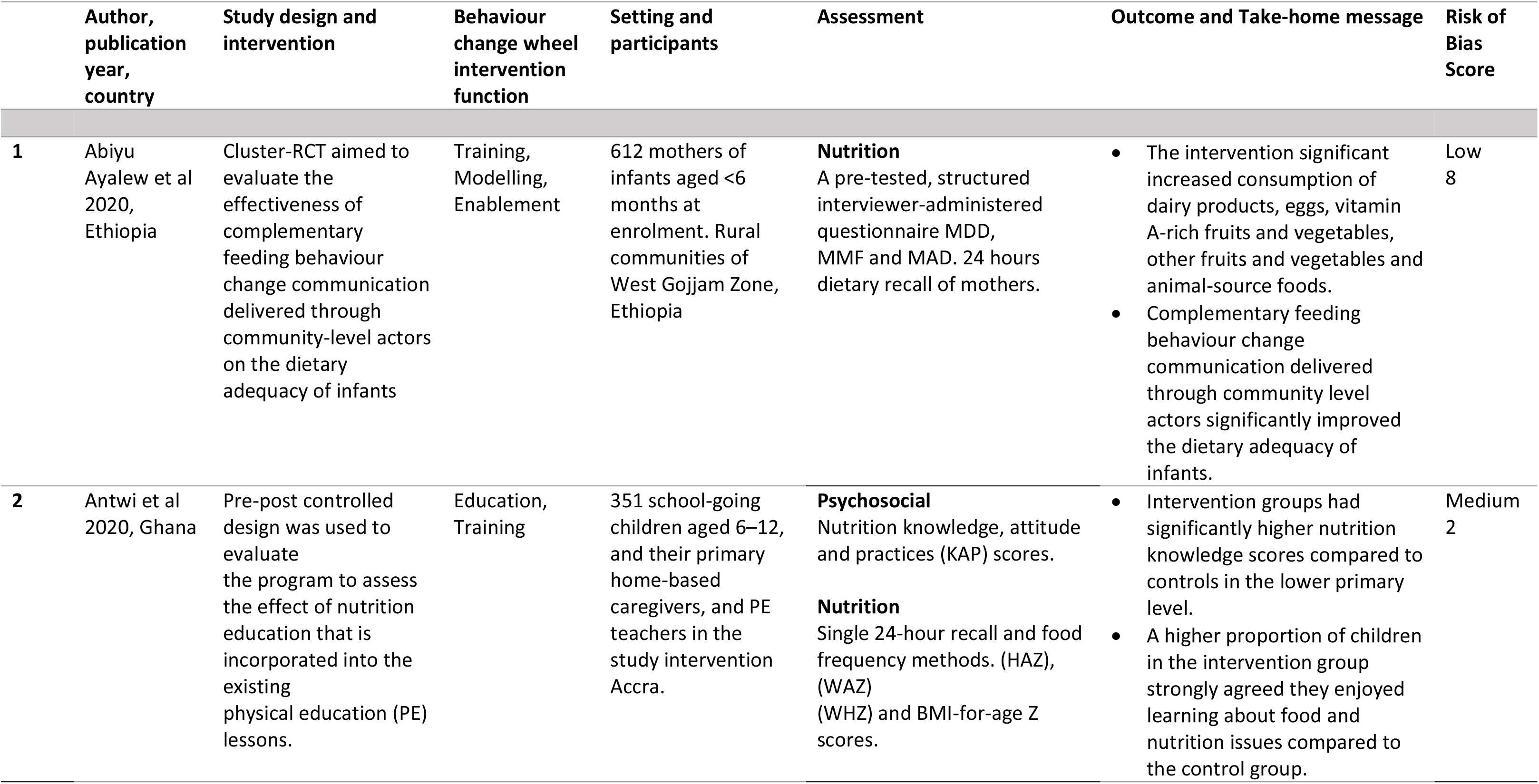
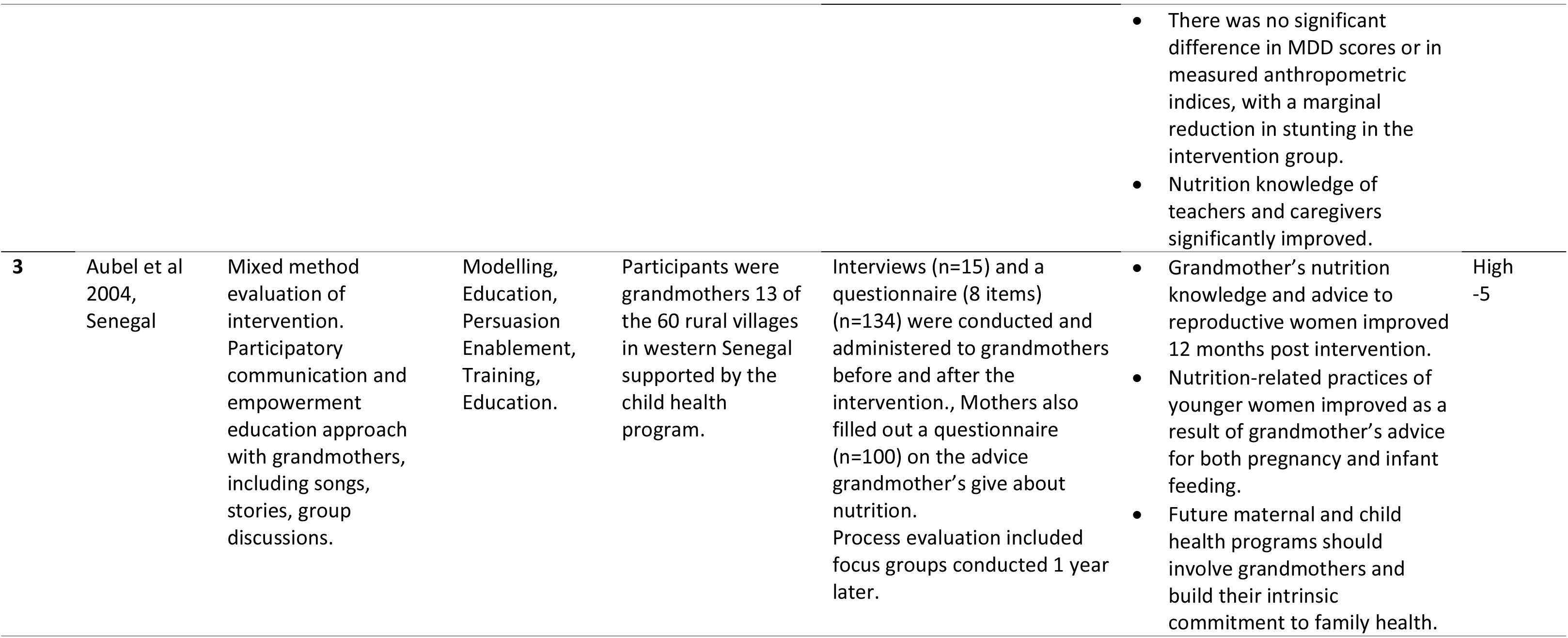
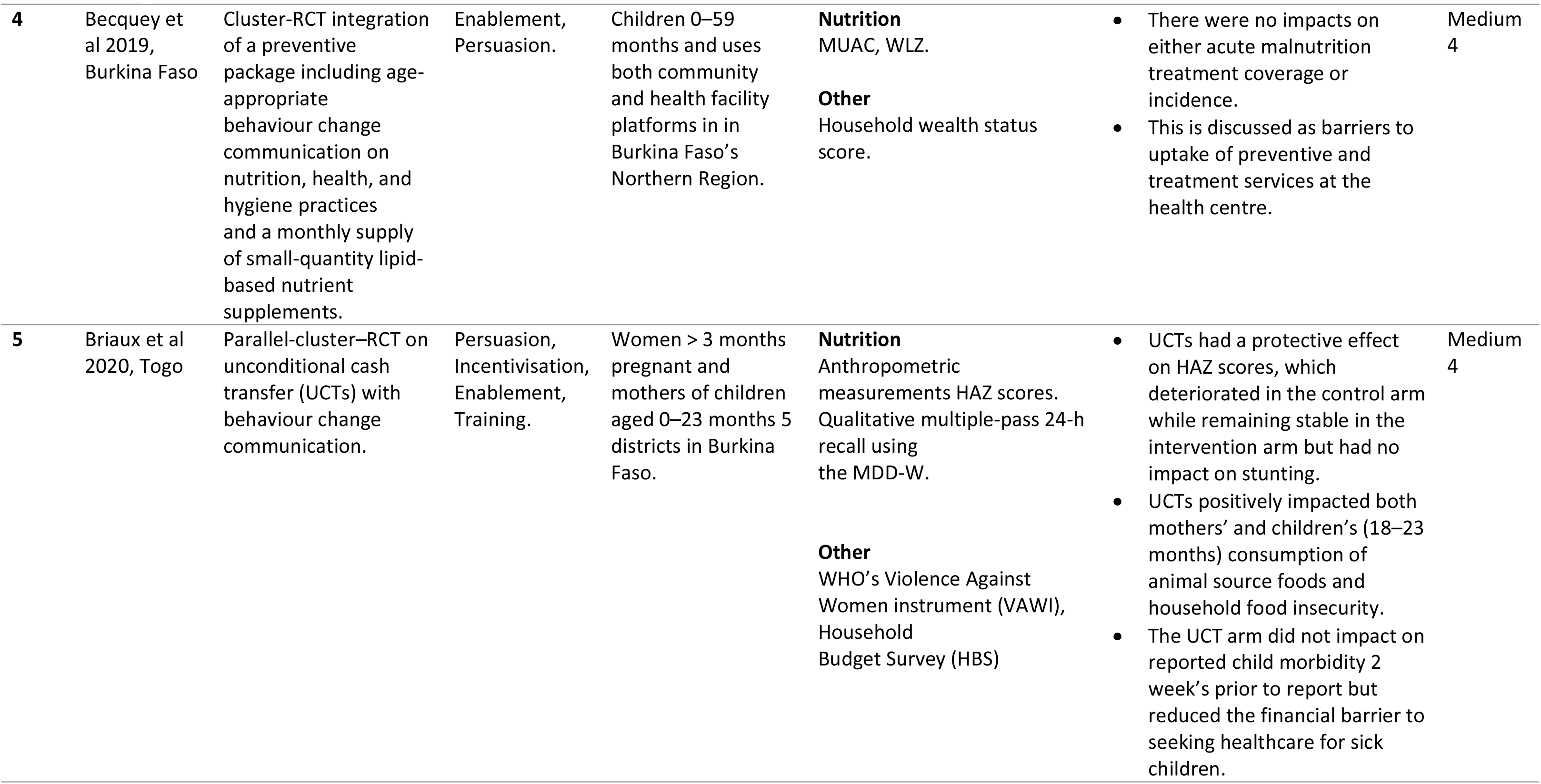
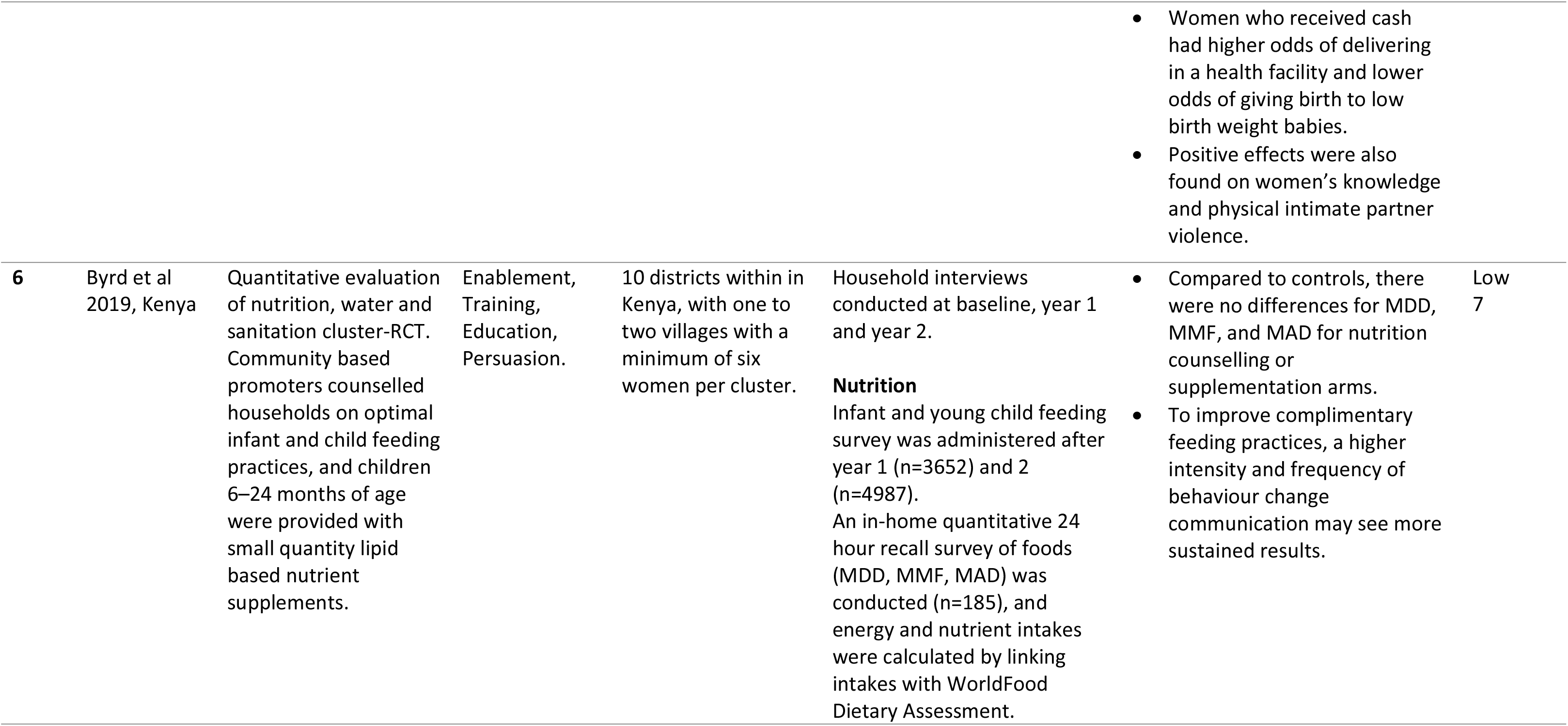
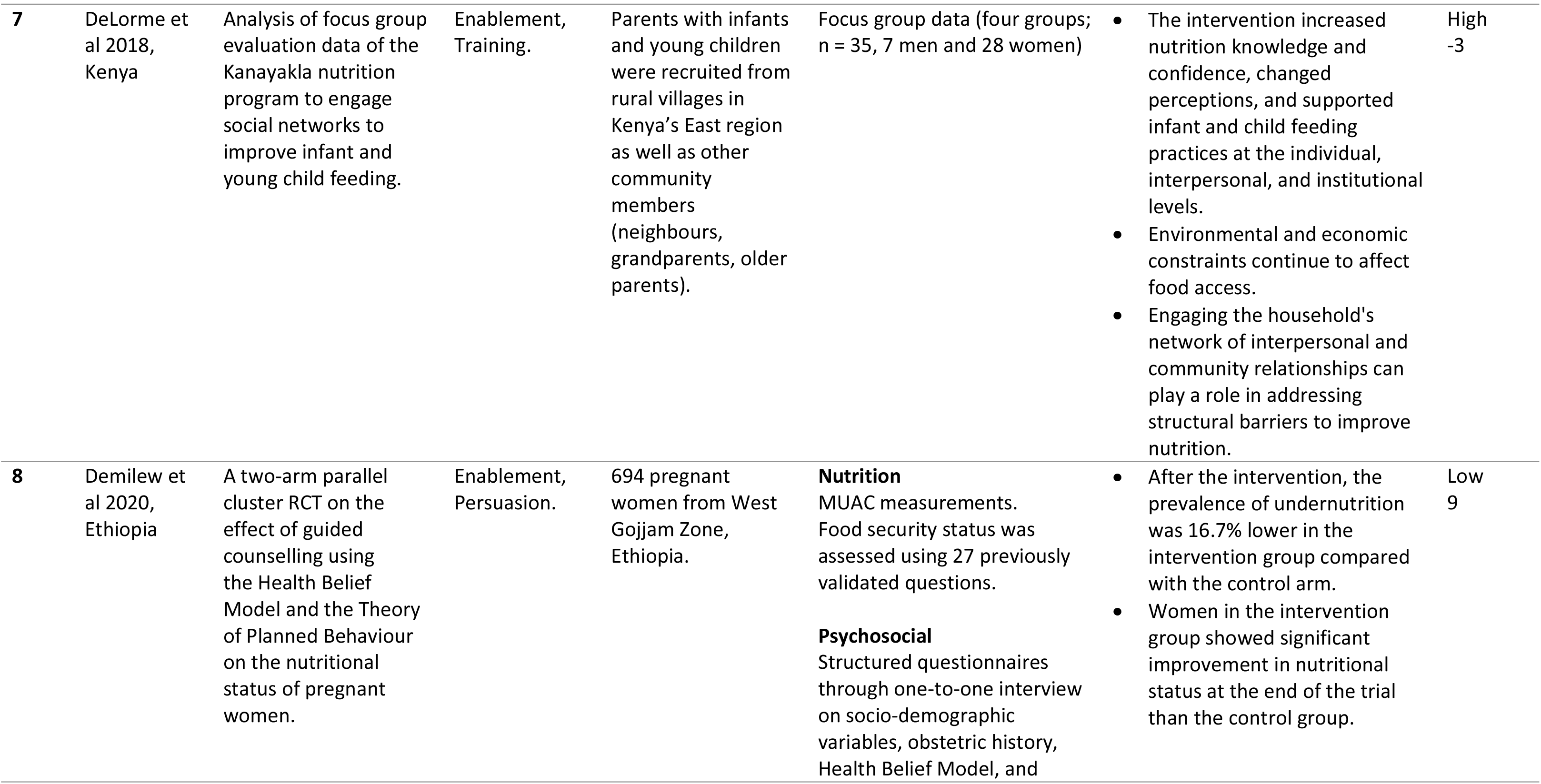
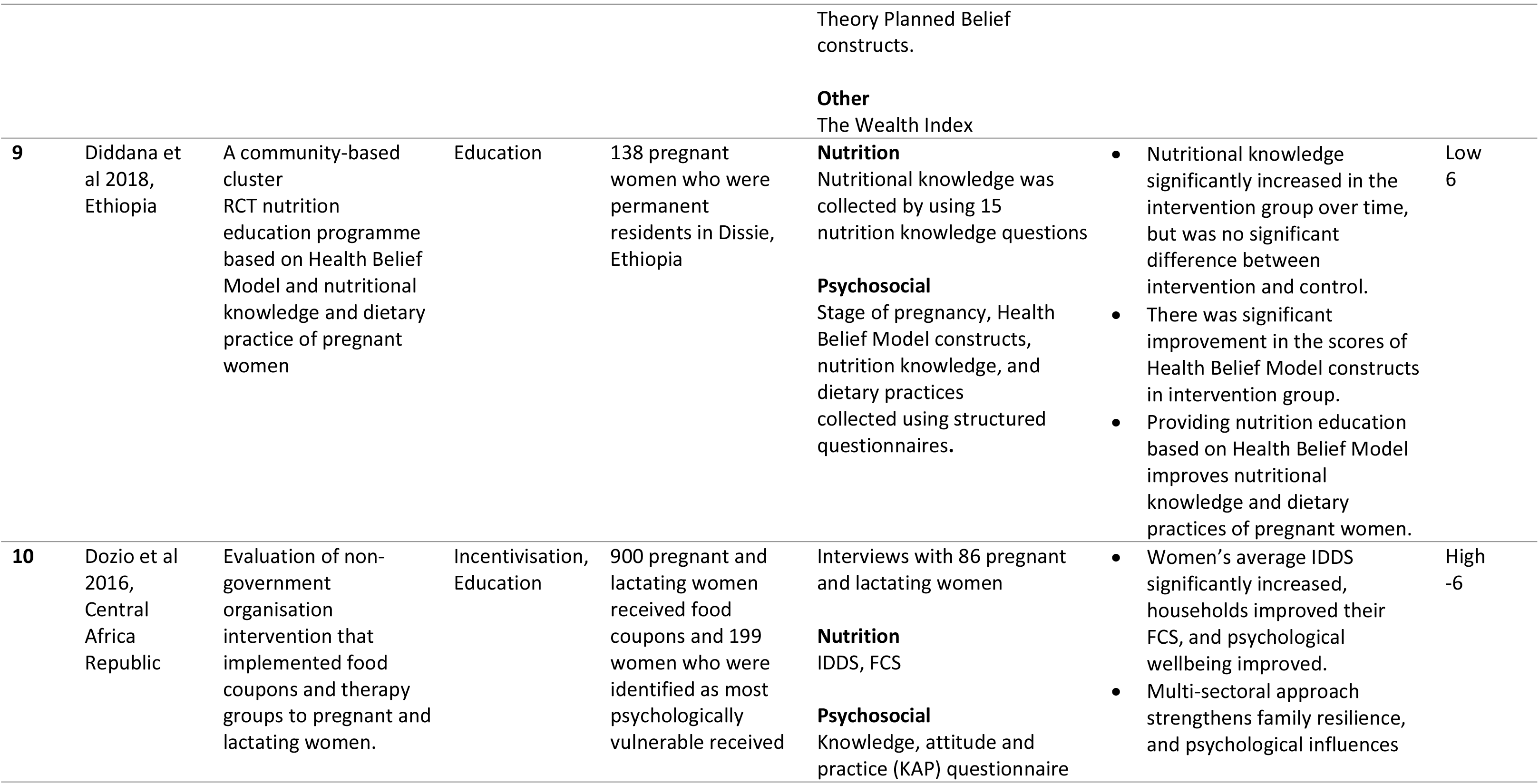
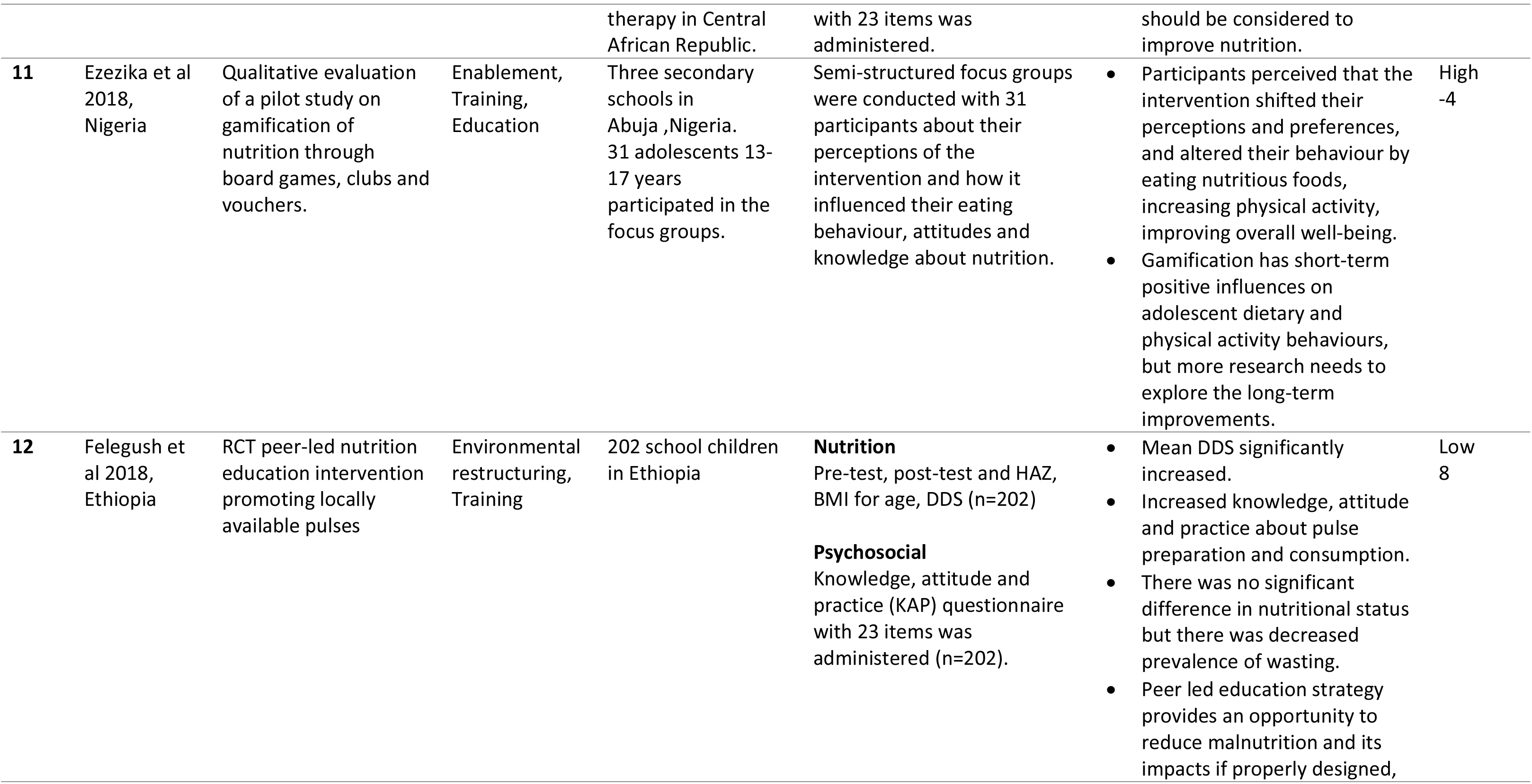
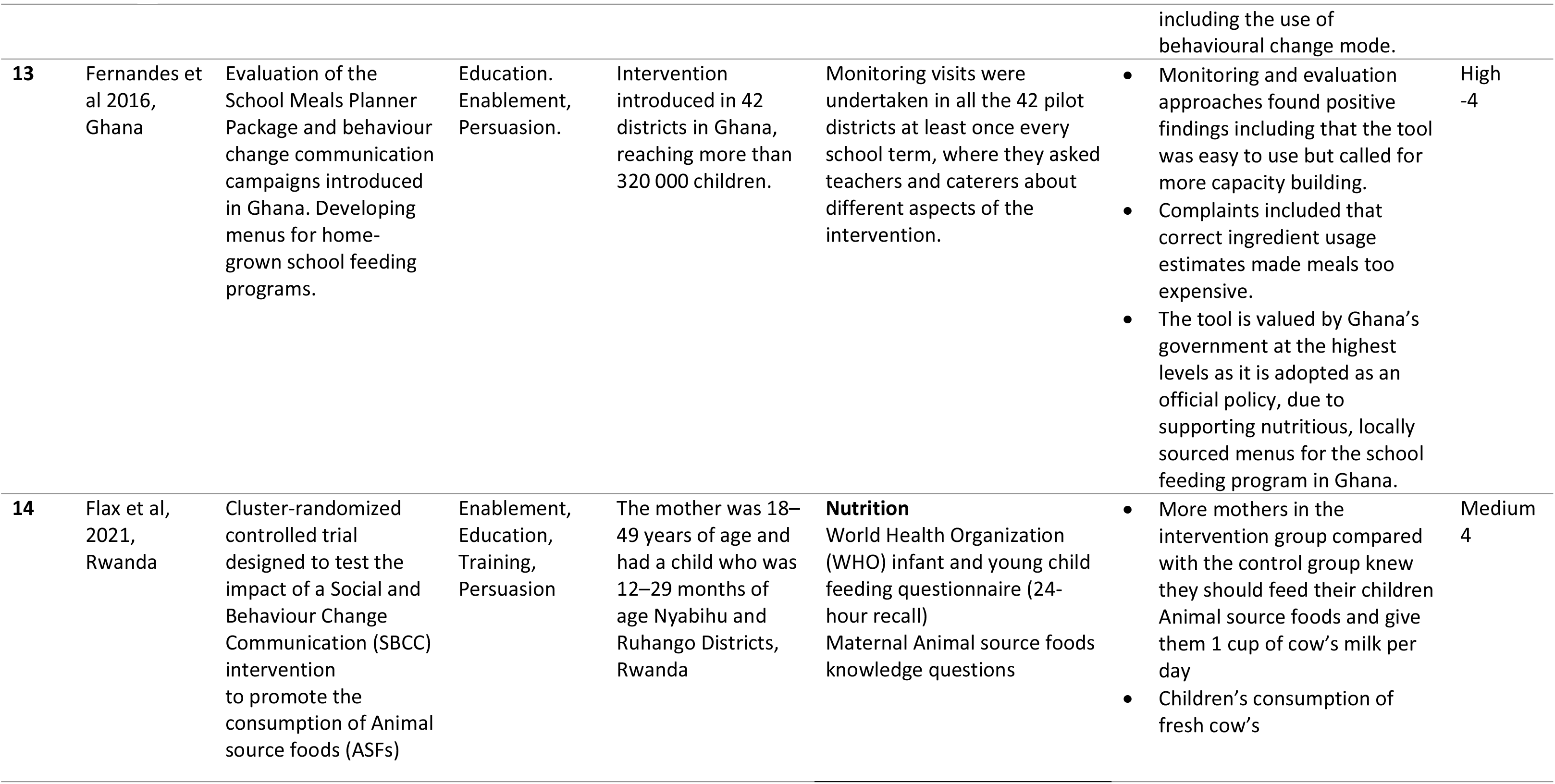
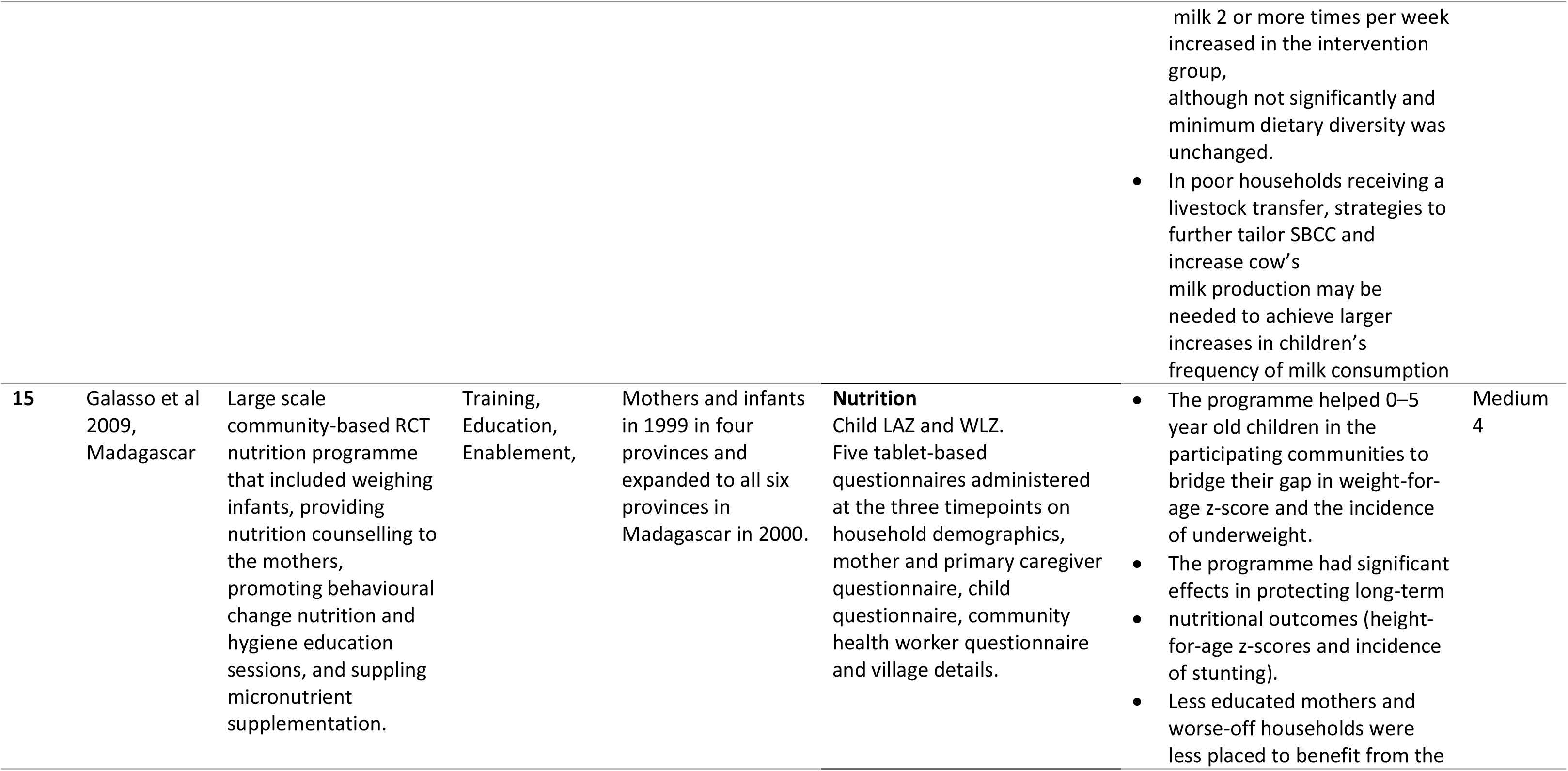
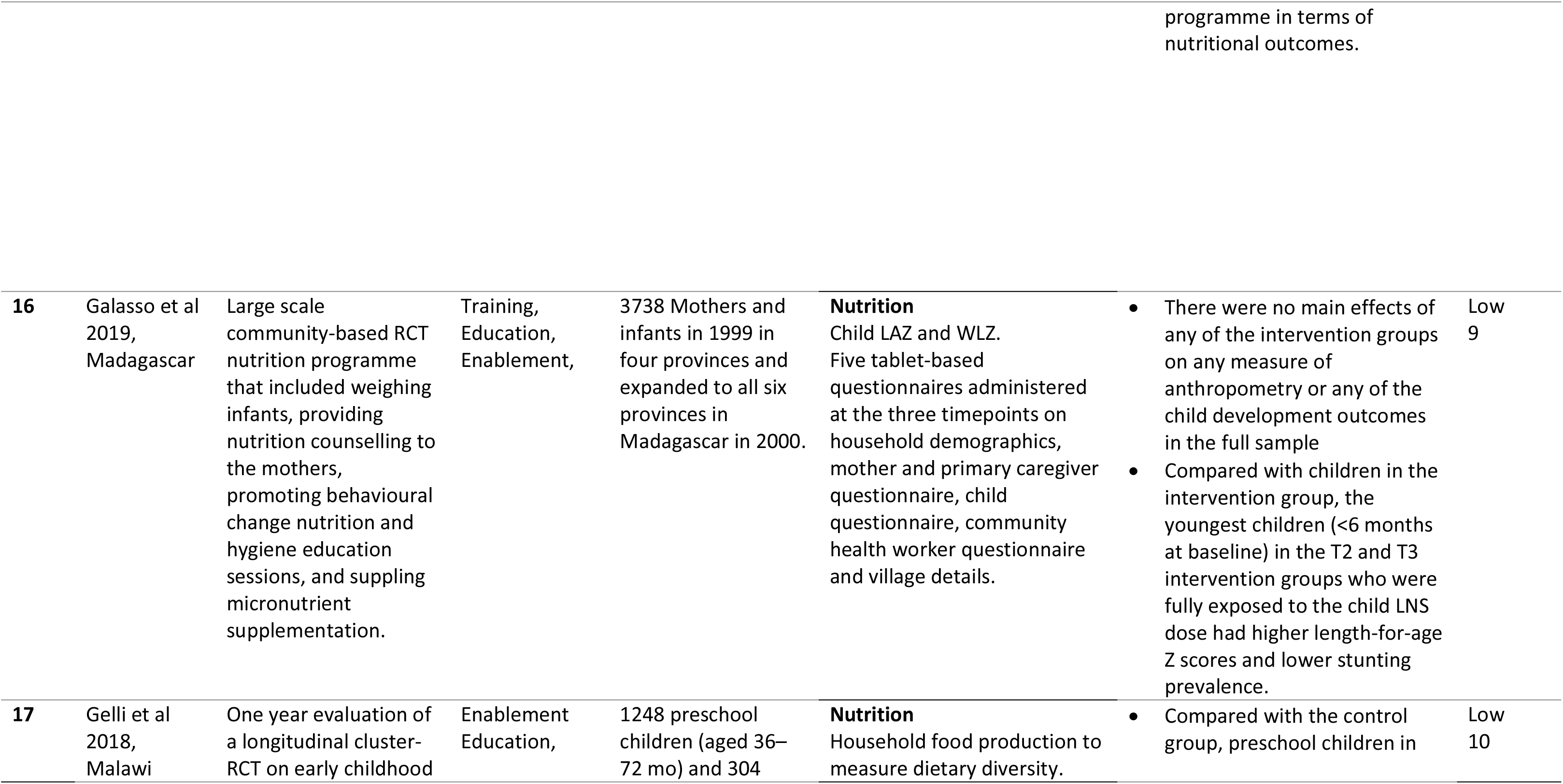
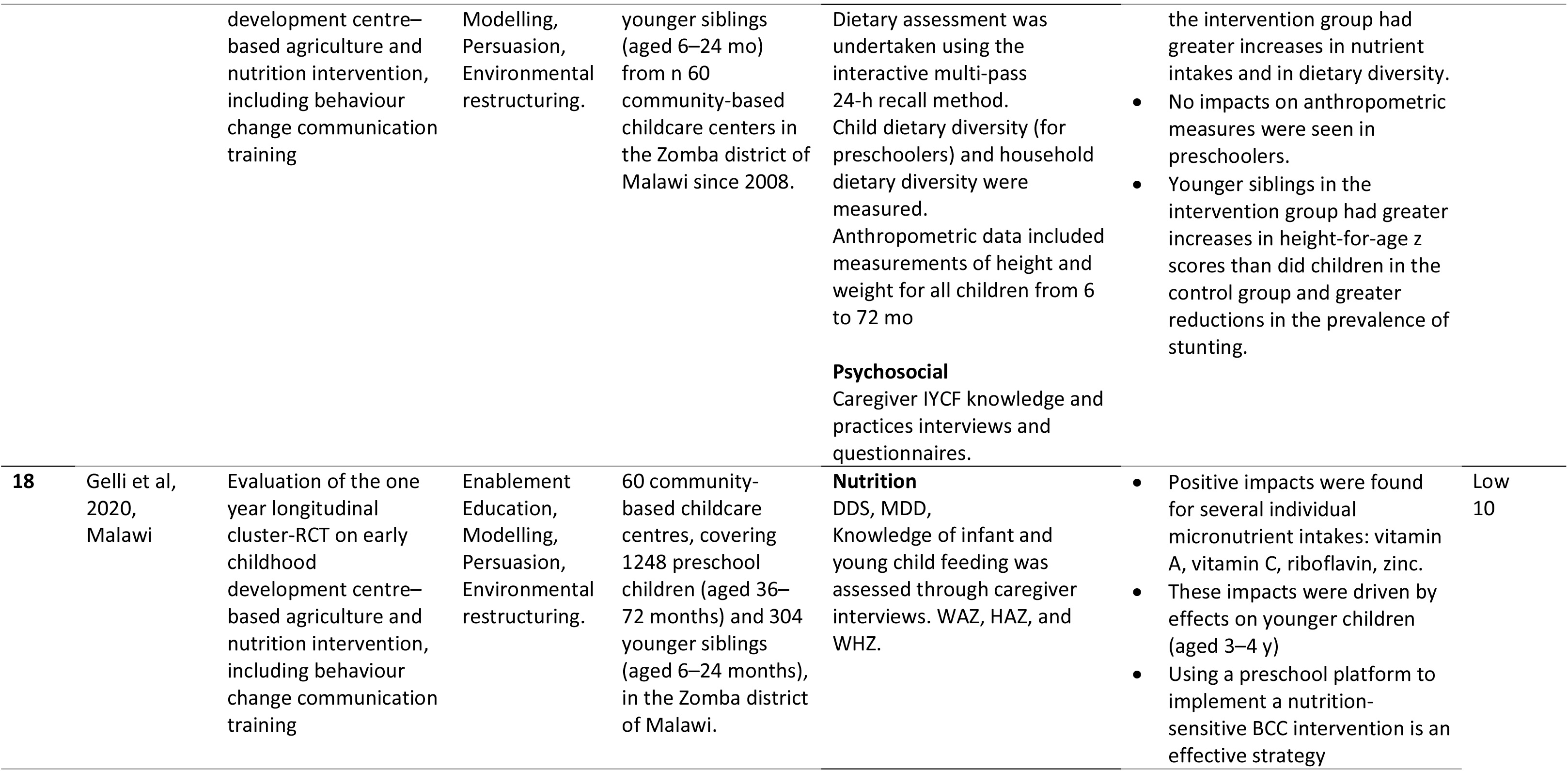
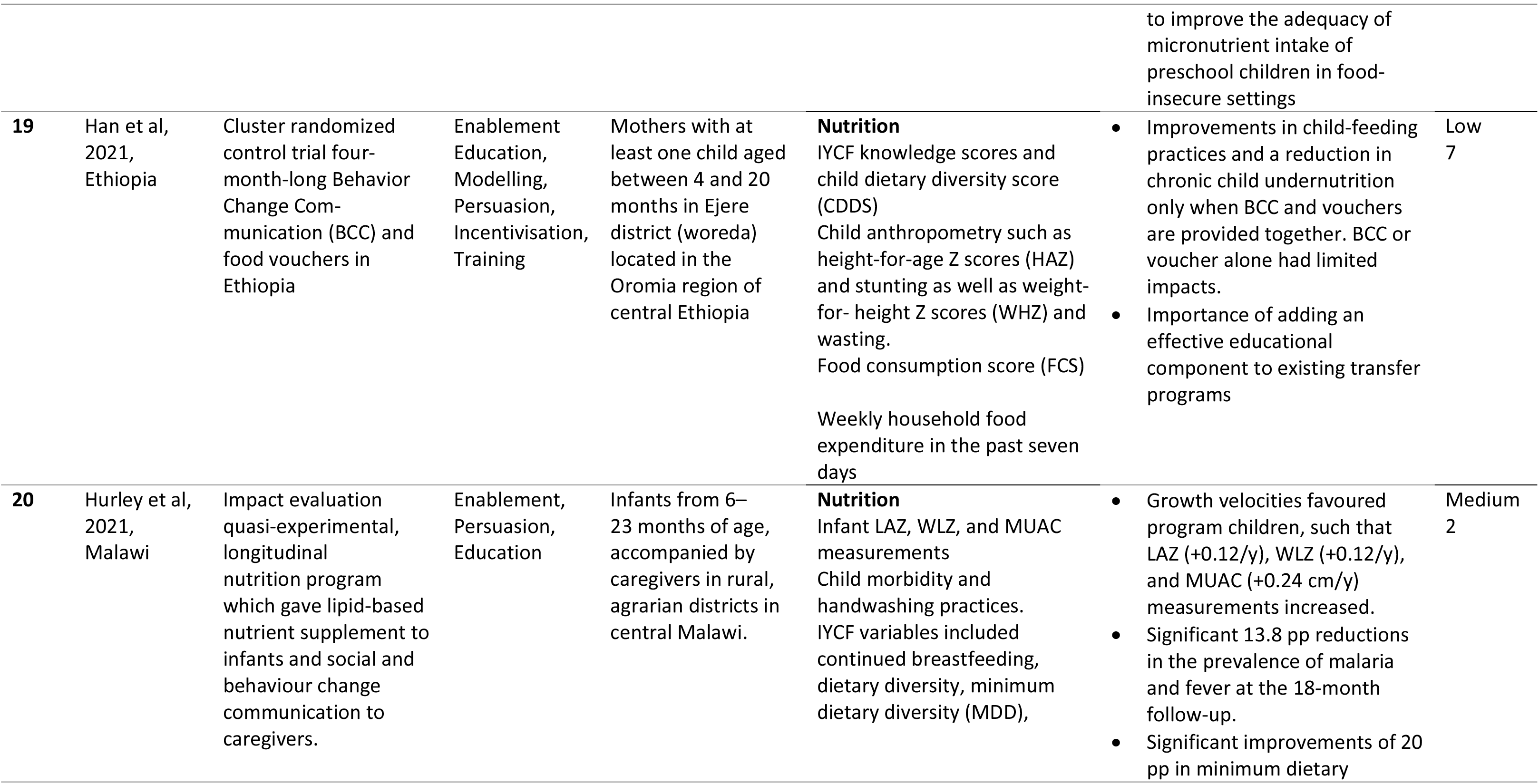
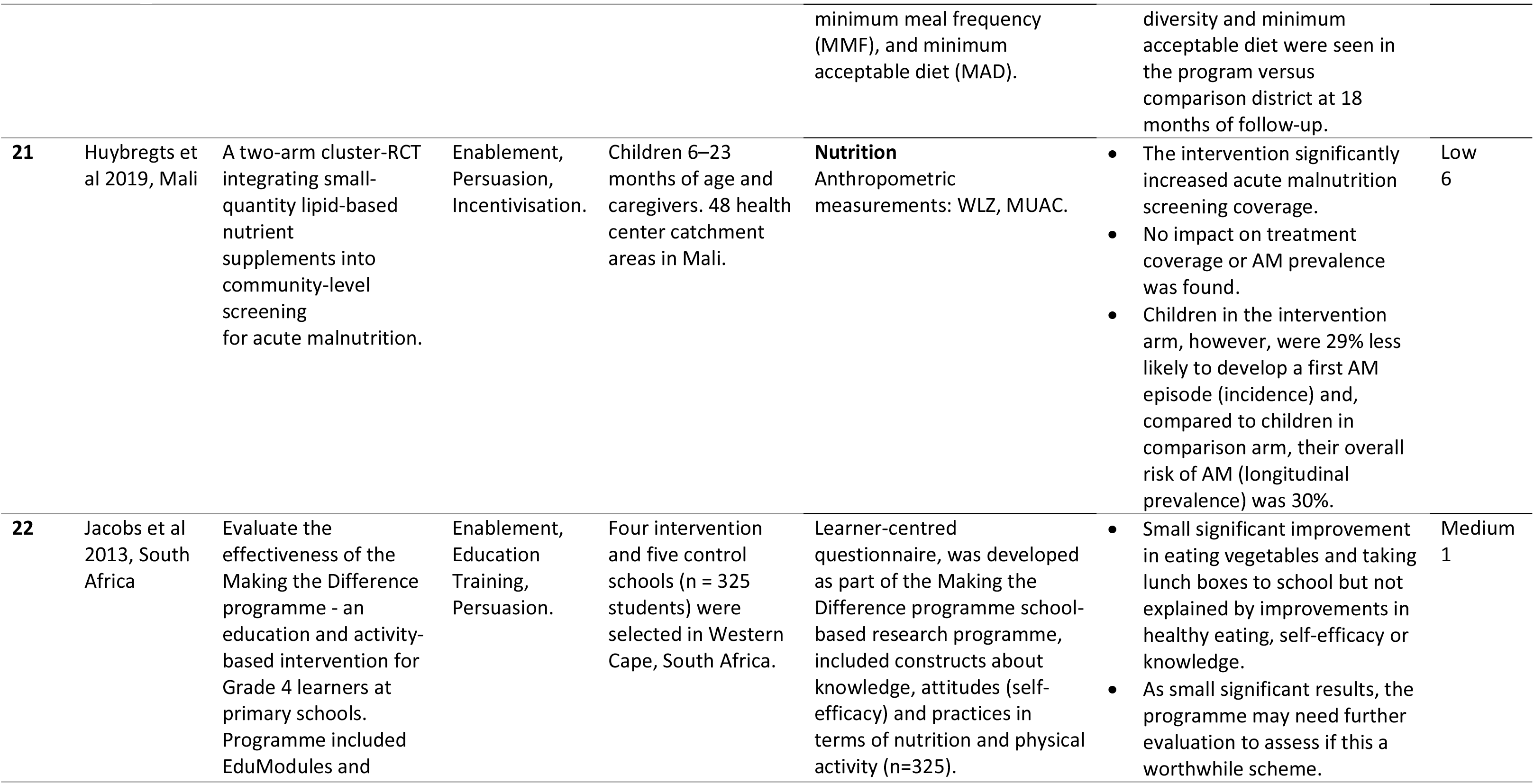
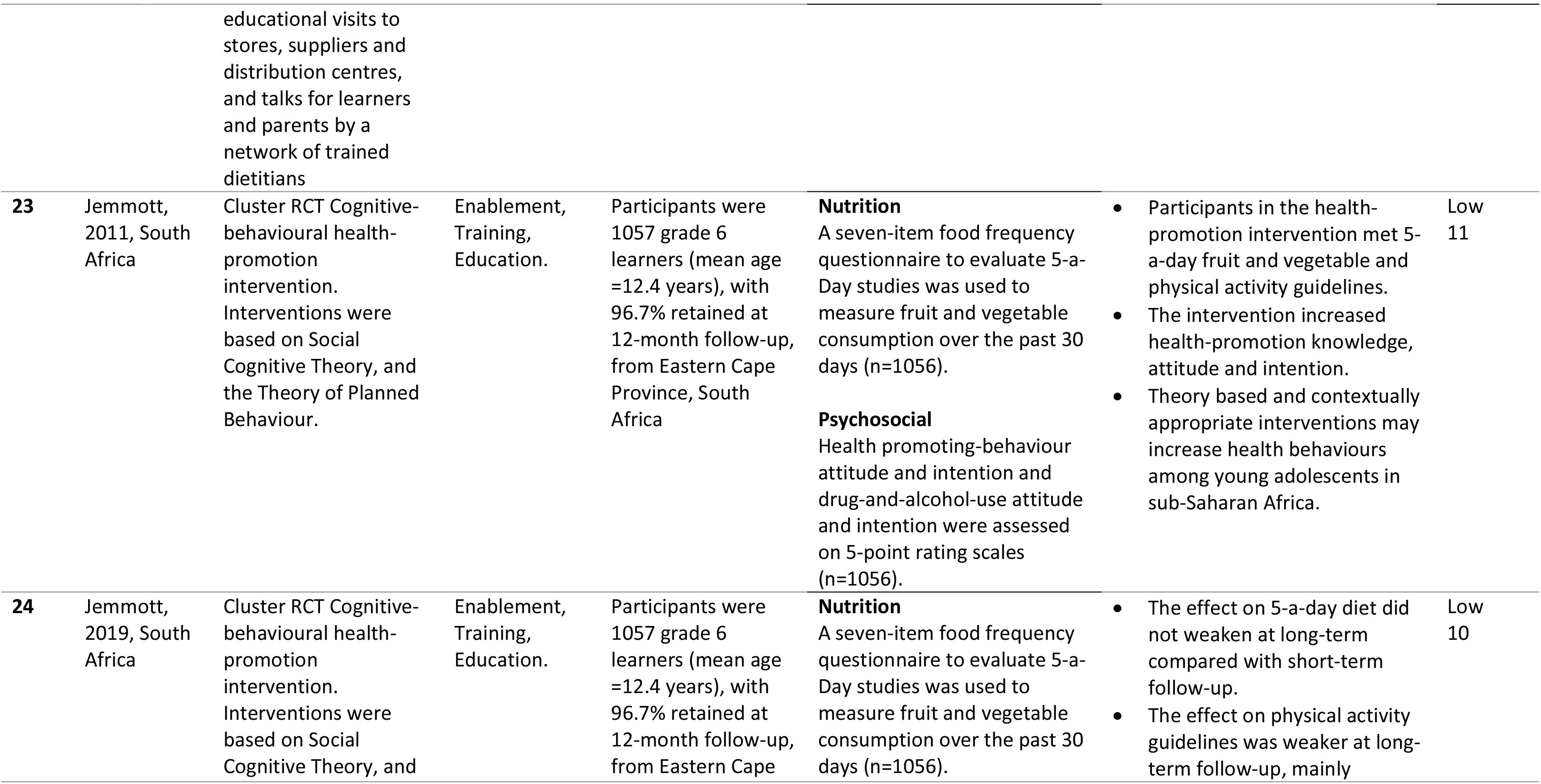
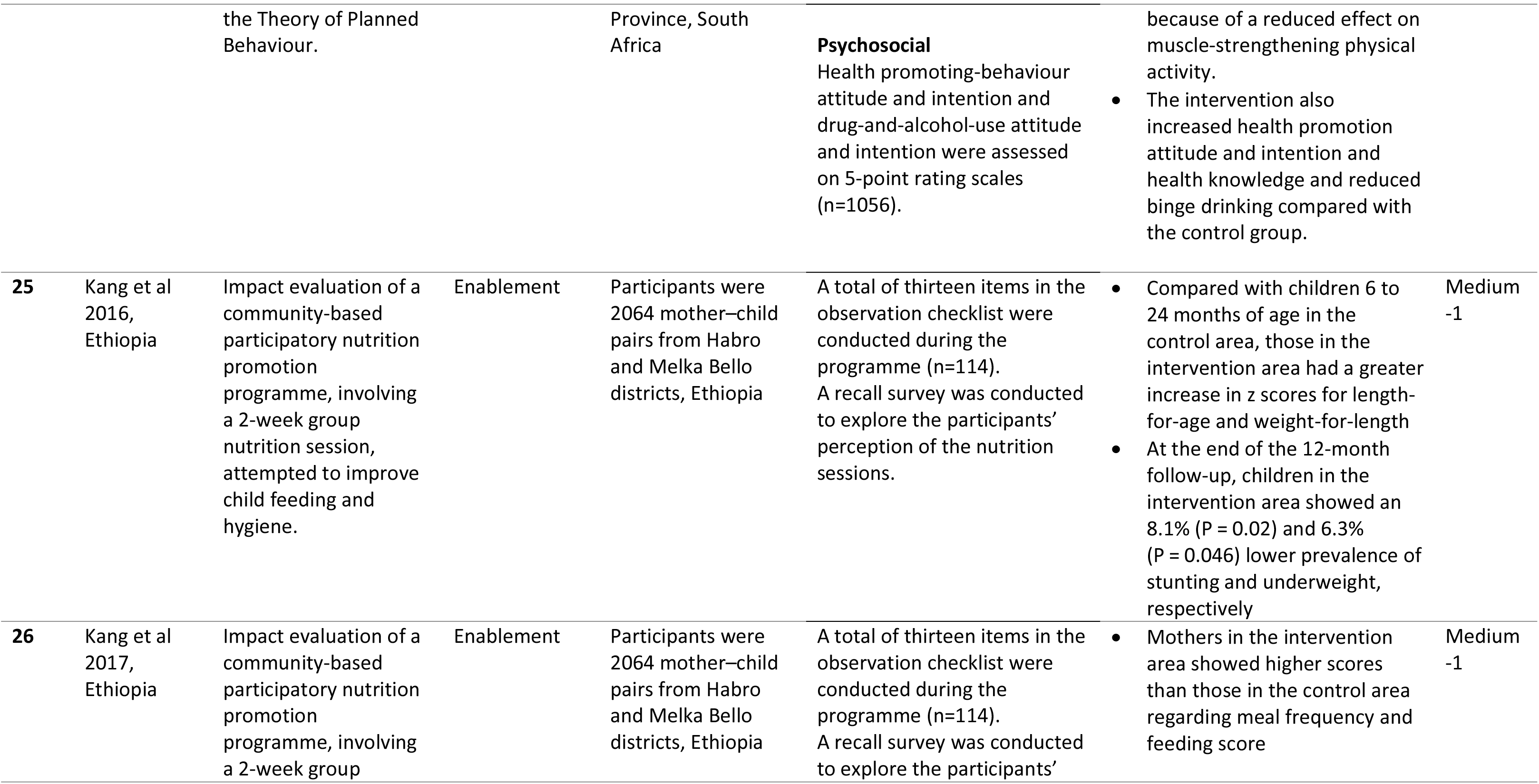
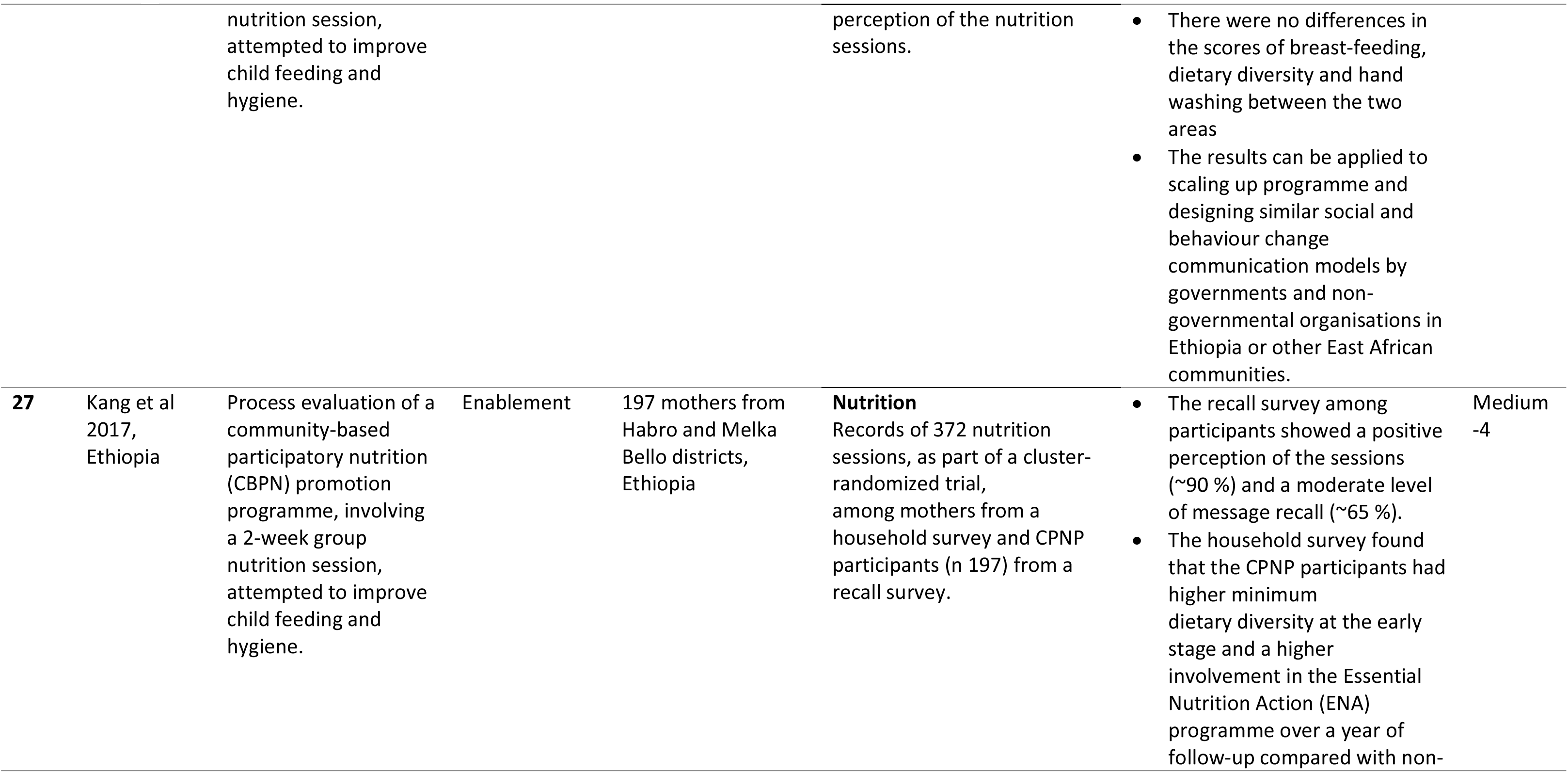
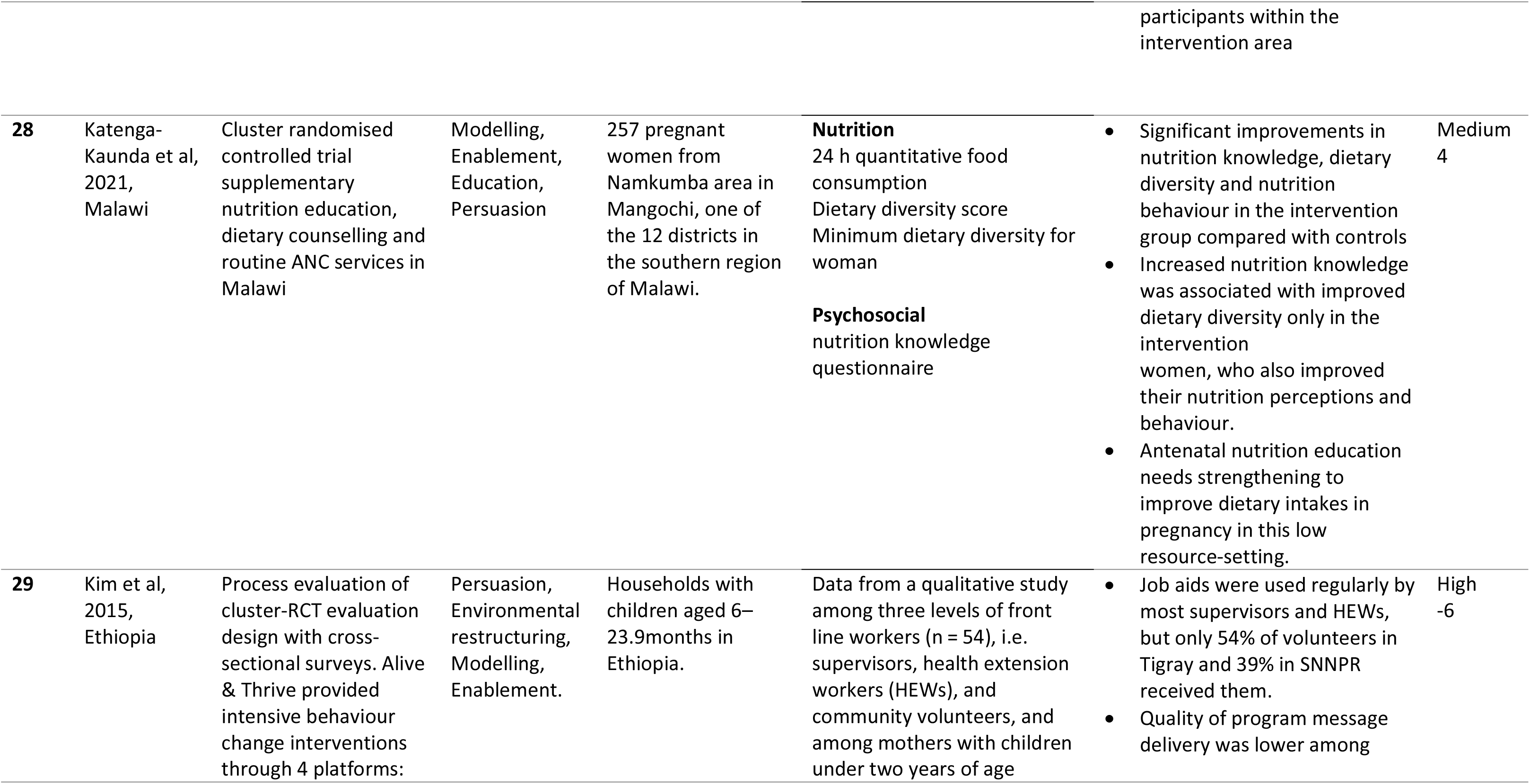
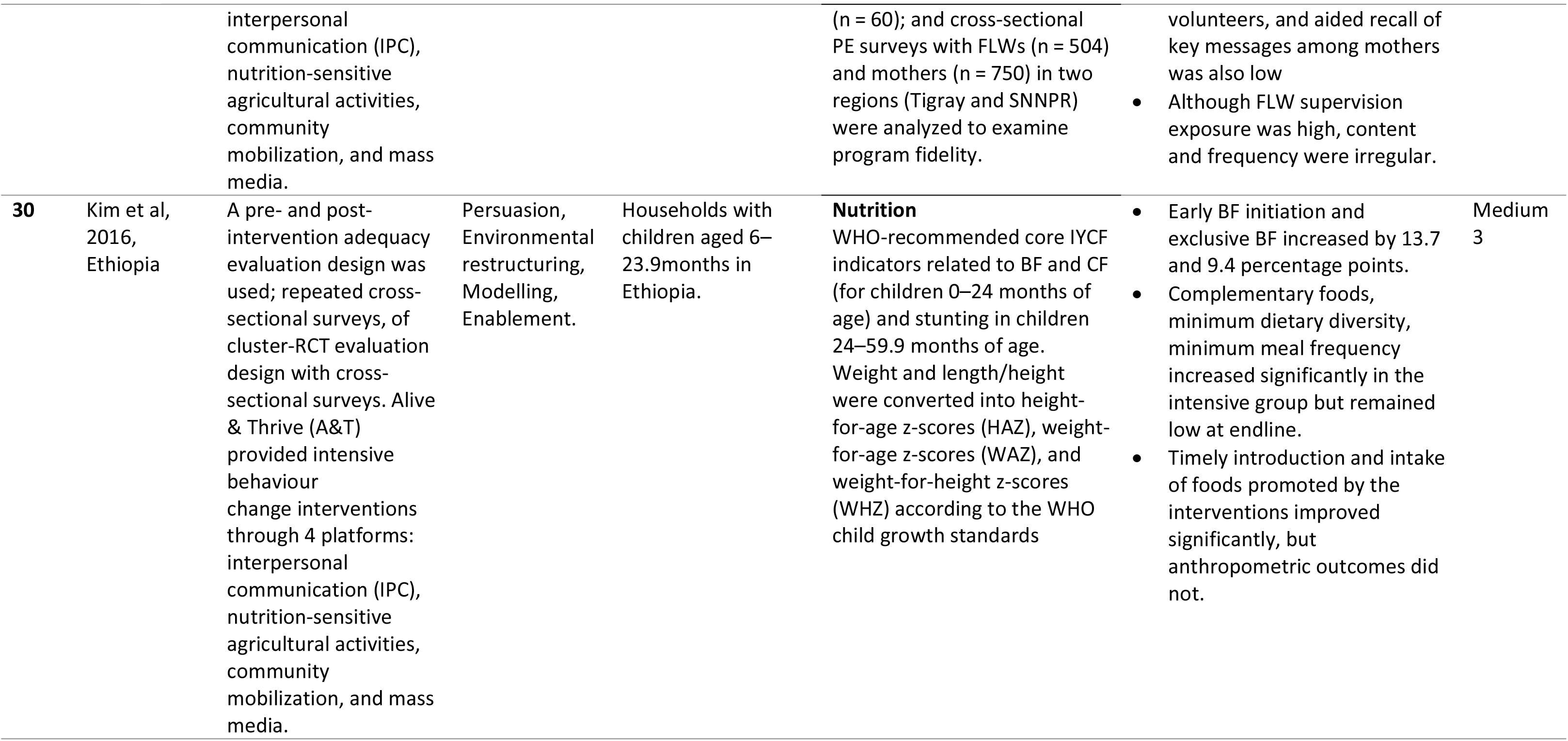
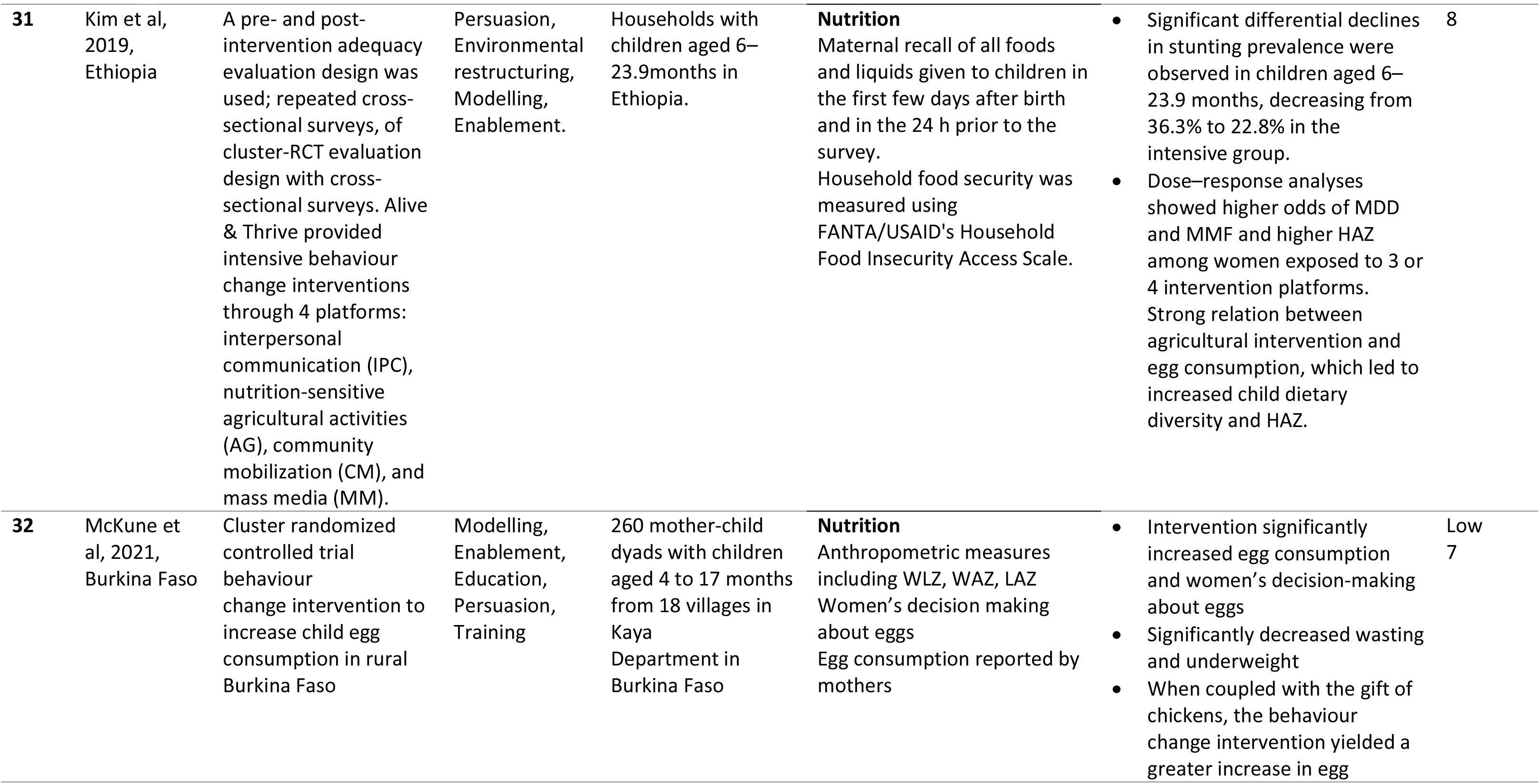
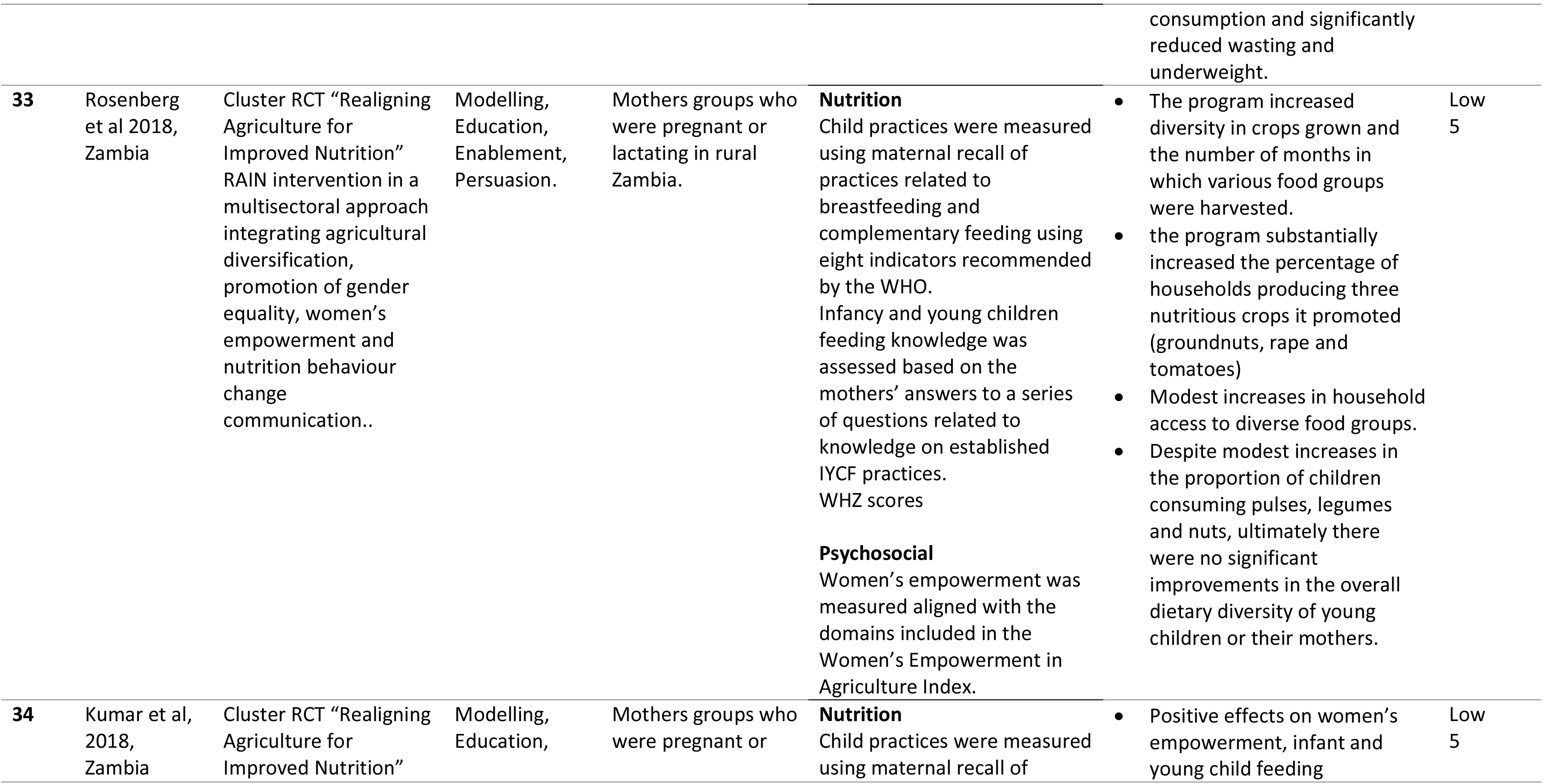
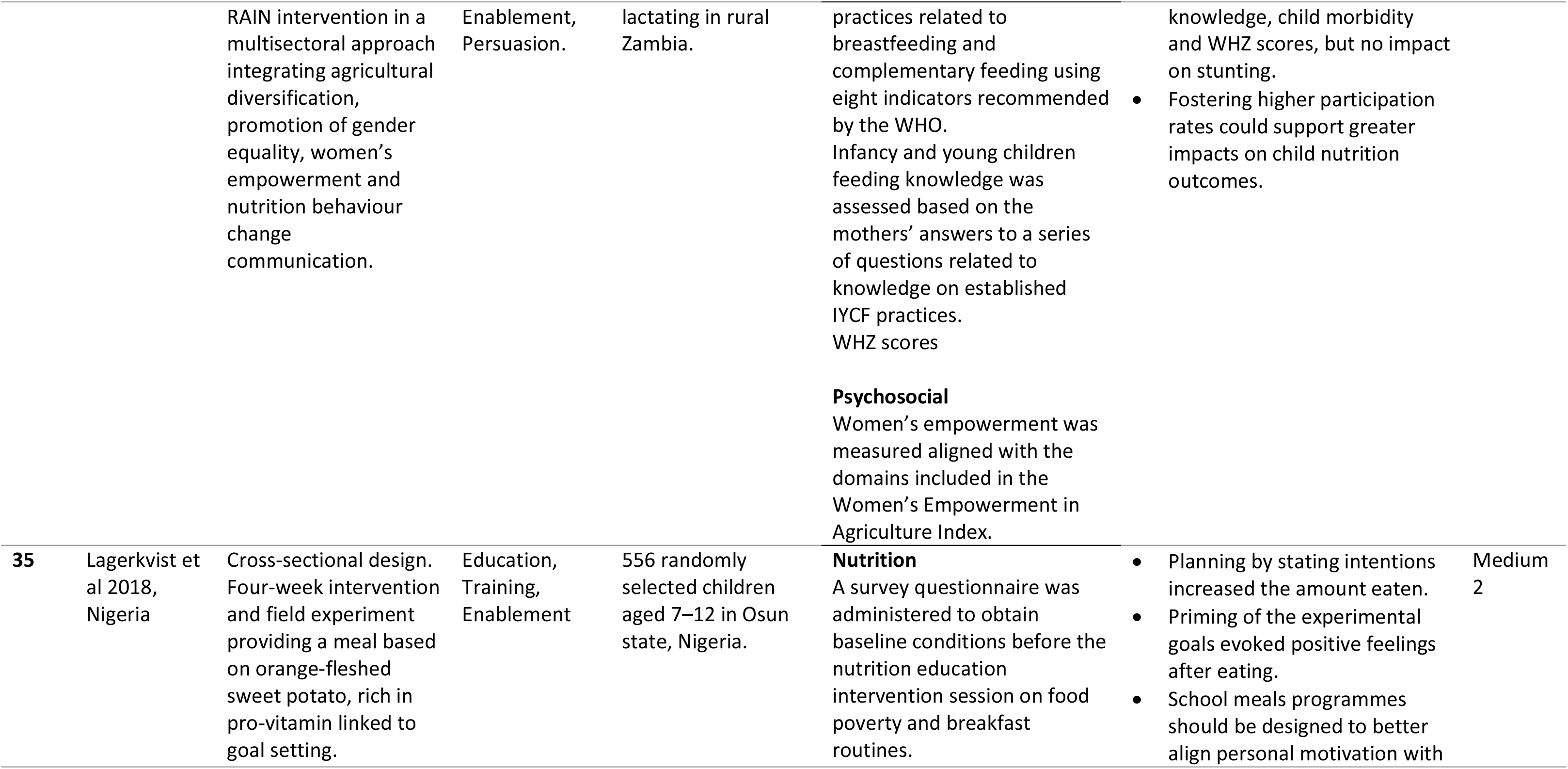
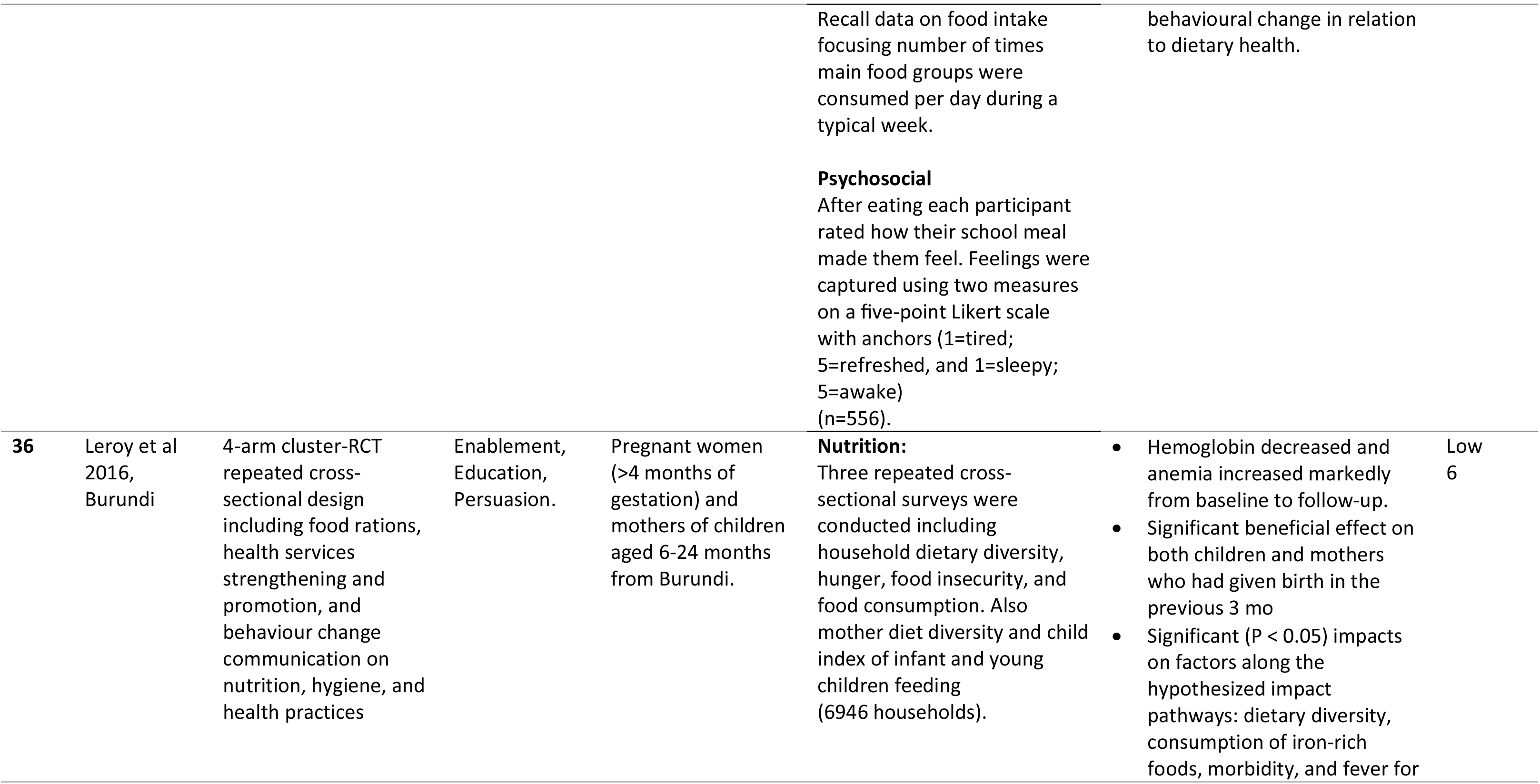
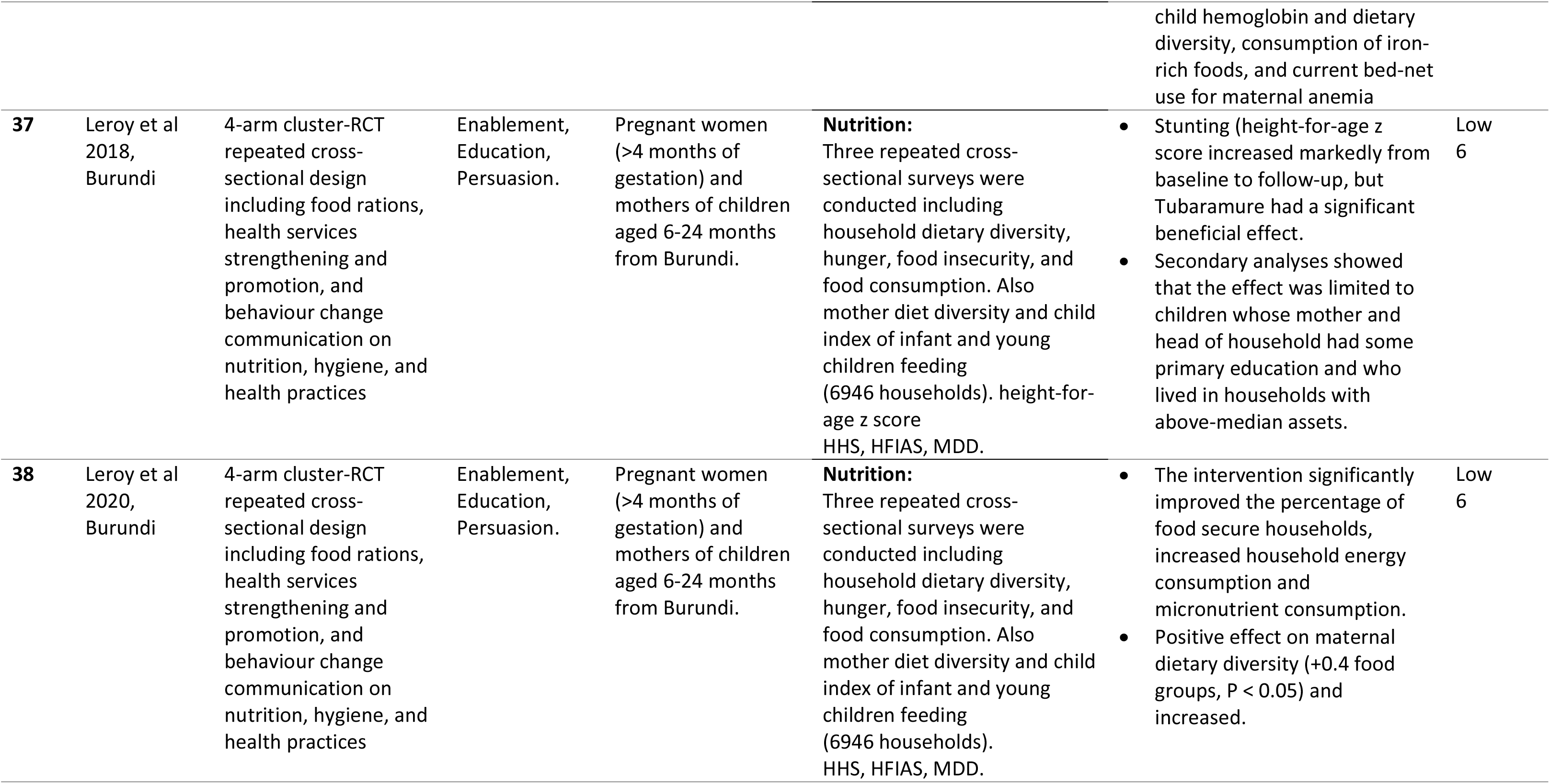
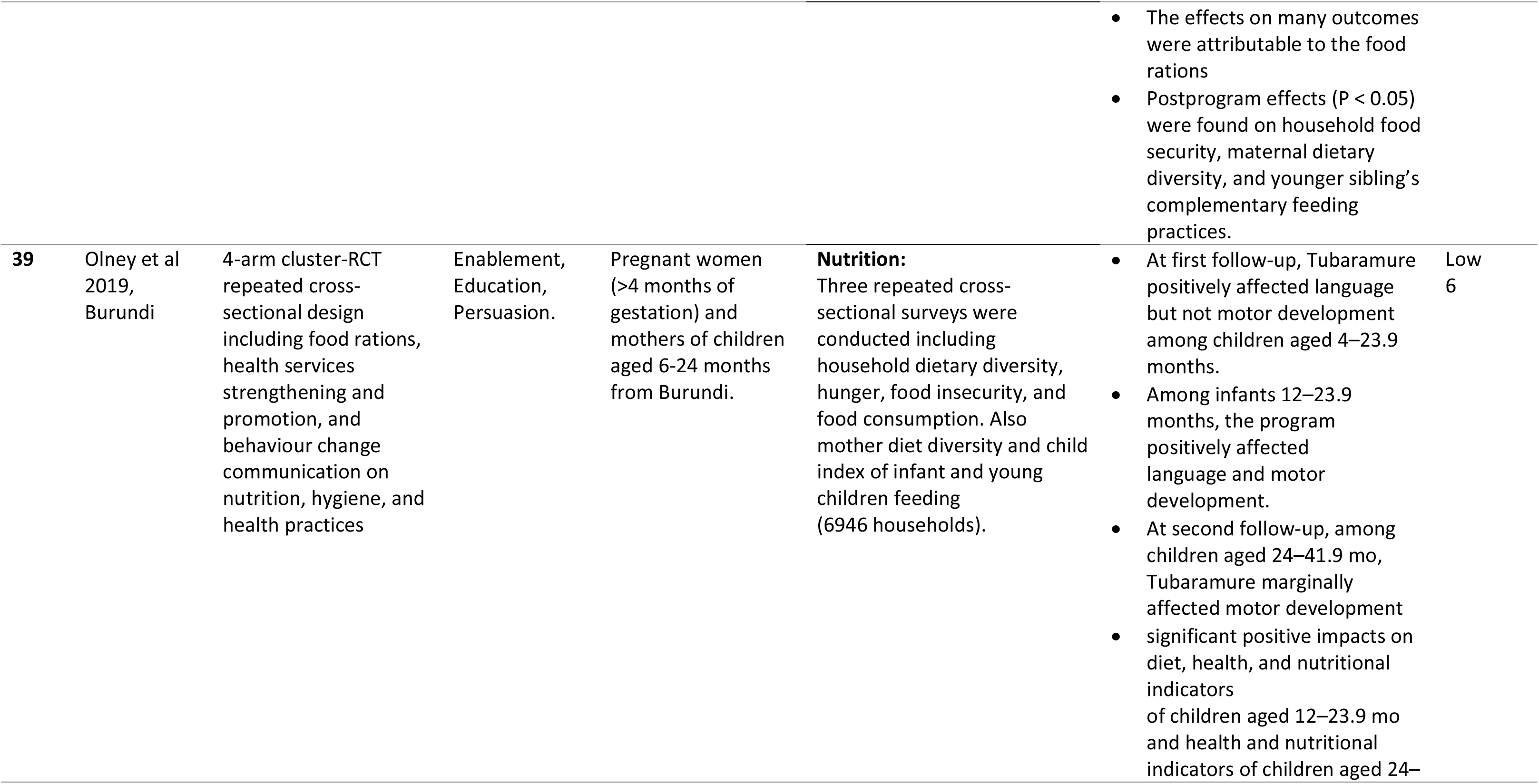
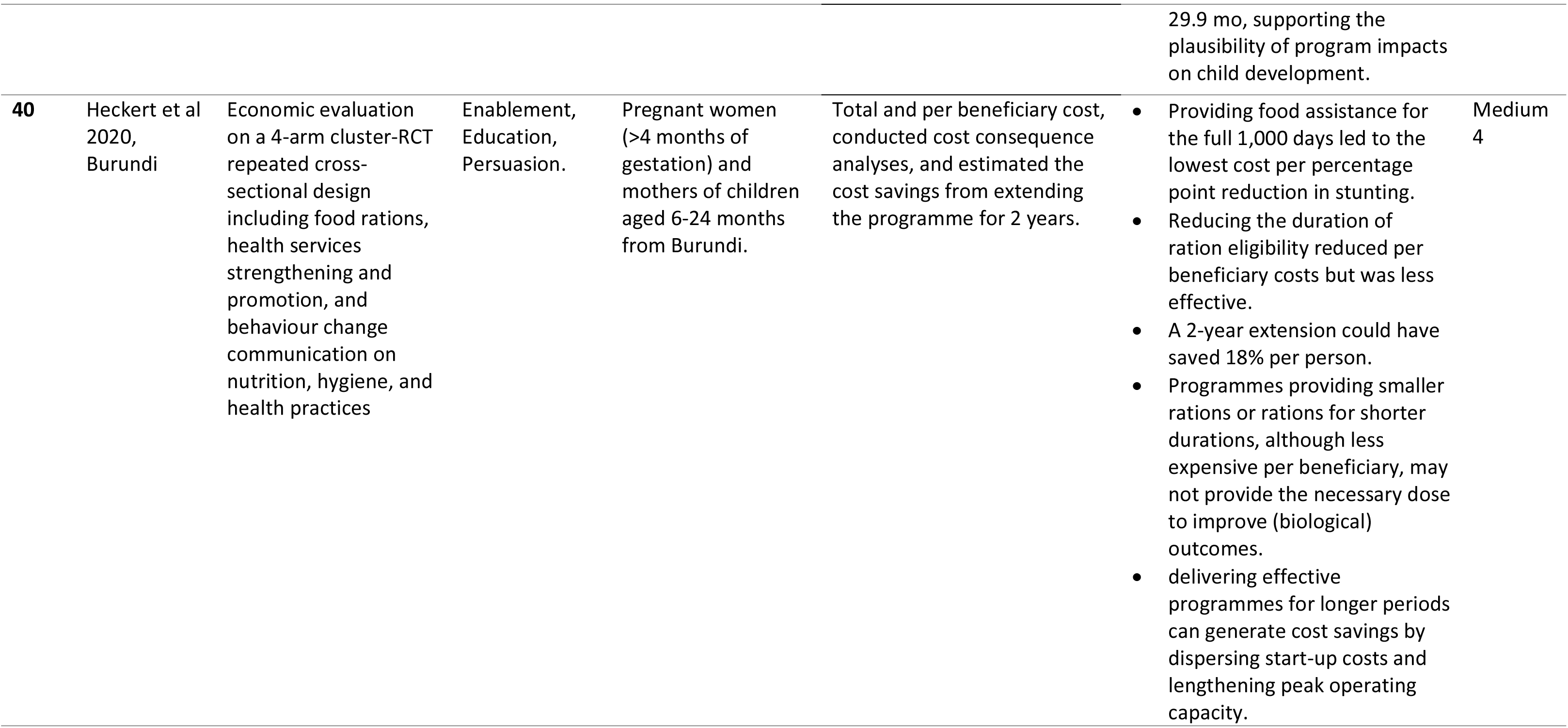
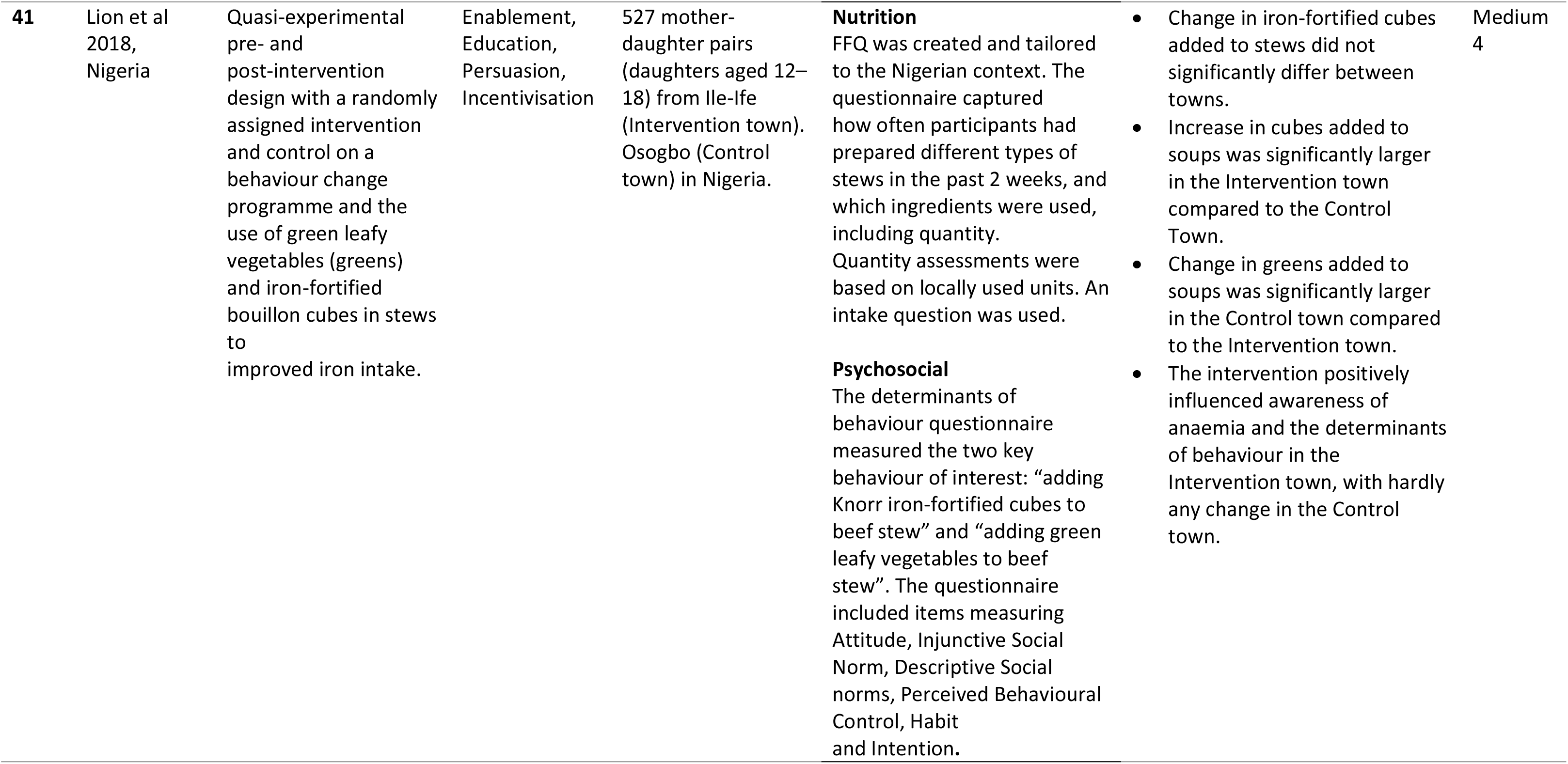
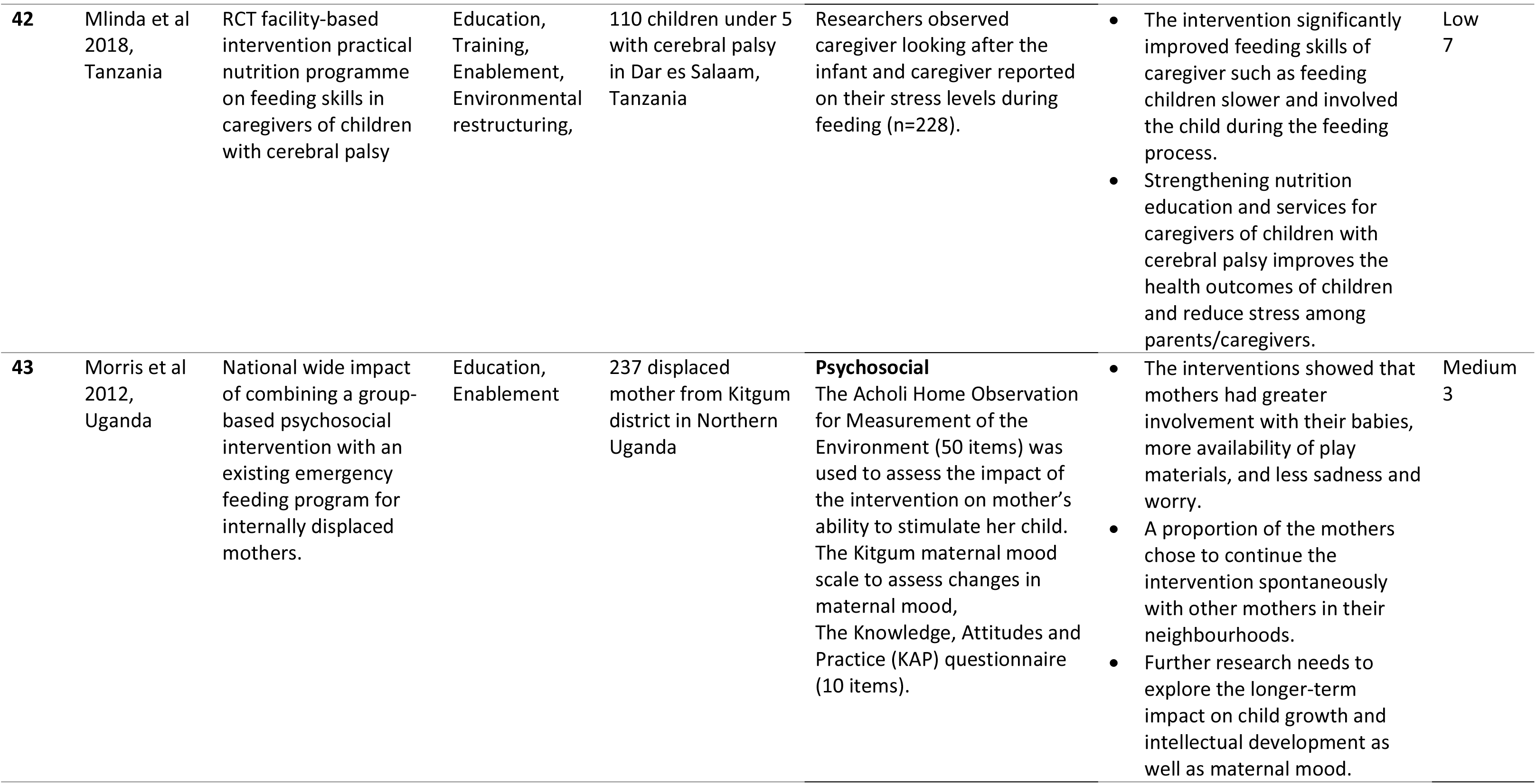
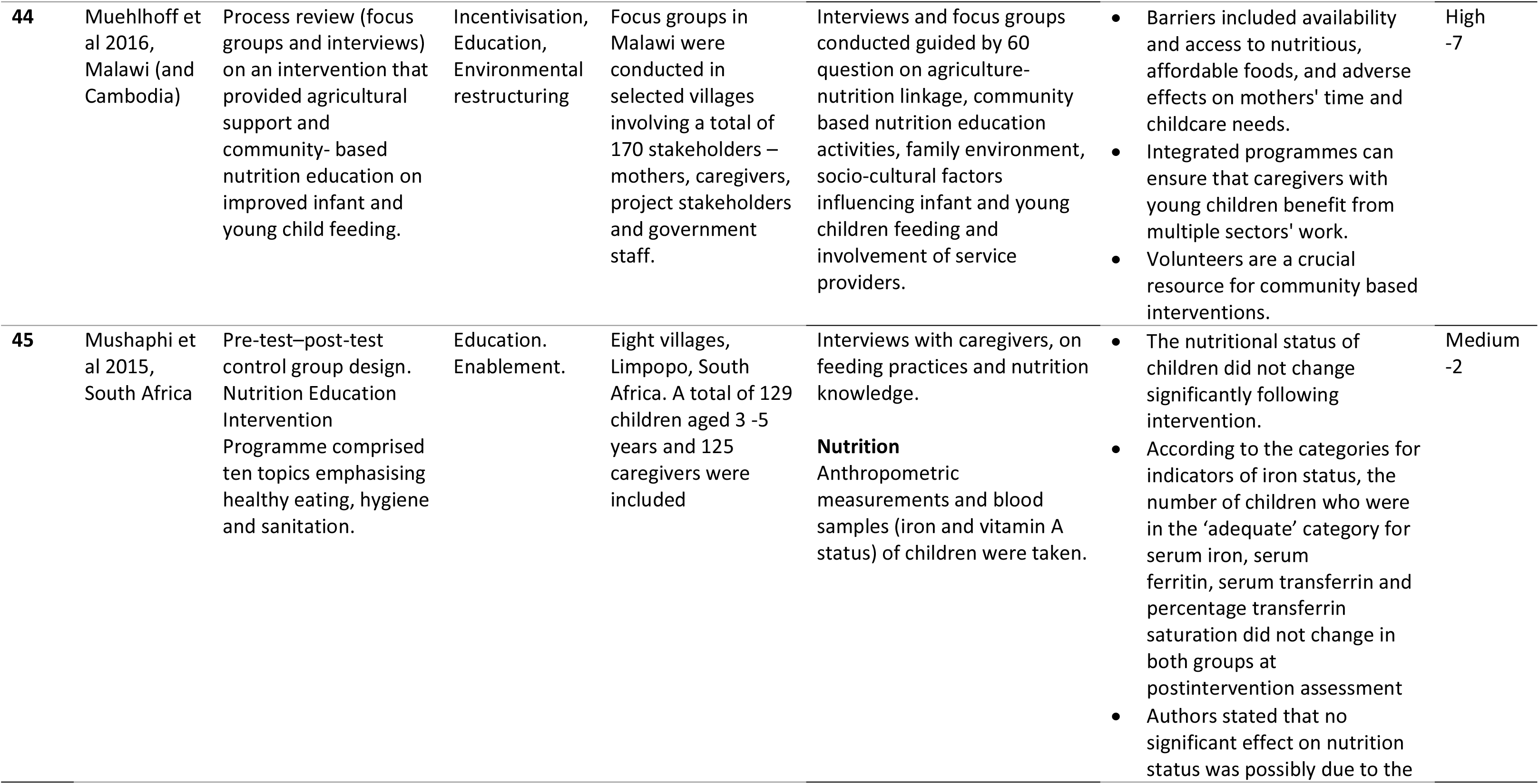
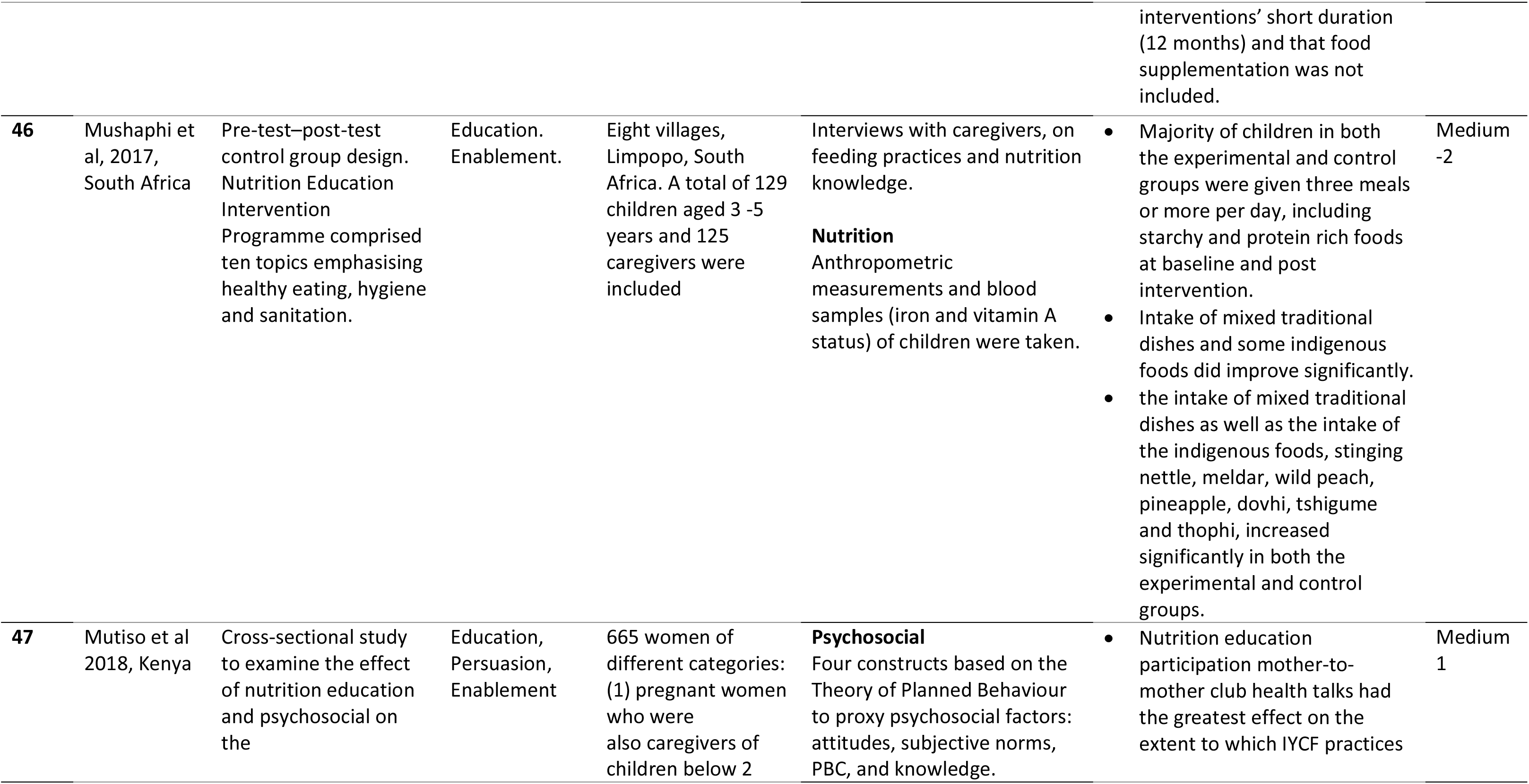
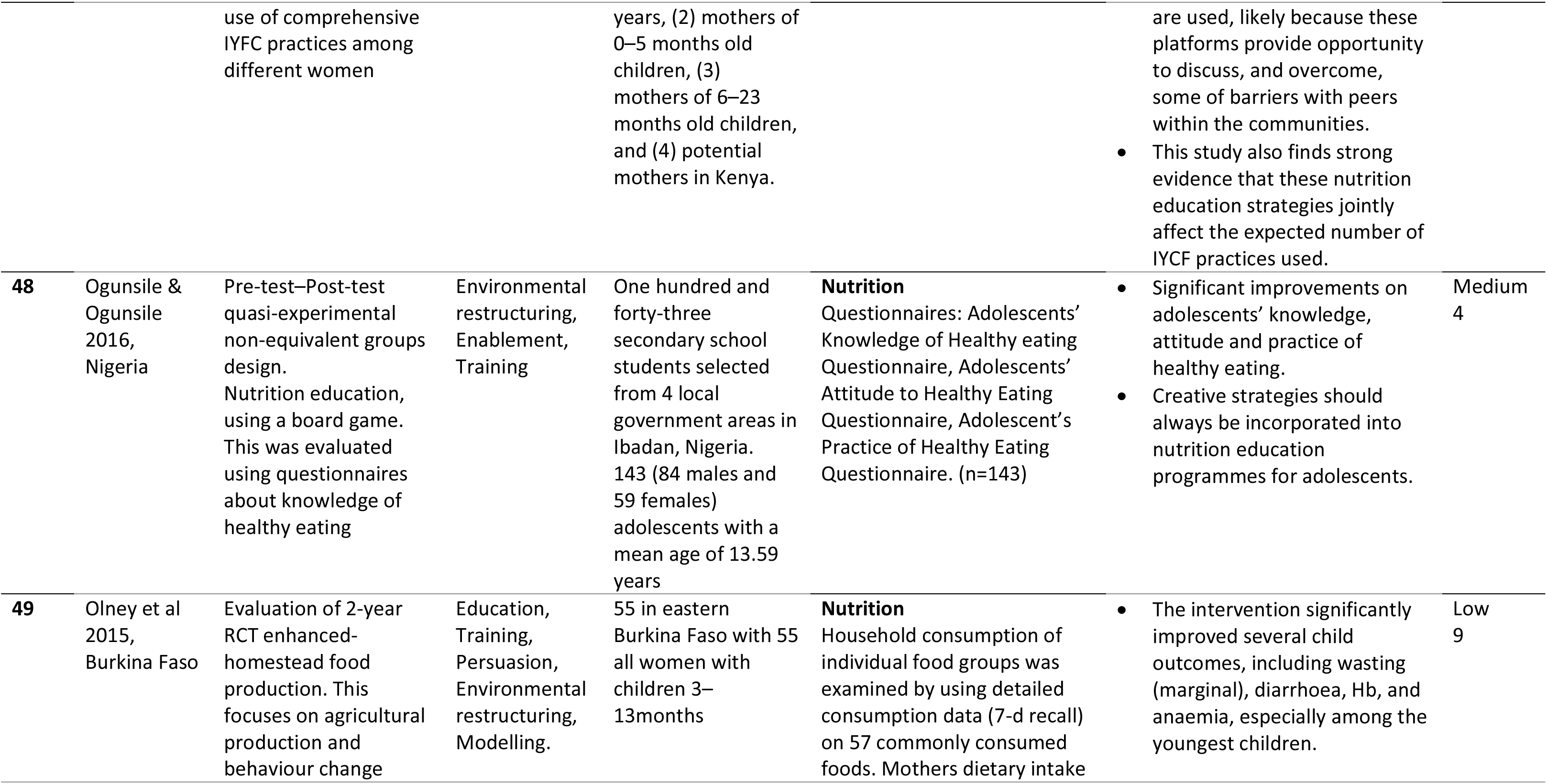
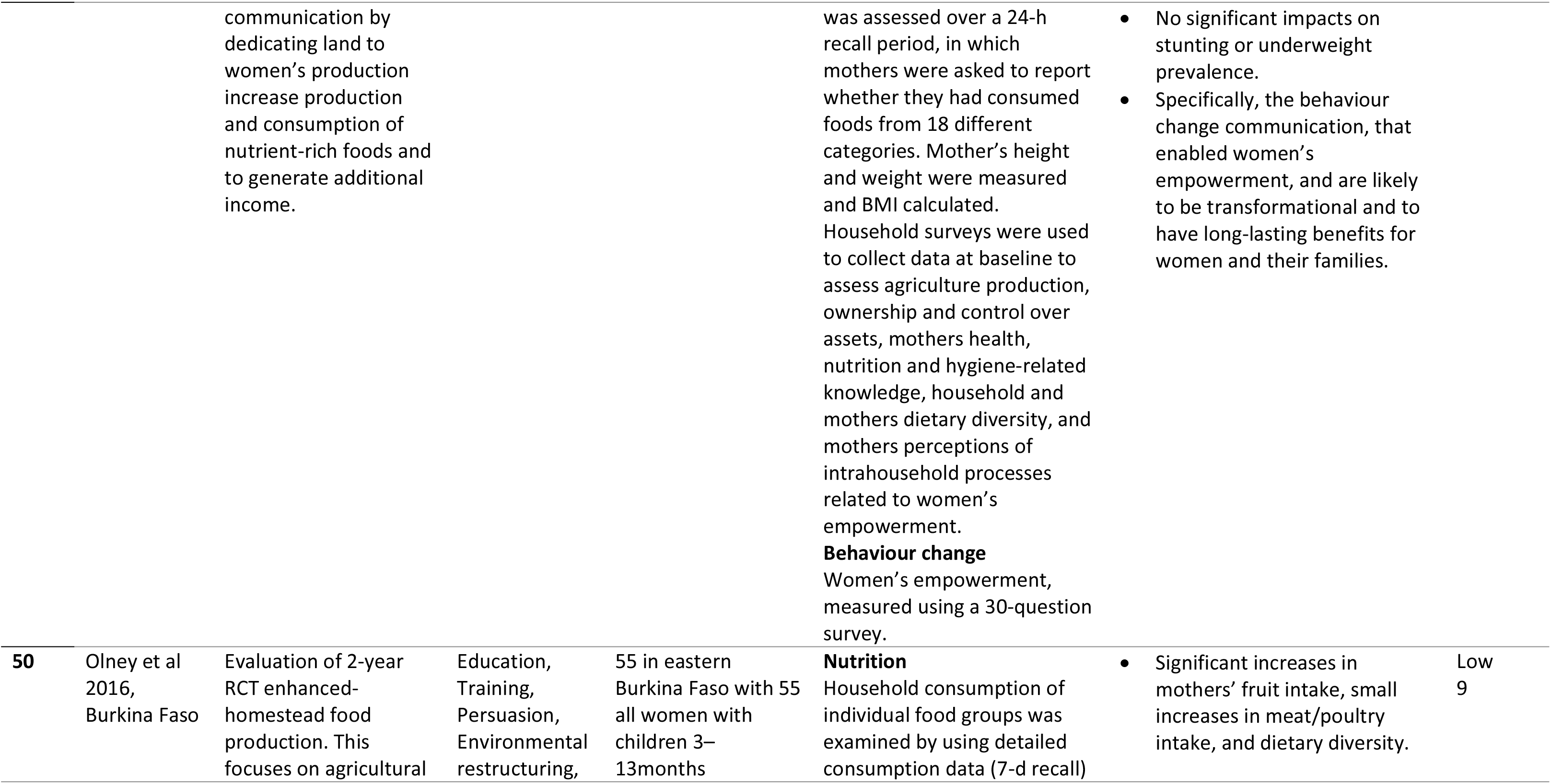
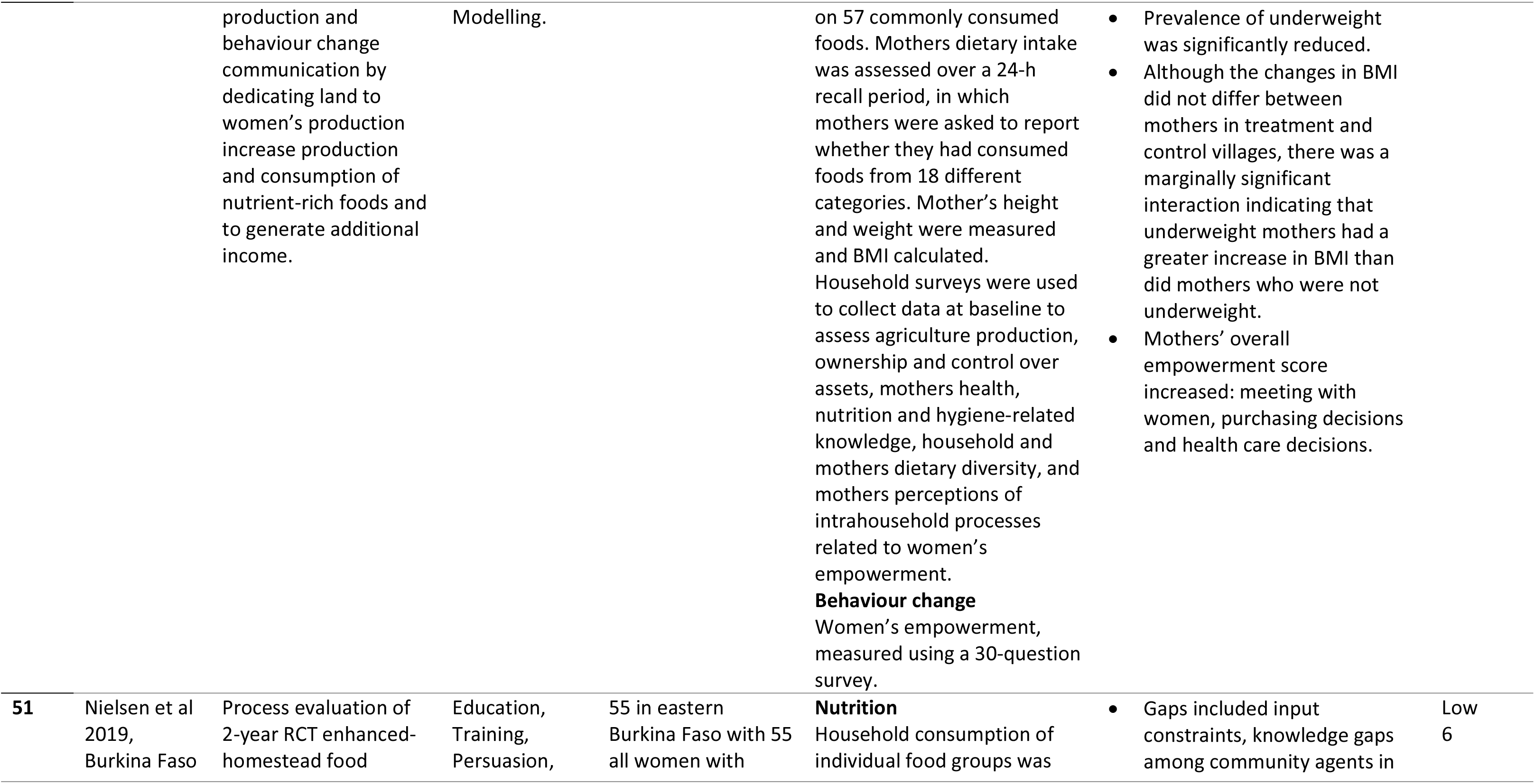
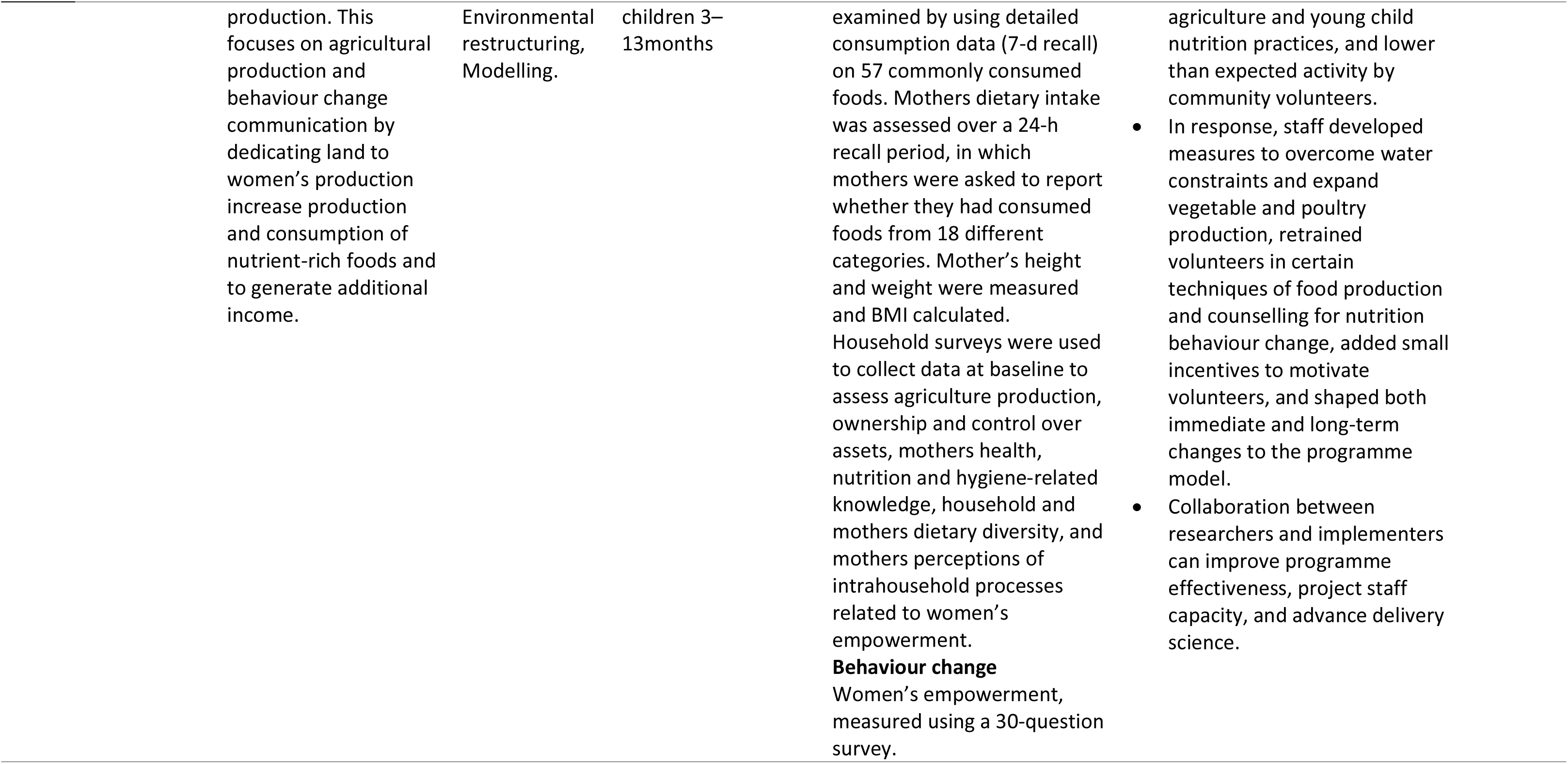
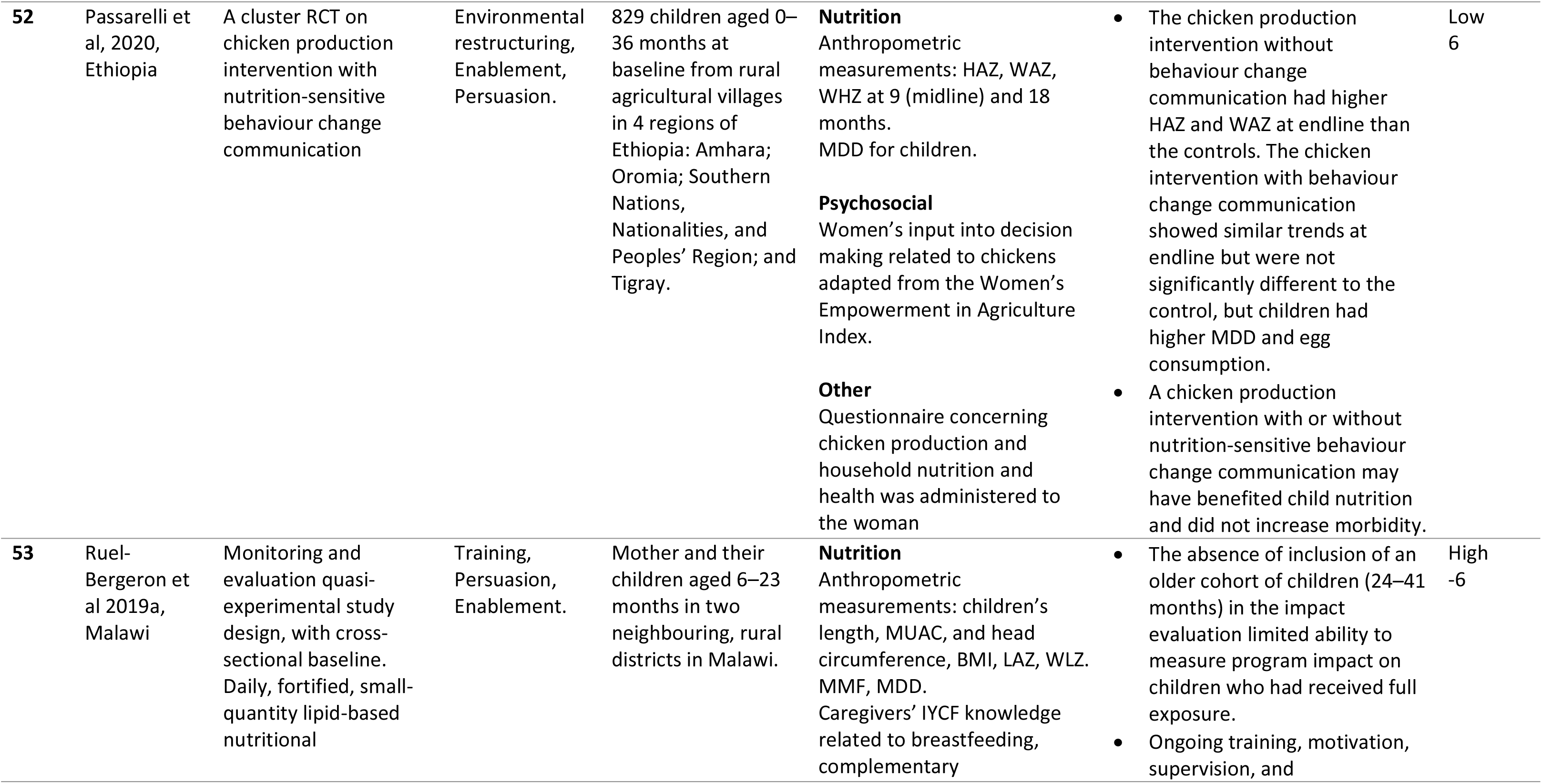
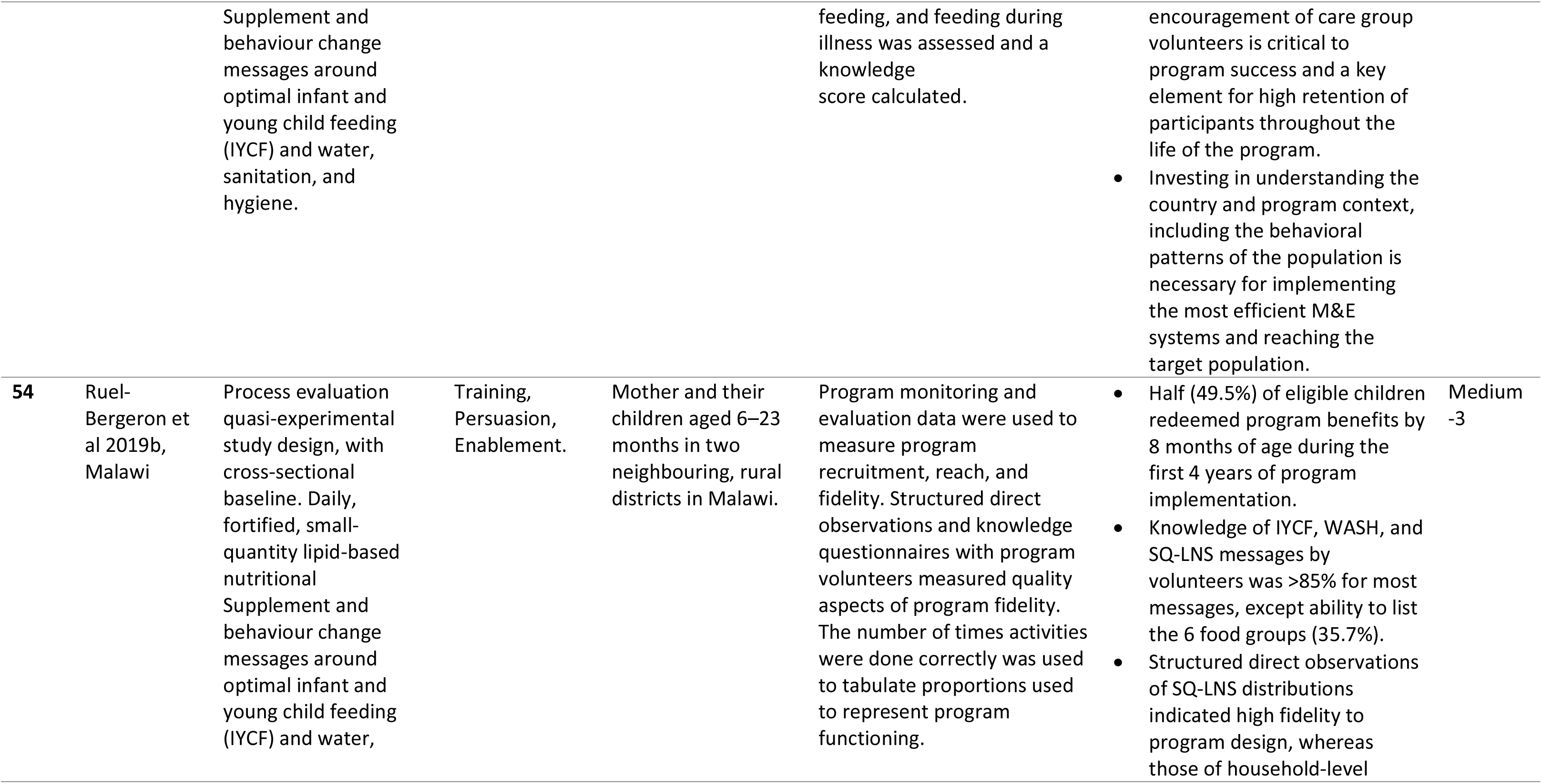
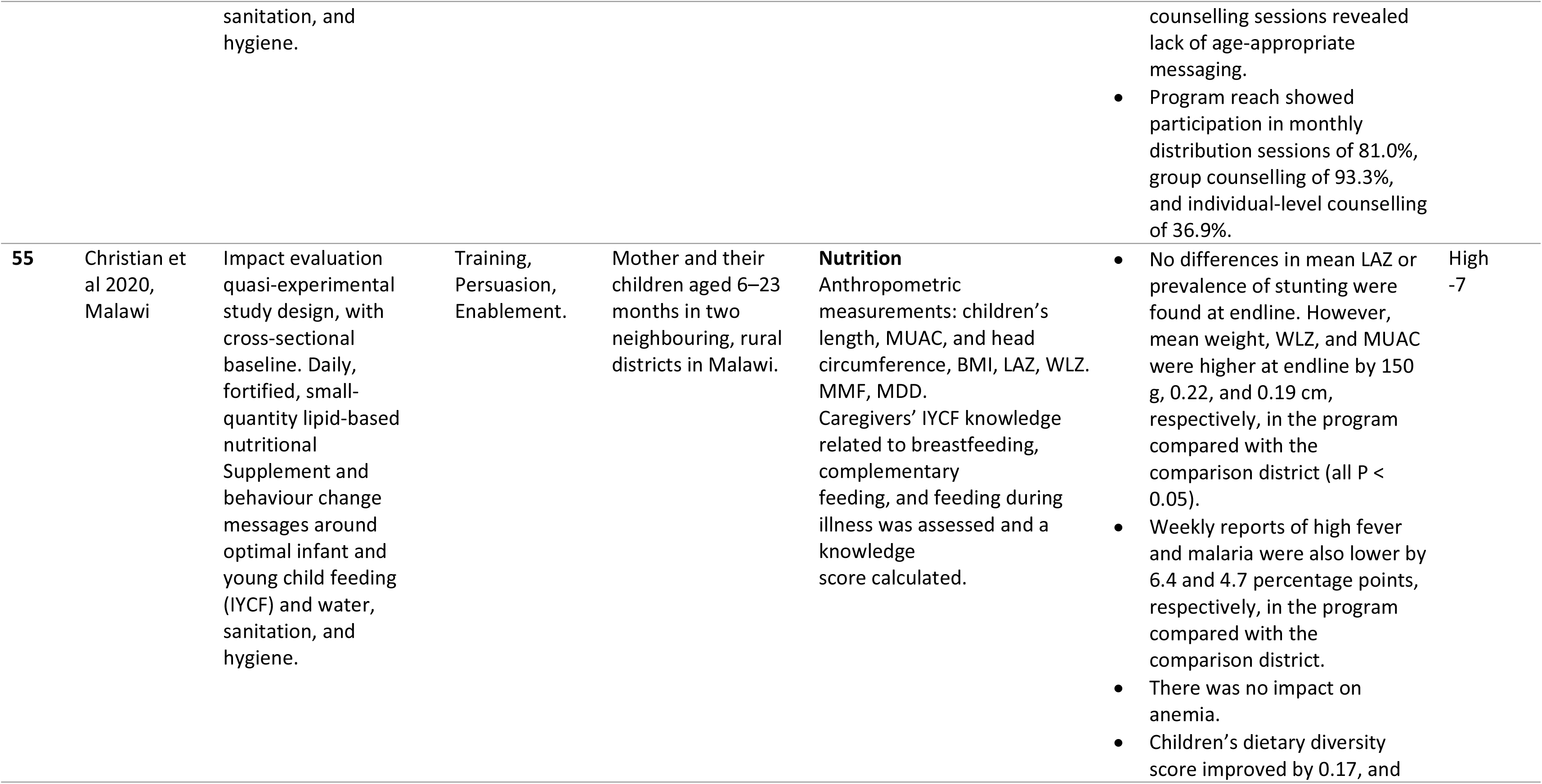
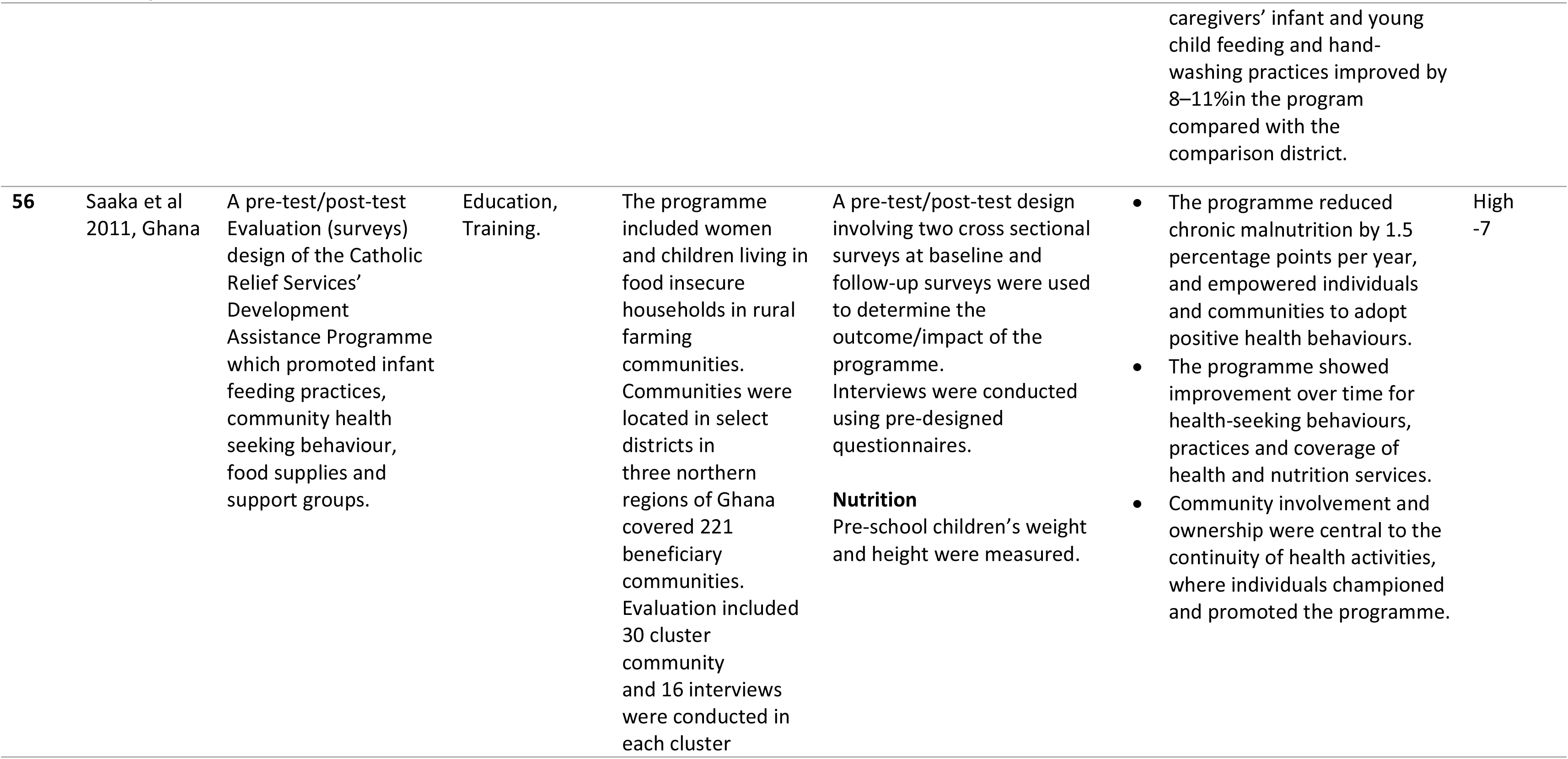
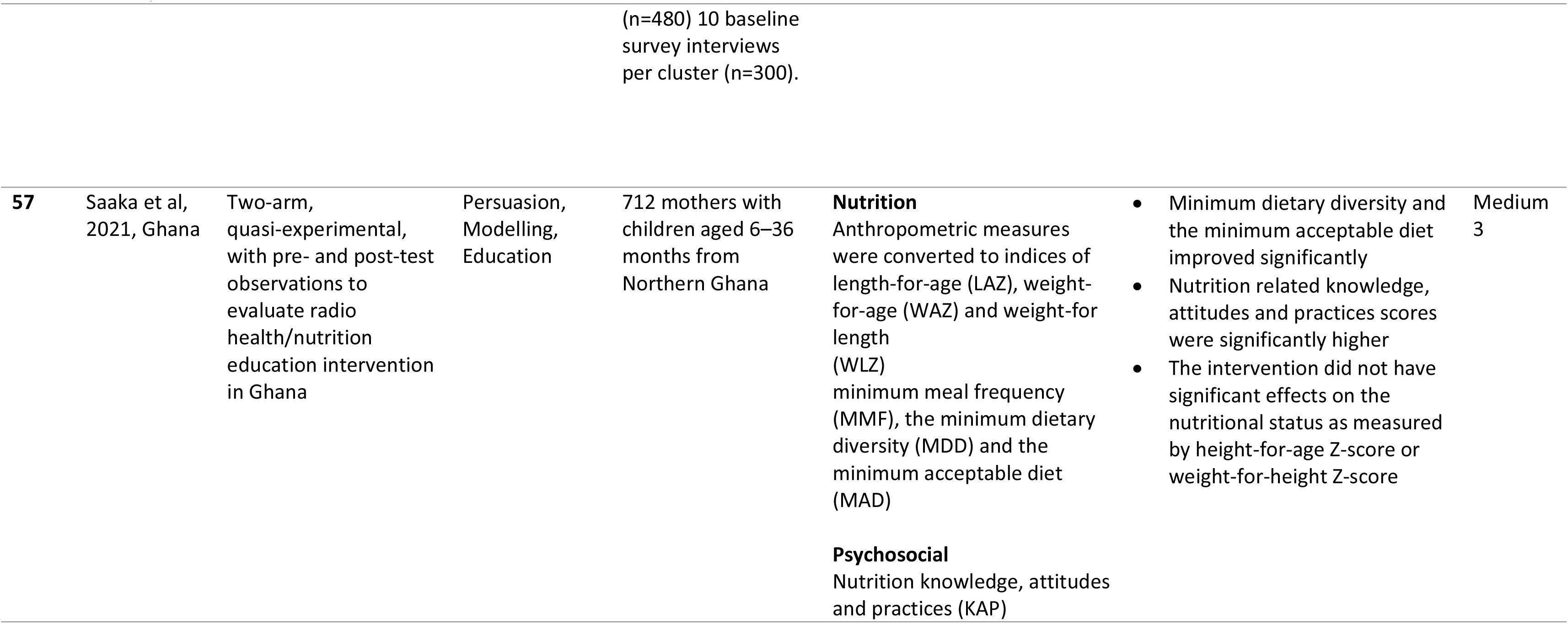
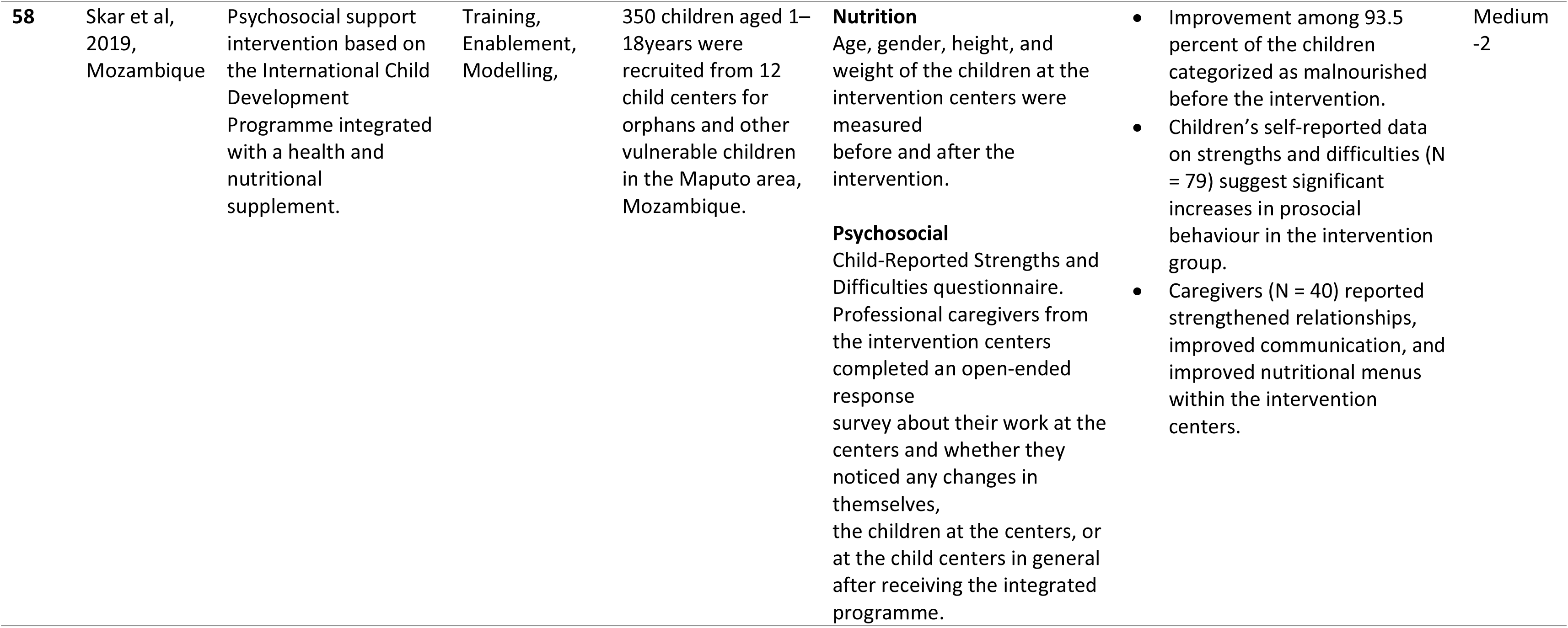
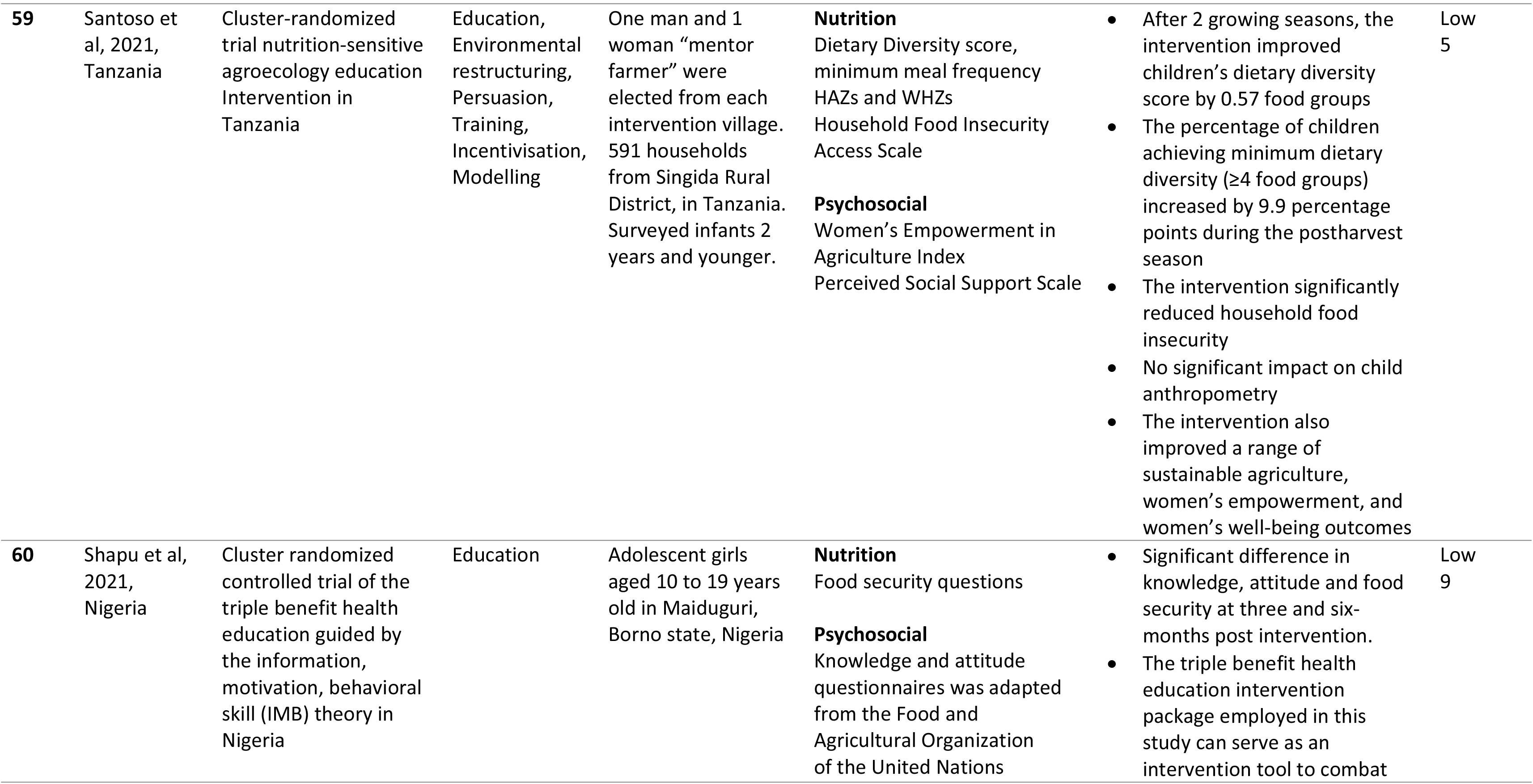
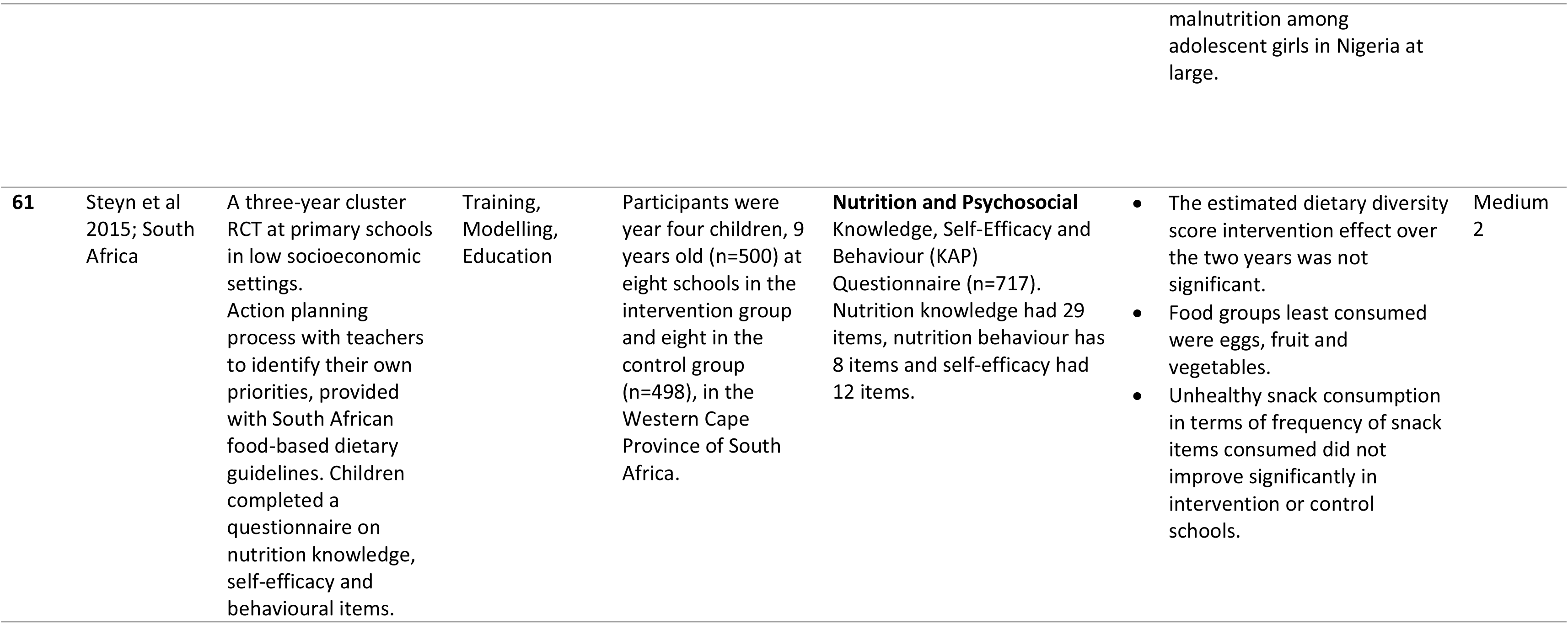
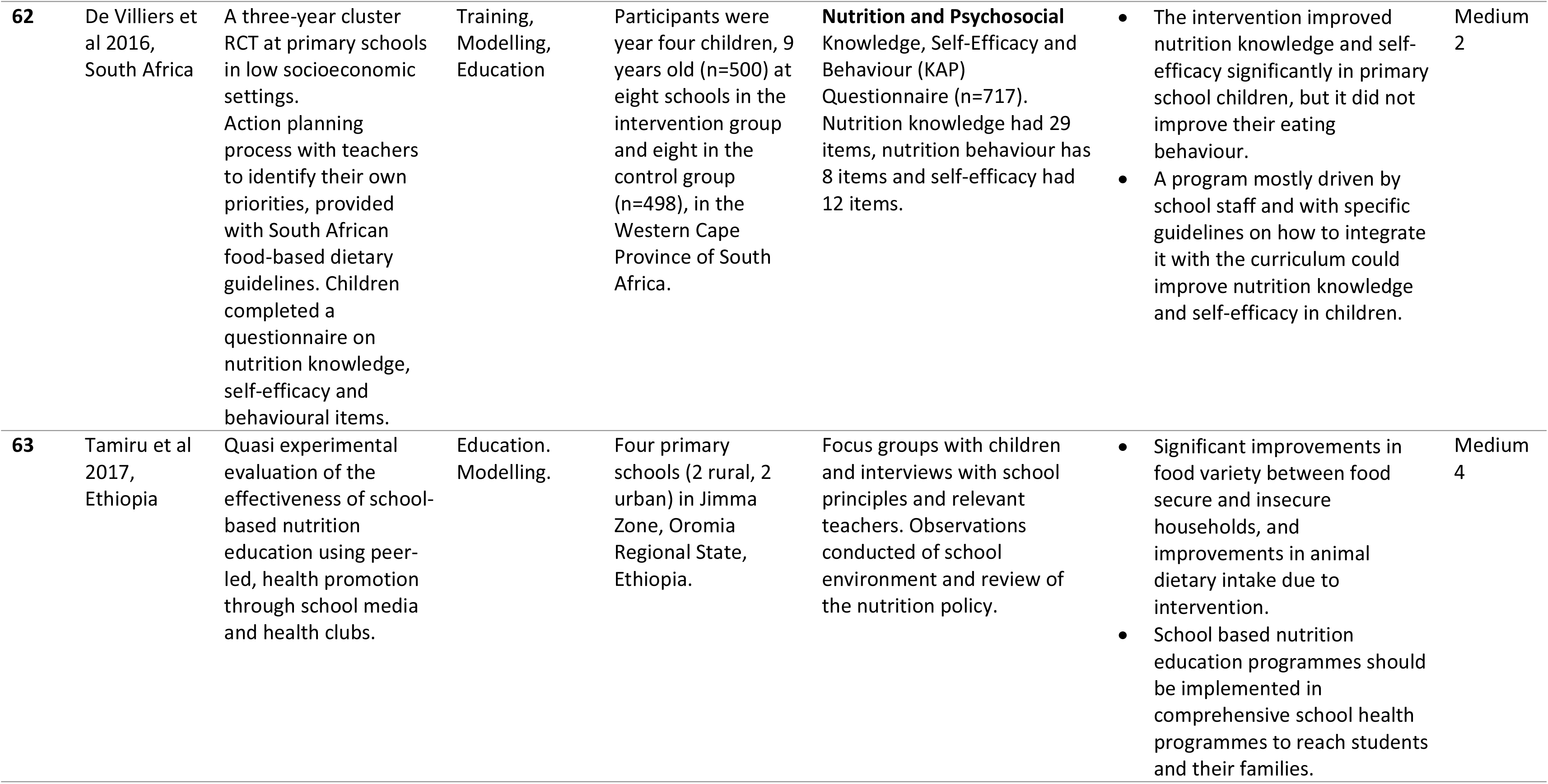
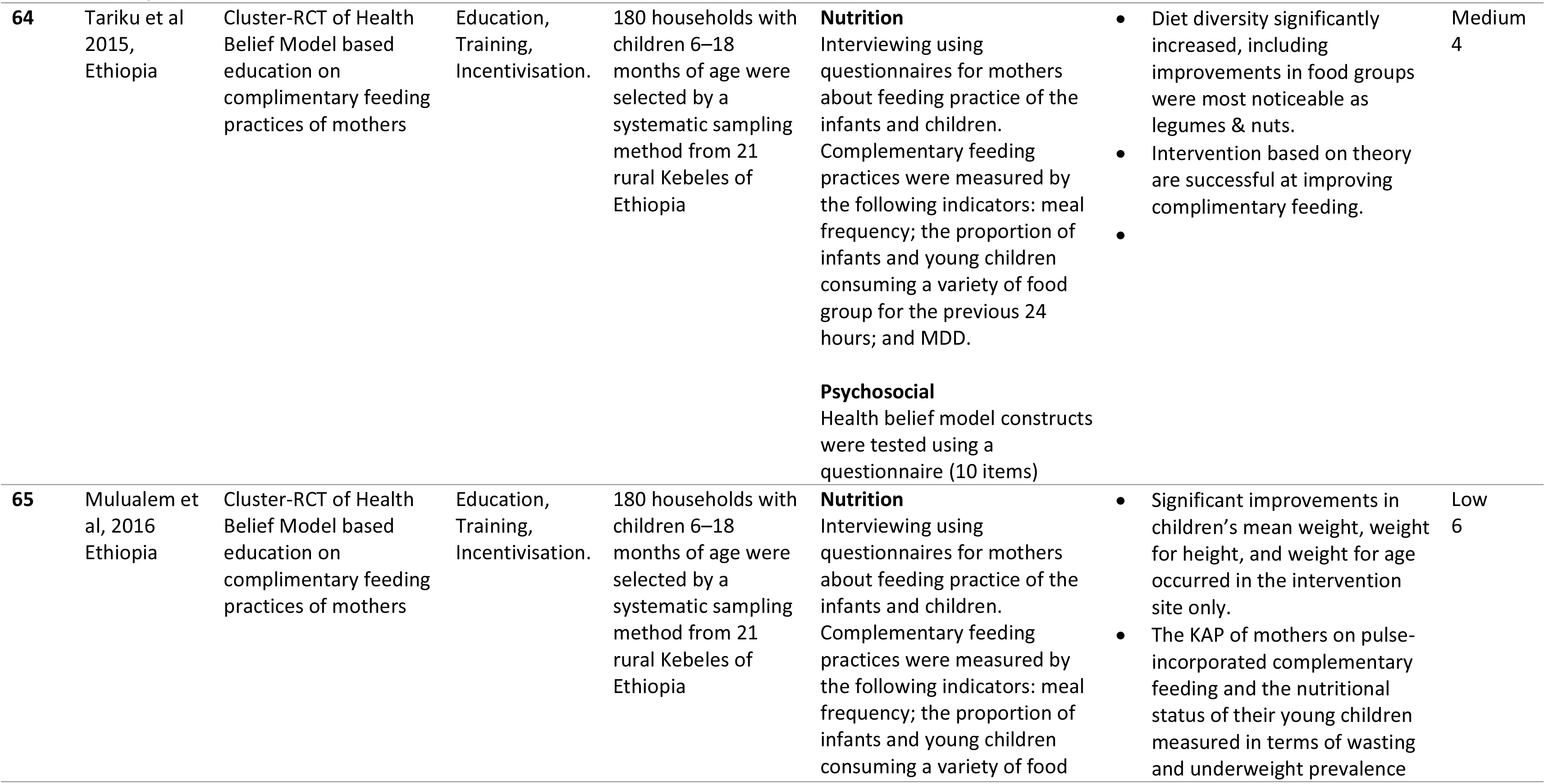
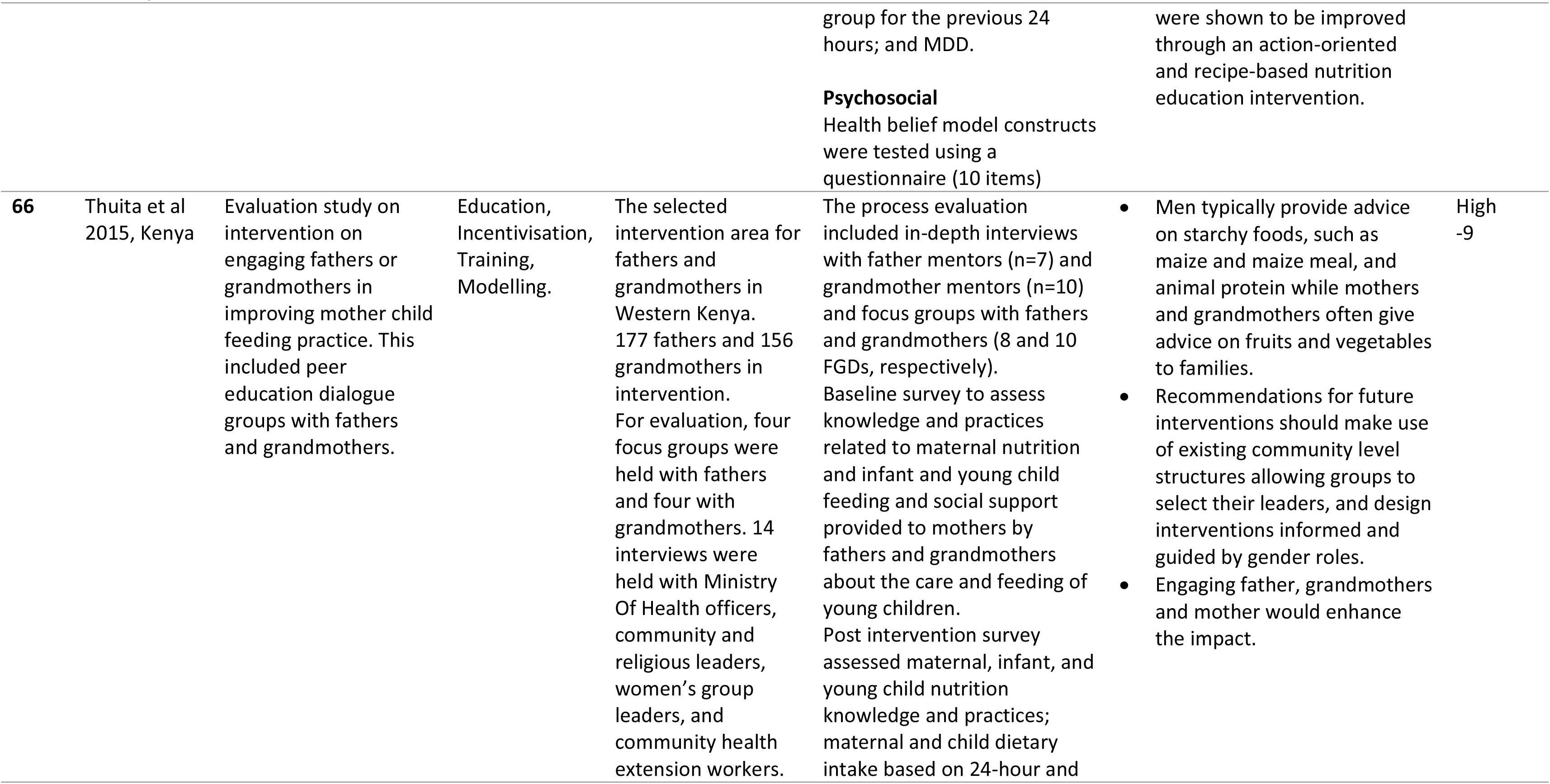
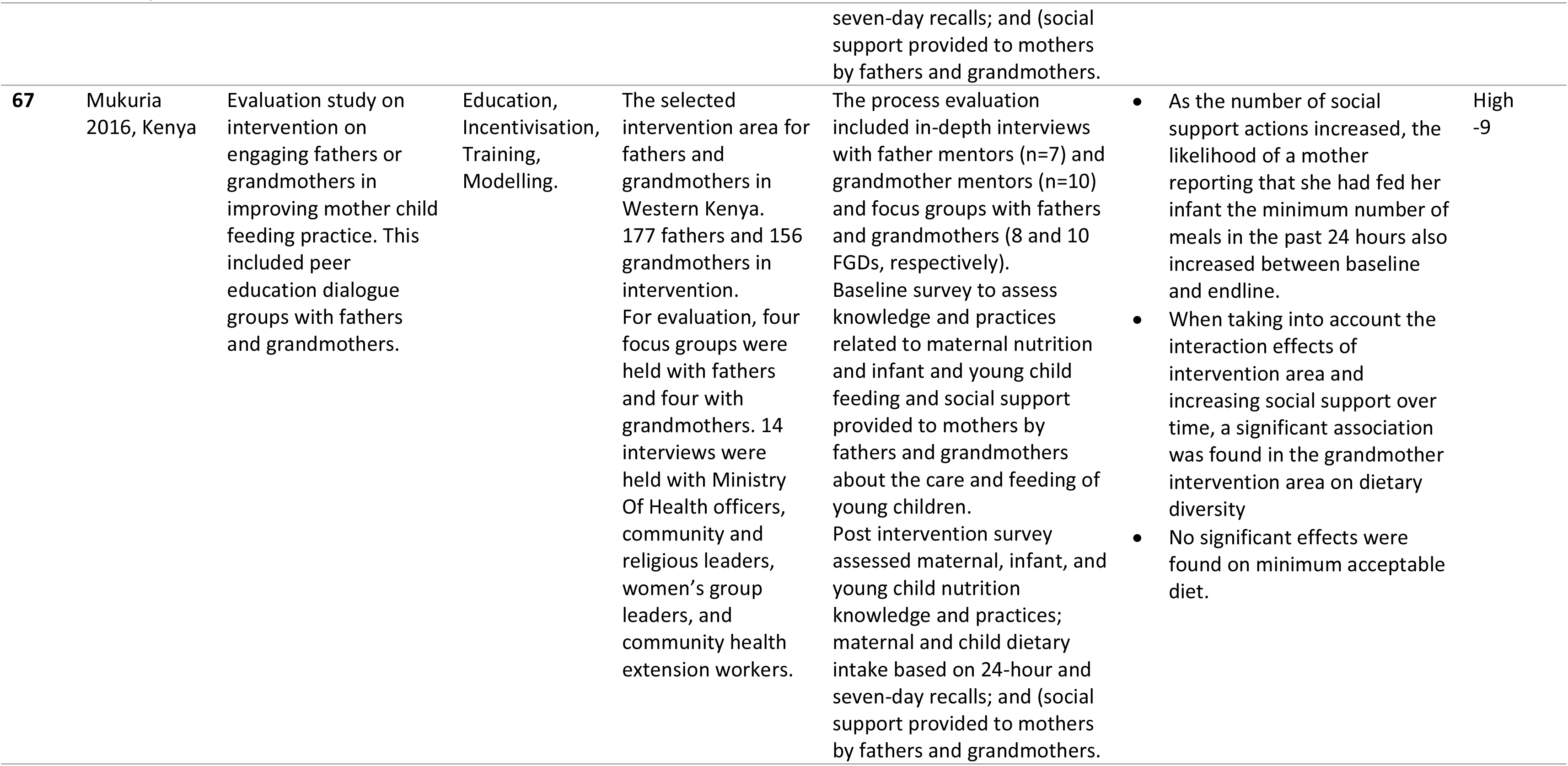
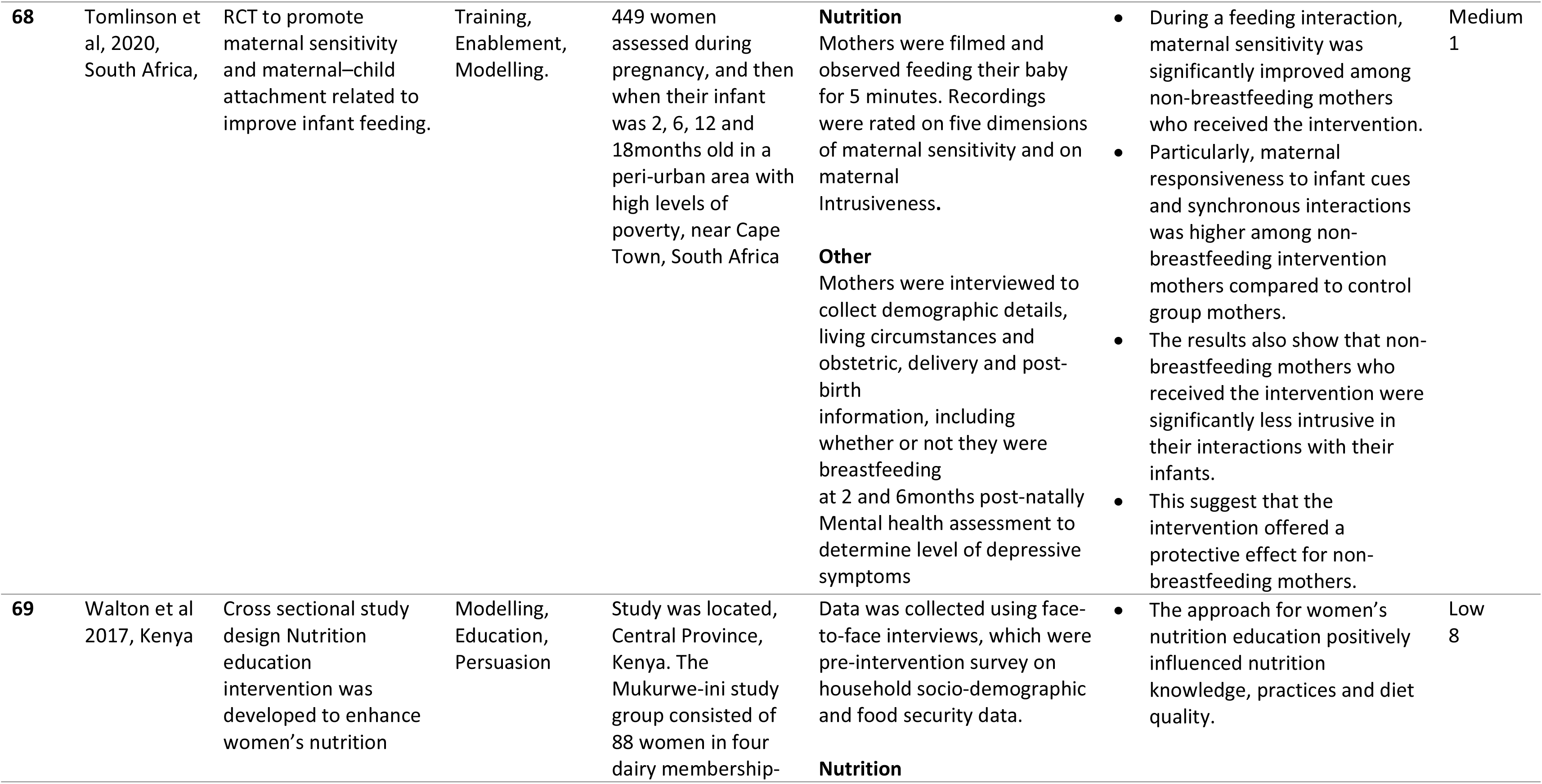
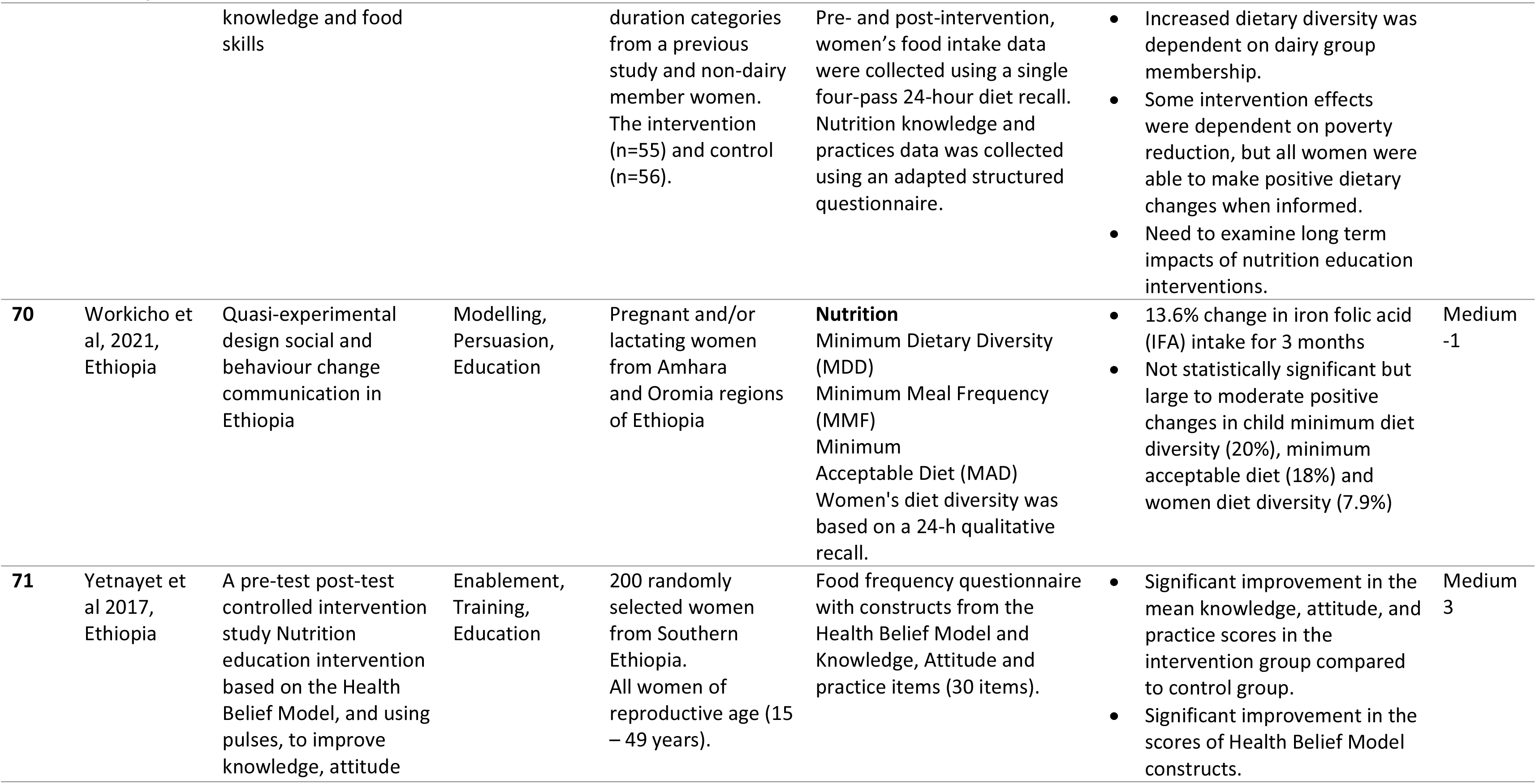
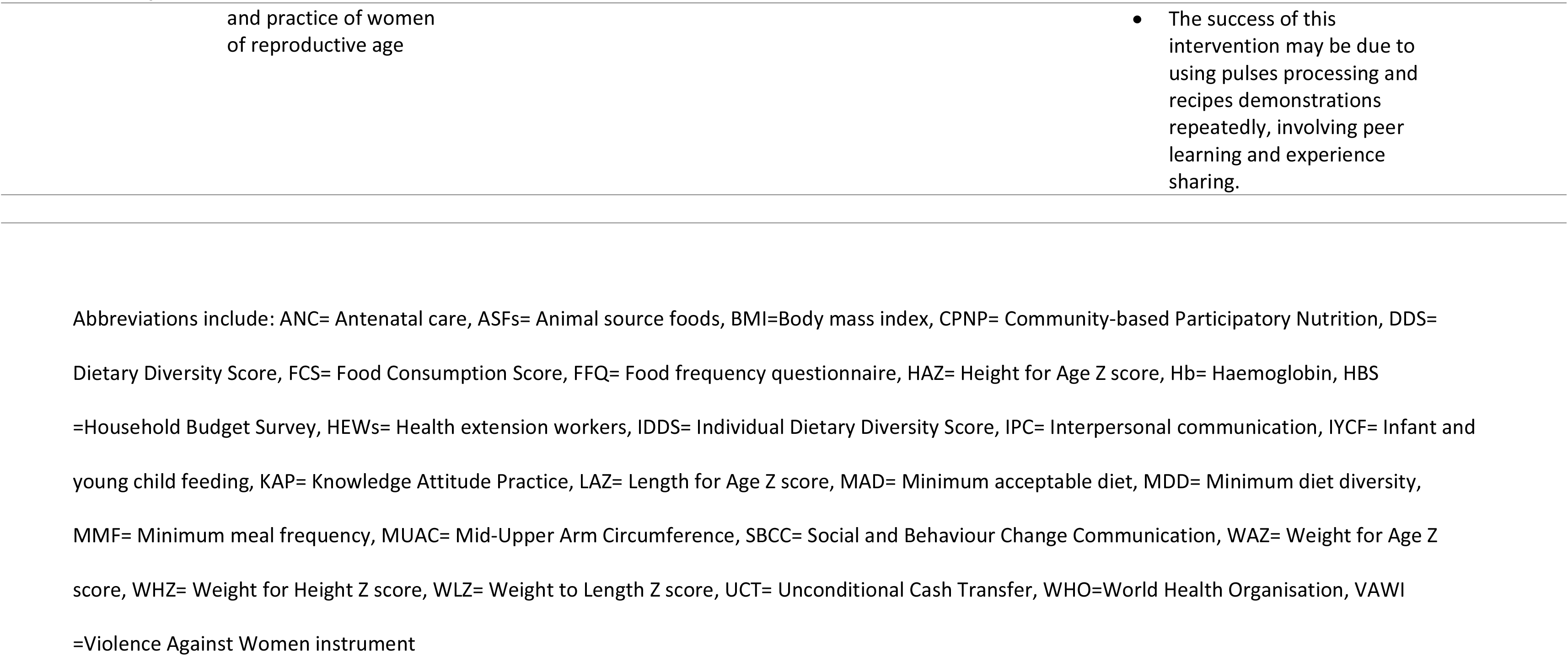
Descriptions of included studies found in the systematic review

### Anthropometric markers

A mother and child lipid supplementation RCT, graded as medium quality, combined with maternal nutrition counselling in Madagascar reported an increase in infants’ length-for-age z-scores (LAZ) of 0.210–0.216SD and stunting reduced by 8.2–9.0% compared to the control^(20, 75)^. Two similar medium-low-quality interventions in Malawi found mixed results, where one intervention had no effect on infant LAZ scores or stunting prevalence, but both interventions found significantly higher weight-for-length z-score (WLZ) and mid-upper arm circumference at 18 months to three years follow-up^(77, 78, 82, 85)^. These results have to be viewed with caution, however, since the trial was low quality being not randomised or blinded. A high quality intervention that used Theory of Planned Behaviour constructs (attitudes, subjective norm, and perceived behavioural control) to predict school children’s behavioural intention to eat pulses in Ethiopia reduced wasting prevalence from 17% to 5% (p<0.001) but had no effect on stunting prevalence^(42)^. An agricultural production and behaviour change communication intervention for mothers in Burkina Faso, the study of which was rated high quality, significantly improved child wasting, diarrhoea, haemoglobin levels and rates of anaemia, especially among the youngest, as well as reducing underweight prevalence^(54, 56, 109)^. Two other high-quality studies of interventions that provided food rations or vouchers and behaviour change communication to mothers in Burundi and Ethiopia, showed reduced prevalence of child stunting in the intervention compared to the control arm^(48–52, 70, 111)^. Behaviour change communication and unconditional cash transfers intervention which was deemed medium quality, to mothers in Togo reduced low birthweight (<2,500g) and protected infant height-for-age z-scores (HAZ) in the intervention arm compared to HAZ decline in the control arm, but had no impact on stunting^(61)^. One chicken production intervention, rated high quality, improved infant HAZ and weight-for-age z-scores (WAZ) both with and without behaviour change communication in Ethiopia^(64)^. A RCT, graded high quality, that provided families with eggs and behaviour change communication significantly decreased the prevalence of infant wasting and underweight^(74)^. An agricultural (tools and agricultural extension workers), behaviour change communication and nutrition counselling intervention, graded high quality, in Ethiopia reduced prevalence of stunting (36.3% to 22.8%) in children aged 6–23 months^(67, 68, 76)^. A psychosocial intervention that promoted infant feeding practices improved food supplies and engaged religious support groups in Ghana reduced HAZ by 1.5% / year^(91)^. This intervention was deemed low quality as it was not an RCT and also lacked quality behaviour change measures. In Ethiopia, a nutrition counselling intervention, graded high quality and based on Social Cognitive Theory and Theory of Planned Behaviour, led to a 17% lower prevalence of undernutrition among pregnant women compared with the control^(62)^. A high quality RCT based on behaviour change communication and agricultural support in Malawi had no significant impact on pre-schooler growth, but reduced stunting among younger siblings^(44)^. A medium rated quality education intervention in Ghana based on Social Cognitive Theory found marginal difference in HAZ and WAZ between school children in each treatment arm^(84)^. One RCT, graded high-quality, that trained local Tanzanian women and farmers to deliver agricultural education to households as well as them receiving seedlings did not find significantly improved infant HAZs and WHZs scores^(72)^. Two lipid supplementation interventions, graded as medium-high quality, which also included behaviour change communication had no significant impact on the prevalence of infant undernutrition, weight for length, or mid-upper arm circumference in Burkina Faso^(60)^ or Mali^(63)^, but in Mali it was found that children in the intervention arm were 29% less likely to experience a first acute malnutrition incident.

### Dietary outcomes

A behaviour change communication intervention, graded as high-quality, among pre-schoolers in Malawi increased intake of energy, protein and micronutrients^(44)^. One RCT of an intervention providing behaviour change communication and food rations, rated as high quality, increased energy consumption in pregnant women and mothers from Burundi^(48, 49, 51, 52, 111)^. Two interventions, graded as medium-high quality and based on nutrition education, found no significant changes in the nutrient intake of children in Malawi and South Africa^(44, 88, 89)^. Yet one RCT rated as medium-quality found significant infant dietary diversity improvement with a combined nutrition education and counselling approach in Malawi^(71)^. Three low quality lipid supplementation studies with behaviour change communication found no impact on maternal haemoglobin concentration and anaemia in Malawi^(77, 78, 82)^. These interventions were rated low quality due to the lack of RCT methods including randomisation and blinding. Yet another similar intervention graded as medium quality in Malawi found significant increases in dietary diversity of infants at 18 months follow up^(85)^. Two low-medium rated quality psychosocial interventions increased pregnant women’s and mothers’ dietary diversity, and household food consumption in the Central African Republic^(107)^ and Ethiopia^(79–81)^. One intervention underpinned by Social Cognitive Theory and the Theory of Planned Behaviour, graded high-quality, had school children in South Africa meeting 5-a-day fruit and vegetable guidelines^(45, 46)^. Two behaviour change communication interventions, graded high quality, increased maternal dietary diversity, and the proportion of children consuming >4 food groups in Burundi^(48, 49, 51, 52, 111)^, and egg consumption in Ethiopia^(79–81)^. One behaviour change communication RCT, deemed medium-quality, found increased infant consumption of milk, but this was not significant^(69)^. Total household food consumption increased for communities in Burkina Faso^(54, 56, 109)^, for pre-schoolers and infants in Malawi^(44, 77, 78, 82)^ and Ethiopia^(59, 67, 68, 76)^, and mother and daughter pairs in Nigeria^(97)^. These interventions were deemed medium to high quality studies due to their rigorous RCT methods. A intervention graded medium-quality that delivered radio dramas on maternal nutrition in Ghana found improved infant dietary diversity^(86)^. One RCT, deemed high-quality, that trained local Tanzanian women and farmers to deliver agricultural education to households as well as them receiving seedlings found significantly improved infant dietary diversity and food insecurity^(72)^. A intervention graded medium quality that delivered behaviour change communication and unconditional cash transfers to mothers in Togo improved household food security^(61)^. Interventions graded medium quality that included nutrition education and cooking demonstrations increased diet diversity scores of women and children in Ethiopia^(42)^ and South Africa^(88, 89)^. These interventions were deemed medium quality due to the self-reported measures. Information and education interventions with medium to low quality had no effect on dietary diversity^(84, 96)^ and strategies to increase self-efficacy had no effect on diet intake in primary school children^(40, 41, 87)^.

### Psychosocial outcomes

Interventions aimed to address psychosocial outcomes including nutrition knowledge (n=17)^(40–42, 44–46, 58, 61, 71, 73, 84, 90, 94, 96, 97, 103, 104)^, practices (e.g. food/recipes preparation, healthy eating)(n=5)^(42, 90, 94, 96, 98)^, attitudes (n=6)^(42, 45, 46, 62, 90, 94)^, intentions (n=4)^(45, 46, 62, 99)^ and confidence in eating healthy food (n=1)^(104)^. These were variously underpinned by behaviour change theory(n=10)^(42, 45, 46, 58, 62, 84, 90, 94, 96, 98)^, strategies to increase self-efficacy (n=5)^(40–42, 87, 99)^, behaviour change communication (n=2)^(44, 61)^ or behaviour change techniques (n=1)^(97)^. In two interventions rated high quality and one rated low quality, women’s capability to feed themselves and their children more nutritious food increased, including interventions based on behaviour change communication in Burkina Faso^(54, 56, 109)^ and Zambia^(83, 110)^, and a psychosocial intervention in Ghana^(91)^. Five medium-high rated quality psychosocial interventions achieved positive psychosocial outcomes, which improved caregiver feeding behaviours. This included increasing women’s psychological well-being in the Central African Republic^(107)^ and Uganda^(53)^. Caregivers of children with cerebral palsy in Tanzania improved their feeding skills^(70)^. Caregiver relationships and communication improved leading to children’s prosocial behaviour in Mozambique^(65)^. Mother’s expression of love and responsiveness during feeding interactions increased in South Africa^(66)^. An intervention graded medium-quality which delivered radio dramas on maternal nutrition in Ghana found increased mother’s knowledge of feeding practices^(86)^. Qualitative evaluations identified barriers to the success of behavioural interventions including availability of, and access to, nutritious and affordable foods and less time for mothers to care for children^(106)^.

### Behavioural intervention components

Interventions were coded on the basis of involving one or more Behaviour Change Wheel intervention functions (**Error! Reference source not found.**)^(28)^. Functions (Appendix C) included education(n=51)^(20, 39–41, 44–47, 49, 51–54, 56, 58, 69–75, 79–81, 83–89, 91, 92, 95, 97–102, 105–109, 111)^, enablement(n=50)^(20, 39, 44–47, 49, 51–54, 56, 59–71, 74–83, 85, 87–90, 93, 94, 97–99, 103–105, 108–111)^, persuasion(n=38)^(39, 49, 51, 52, 54, 56, 60–64, 67–72, 74, 76–78, 82, 83, 86, 96–98, 100, 103, 108–111)^, training(n=36) ^(20, 39–42, 45–47, 54, 56, 59, 61, 65, 66, 69, 70, 72, 74, 75, 77, 78, 82, 84, 87, 90–94, 99, 101–105, 109, 110)^, modelling(n=34)^(40, 41, 45, 46, 54, 56, 59, 65–68, 70–72, 74, 76, 86, 95, 100–102, 109)^, environmental restructuring (n=14)^(42, 44, 47, 54, 56, 64, 67, 68, 72, 76, 90, 106, 109, 110)^, incentivisation (n=11)^(61, 63, 70, 92, 93, 97, 101, 106, 107, 112)^. There were no interventions that included restriction or coercion. Table 4 indicates that in interventions where the Behaviour Change Wheel intervention functions persuasion, environmental restructuring and incentivisation were used, body composition and diet intake improved (BCW = Behaviour Change Wheel; Heat map to indicate the percentage of interventions where outcomes improved or did not improve. Key: Darker green and red indicate higher percentage of interventions per outcome). Education, enablement and training appeared to have less impact on these maternal and infant body composition and diet intake outcomes. However, psychosocial outcomes seemed to improve in interventions with training, incentivisation, modelling and education functions, and less so in interventions which included enablement such as nutrient supplementation. Interventions with multiple Behaviour Change Wheel intervention functions tended also to be those that improved maternal and child nutrition and health outcomes. The optimal number of intervention functions is not possible to determine because so few of the interventions had more than three. Our ability to draw conclusions about the effectiveness of intervention functions is limited by the fact that many of the studies do not measure all outcomes; measurements of nutritional intake are often omitted.

**Table 4.** The numbers of interventions using Behaviour Change Wheel intervention functions and the percentage of those where outcomes improved, did not improve or were not measured.

| Behaviour Change Wheel Intervention Function (BCW IF) | Number of interventions including each function | Anthropometric markers |  |  | Dietary outcomes |  |  | Psychosocial outcomes |  |  |
| --- | --- | --- | --- | --- | --- | --- | --- | --- | --- | --- |
|  |  | Significant | Non-Significant | Not measured | Significant | Non-Significant | Not measured | Significant | Non-Significant | Not measured |
| Enablement | 50 | 30.0% | 8.0% | 64.0% | 50.0% | 12.0% | 40.0% | 54.0% | 6.0% | 42.0% |
| Education | 51 | 19.6% | 11.8% | 68.6% | 52.9% | 13.7% | 33.3% | 76.5% | 3.9% | 19.6% |
| Training | 36 | 27.8% | 11.1% | 61.1% | 41.7% | 16.7% | 41.7% | 83.3% | 5.6% | 11.1% |
| Persuasion | 38 | 34.2% | 15.8% | 52.6% | 57.9% | 10.5% | 34.2% | 52.6% | 5.3% | 44.7% |
| Modelling | 34 | 23.5% | 14.7% | 64.7% | 64.7% | 11.8% | 26.5% | 70.6% | 0.0% | 32.4% |
| Environmental reconstructing | 14 | 35.7% | 21.4% | 50.0% | 71.4% | 0.0% | 35.7% | 71.4% | 0.0% | 35.7% |
| Incentivisation | 11 | 36.4% | 9.1% | 54.5% | 72.7% | 0.0% | 27.3% | 90.9% | 0.0% | 9.1% |
| 1 BCW IF | 5 | 20.0% | 0.0% | 80.0% | 60.0% | 20.0% | 20.0% | 40.0% | 20.0% | 40.0% |
| 2 BCW IF | 11 | 18.2% | 18.2% | 63.6% | 54.5% | 9.1% | 36.4% | 54.5% | 0.0% | 45.5% |
| 3 BCW IF | 32 | 25.0% | 9.4% | 65.6% | 34.4% | 18.8% | 46.9% | 65.6% | 0.0% | 34.4% |
| 4 BCW IF | 14 | 21.4% | 7.1% | 78.6% | 71.4% | 14.3% | 21.4% | 57.1% | 14.3% | 35.7% |
| 5 BCW IF | 6 | 50.0% | 16.7% | 33.3% | 83.3% | 0.0% | 16.7% | 100.0% | 0.0% | 0.0% |
| 6 BCW IF | 3 | 33.3% | 33.3% | 33.3% | 100.0% | 0.0% | 0.0% | 66.7% | 0.0% | 33.3% |

Table 5 describes the way in which people’s capability, opportunity and motivation are addressed in interventions to improve maternal and child nutrition and health outcomes. For example, food demonstrations and provision of agricultural land address physical capability and opportunity components respectively as specified by the COM-B model as necessary to change behaviour.

**Table 5.**
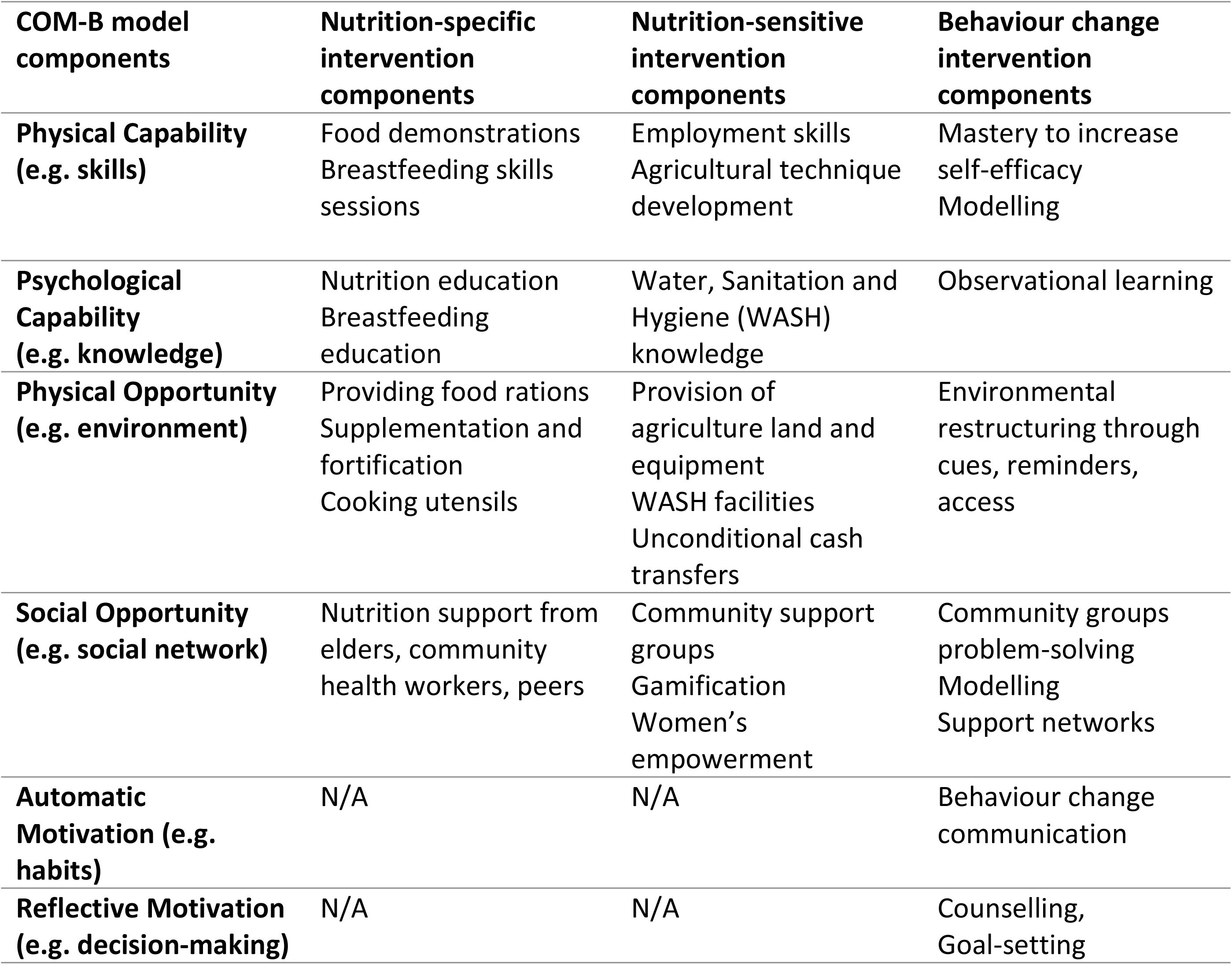
The categories of studies’ intervention components analysed to the COM-B model

| **COM-B model components** | **Nutrition-specific intervention components** | **Nutrition-sensitive intervention components** | **Behaviour change intervention components** |
| --- | --- | --- | --- |
| **Physical Capability (e.g. skills)** | Food demonstrations<br>Breastfeeding skills sessions | Employment skills<br>Agricultural technique development | Mastery to increase self-efficacy<br>Modelling |
| **Psychological Capability (e.g. knowledge)** | Nutrition education<br>Breastfeeding education | Water, Sanitation and Hygiene (WASH) knowledge | Observational learning |
| **Physical Opportunity (e.g. environment)** | Providing food rations<br>Supplementation and fortification<br>Cooking utensils | Provision of agriculture land and equipment<br>WASH facilities<br>Unconditional cash transfers | Environmental restructuring through cues, reminders, access |
| **Social Opportunity (e.g. social network)** | Nutrition support from elders, community health workers, peers | Community support groups<br>Gamification<br>Women's empowerment | Community groups problem-solving<br>Modelling<br>Support networks |
| **Automatic Motivation (e.g. habits)** | N/A | N/A | Behaviour change communication |
| **Reflective Motivation (e.g. decision-making)** | N/A | N/A | Counselling, Goal-setting |

## Discussion

This review of 71 studies addressed the question of whether nutrition-specific and nutrition-sensitive interventions were more effective in improving maternal and child nutrition outcomes in sub-Saharan Africa if they included behaviour change functions. It was found that those that were based on behaviour change theory, counselling and communication, produced the most significant positive impacts on infant and children wasting, underweight and stunting, and improved diet outcomes including dietary diversity and total food consumption. There is very little evidence that these interventions improved maternal and infant nutrient intake, which could be because changes in nutrient intake were not often measured due to a lack of comprehensive data on the nutrient composition of foods. These interventions reviewed also identified improvements in maternal psychosocial outcomes in relation to nutrition, including knowledge, practice, attitude, intention, confidence, capability and wellbeing.

Interventions which included the Behaviour Change Wheel intervention functions, i.e. incentivisation, persuasion and environmental restructuring, tended to be those that improved maternal and child nutrition and psychosocial outcomes. The COM-B model suggests that omitting motivational components reduces the effectiveness of behaviour change interventions. This analysis in this table 5 demonstrates that nutrition-specific and nutrition-sensitive interventions cannot alone address motivational components and require additional behaviour change functions in order to improve maternal and child nutrition and health outcomes.

### Strengths and limitations

A strength of this systematic review is the inclusion of a wide variety of studies using a range of intervention methods and outcomes to provide a broad overview of the way in which behavioural functions may increase effectiveness of nutrition interventions to improve maternal and child nutrition outcomes in sub-Saharan Africa. One limitation was that breastfeeding interventions were excluded. This was due to previous systematic reviews finding limited evidence of behaviour change components in the interventions^(33, 34)^. One weakness of the review is that mainly papers written and published in English were reviewed. Another is that not all authors of these papers explicitly identified behaviour change functions in their descriptions of interventions, meaning that we may have missed other relevant papers; all titles, abstracts and full texts were double-screened against the inclusion and exclusion criteria, however, in order to minimise this possibility. Consistency in the types of data extracted and in judgements of the quality of the studies was ensured by having two independent reviewers review the included studies.

### Implications

Improving nutritional outcomes requires behaviour change at some level. To change nutrition behaviour, the COM-B model suggests that capability, opportunity and motivation should all be addressed^(28)^. Our analysis demonstrates that most nutrition-specific and nutrition-sensitive interventions only address participants’ capability and opportunities and require the addition of behaviour change functions such as behaviour change communication and counselling to address missing motivational components. For example, one trial in Burkina Faso gave women land and seedlings (physical opportunity) and also provided volunteers to facilitate women’s groups using behaviour change communication that was empowering and motivating; this package of interventions led to improvements in maternal and infant body composition, diet intake and psychosocial outcomes which have potential for long-lasting nutritional benefits for women and their families^(54–56)^. Research would suggest that these behaviour change communications and women’s groups may have engaged with both automatic and reflective motivation^(28)^. Automatic motivation is defined as the unconscious process that “energizes and directs behaviour” ^(28)^(p4), including habits and emotional responses. Reflective motivation describes conscious processes such as analytical decision-making. Education functions of the communications in the trial in Burkina Faso may have changed the conscious decisions women were making about how to feed themselves and their children (reflective motivation), and the groups may have created new social norms that encouraged women to change their dietary habits (automatic motivation).

This review linked three Behaviour Change Wheel intervention functions to improved outcomes. These were incentivisation, persuasion and environmental restructuring. All are hypothesised to change motivations^(28)^. Providing incentives, which include cash transfers and food vouchers, has been found to improve health outcomes, probably because it increases household disposable income allowing families to spend more money on food^(61, 107)^. This is particularly significant in improving outcomes in the poorest communities. Another incentive used was nutritional supplements, which increased intervention attendance and uptake^(63)^. In contrast, failing to incentivise community health workers was given as a reason for their lack of motivation for delivering interventions^(60)^. Overall, there is compelling evidence that incentives can be a powerful motivating force in behaviour change interventions^(113)^.

Persuasive communications, which are designed to induce positive or negative feelings or stimulate action, were a common feature of many of the interventions in this review. These tended to focus on improving diet quality and diversity and were delivered through singing groups, storytelling and women’s groups. The Behaviour Change Wheel indicates that persuasion, because it is often emotive, engages both reflective and automatic motivations^(28)^. This distinguishes it from purely educational communications which work by informing conscious decision-making (reflective motivation). In one intervention that included persuasive communication, grandmothers were encouraged through nutrition education to promote improved nutritional practices related to pregnancy and infant feeding within their families^(103)^. This was delivered through songs, storytelling and group discussions and made an emotional appeal to grandmothers’ intrinsic commitment to family well-being. These persuasive communications were therefore explicitly designed to induce an emotional response (automatic motivation) as well as educate and inform (reflective motivation).

Environmental restructuring is the process of changing the physical and social environment, which is hypothesised to indirectly change motivation by cuing behaviour in a way which is often unconscious (automatic motivation). In the context of this review, environmental restructuring referred to provision of land, seedlings, livestock and equipment with the intention of increasing the effectiveness of community food production for both consumption and sale^(54–56, 64, 106)^. It can be speculated that this provision addresses automatic motivations by increasing availability and diversity of food in these communities and supporting creation of new eating habits intended to improve diet quality.

Figure 4 illustrates the way in which all these intervention functions can be drawn together. Encouragingly, the addition of behaviour change functions to nutrition interventions may provide good value for money; economic analysis of the relative cost of behaviour change functions within one nutrition intervention in Burundi indicated that they accounted for approximately 13% of the budget whereas provision of food rations took up 30%^(52)^. Longitudinal studies are required to assess the impact of nutritional outcomes across children’s lifecourse and as communities undergo economic, nutritional, and societal transition^(114)^.

**Figure 4.**
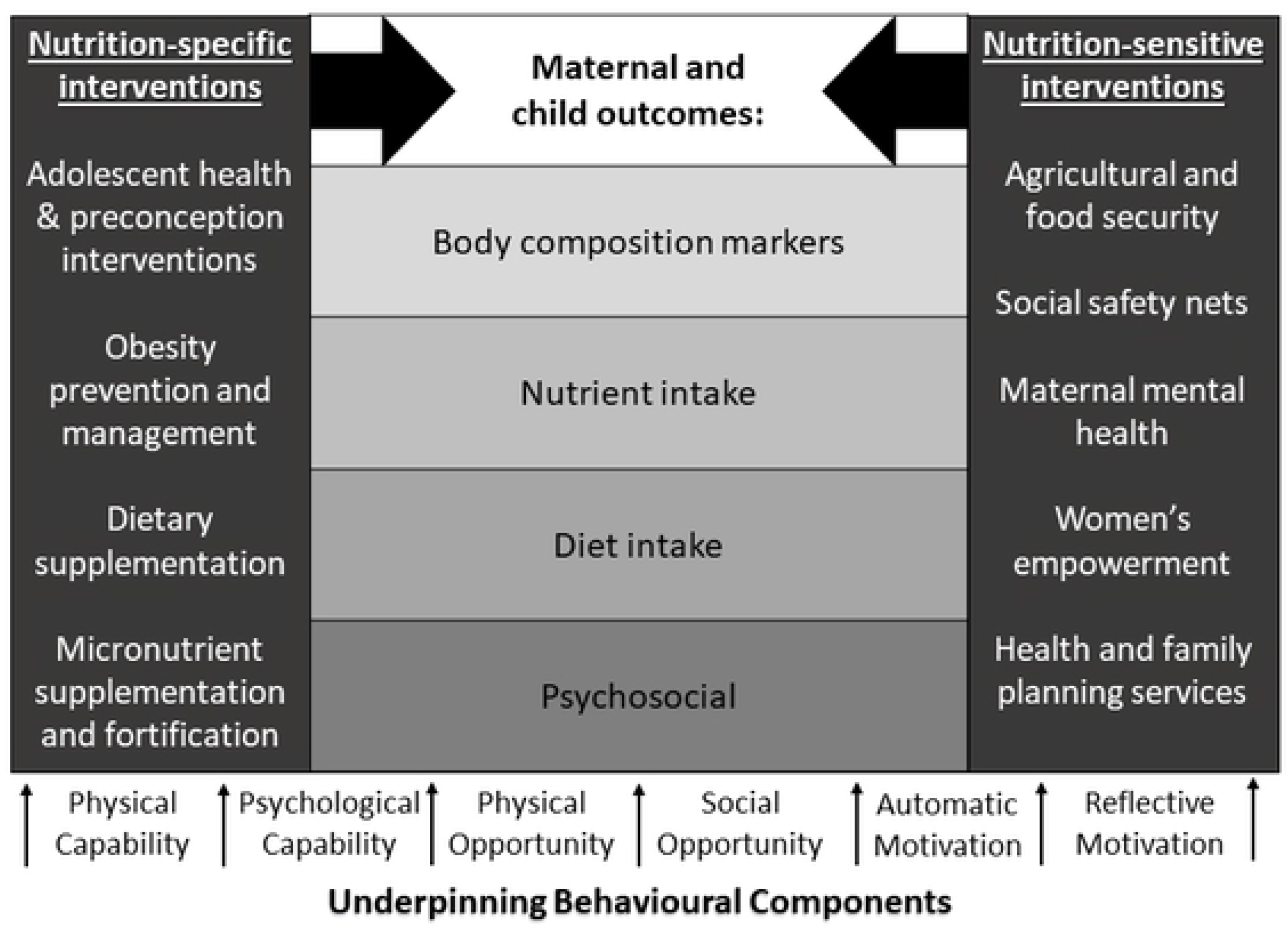
Framework describing the way in which nutrition-specific and nutrition-sensitive interventions may be underpinned by behavioural concepts to improve maternal and child nutritional and psychosocial outcomes in sub-Saharan Africa. (Adaption of Lassi et al, 2017 framework^(115)^)

## Conclusion

This review indicates that nutrition-specific interventions, such as increasing access to food^(20, 75)^, and nutrition-sensitive interventions, such as offering cash transfers^(61)^ are not always enough to improve nutritional status of women and children in sub-Saharan Africa. Behaviour change communication and counselling are also unlikely on their own to be effective because they do not address food insecurity or nutritional deficiencies in high poverty contexts. Our findings indicate that interventions comprising all three Behaviour Change Wheel intervention functions (incentives, persuasion and environmental restructuring) were most likely to be effective in improving nutritional and psychosocial outcomes. By taking a health psychology perspective on this review, specifically drawing on concepts of automatic and reflective motivation from the Behaviour Change Wheel and COM-B model, motivation needs to be considered in designing an intervention to improve nutritional behaviour in the context of sub-Saharan Africa have been identified. To enhance the designs of these interventions, and ultimately improve the nutritional and psychosocial outcomes for mothers and infants in this region, multidisciplinary collaborations are required. Our recommendation would be to task behaviour change and nutrition experts such as health psychologists and nutritionists, as well as intervention designers, policy makers, and commissioners of services to fund and roll out these multicomponent behaviour change interventions.

## Data Availability

Secondary data available on request

## Acknowledgements

We would also like to thank the INPreP group for their contributions to this work: Engelbert A. Nonterah, Abraham Oduro, Cornelius Debpuur, James Adoctor, Paul Welaga, Edith Dambayi, Esmond W. Nonterah, Winfred Ofosu, Doreen Ayibisah, Maxwell Dalaba, Samuel Chatio (Navrongo Health Research Centre); Hermann Sorgho, Palwendé R. Boua, Adelaïde Compaoré, Kadija Ouedraogo, Karim Derra, Aminata Welgo, Halidou Tinto (Clinical Research Unit of Nanoro); Karen J. Hofman, Susan Goldstein, Agnes Erzse, Aviva Tugendhaft, Winfreda Mdewa, Ijeoma Edoka (SAMRC Centre for Health Economics and Decision Science, PRICELESS); Mark Hanson, Marie-Louise Newell, Keith M. Godfrey, Caroline Fall, Polly Hardy-Johnson (Faculty of Medicine, University of Southampton); Shane Norris, Emmanuel Cohen, Stephanie Wrottesley (SAMRC Developmental Pathways for Health Research Unit). We would also like to thank Paula Sands at the University of Southampton Health Sciences library for her support in developing the search strategy.

## Disclaimer

None.

## Authorship

DW, WTL, KW, SHK, MB designed the review, DW, PM, PB, TR, SJ screened and assessed the quality of the papers. All authors contributed to analysis of the data and to drafting and editing the article and approved the final manuscript.

## Appendix A Behaviour change systematic review search strategy

### Search strategy

#### Databases

Cochrane database, EMBASE, Medline, web of science, CINAHL, PSYCHinfo, scopus

#### Inclusion criteria

|  |  |
| --- | --- |
| Study Design | All study designs evaluating maternal and child nutrition interventions. |
| Population | At least one of the following populations living in sub-Saharan Africa:<br>- women of child bearing age (preconception)<br>- pregnant women<br>- fathers<br>- infants, children and adolescents |
| Interventions | Nutrition related interventions |
| Comparator | Non behavioural change intervention |
| Outcomes | At least one of the following outcomes for:<br>- Health outcomes, including physical and psychological<br>- Behavioural outcomes to nutrition<br>- Both short and long term outcomes |

#### Search terms

|  |  |
| --- | --- |
| Population | (Sub Sahara* Africa OR subsahara* OR Africa South of the Sahara )<br>OR<br>(Angola OR Benin OR Botswana OR Burkina Faso OR Burundi OR Cameroon OR Cape Verde OR Central Africa OR Central African Republic OR Chad OR Comoros OR Congo OR Cote d'Ivoire OR Democratic Republic Congo OR Djibouti OR Equatorial Guinea OR Eritrea OR |
|  | Ethiopia OR Gabon OR Gambia OR Ghana OR Guinea OR Guinea Bissau OR Kenya OR Lesotho OR Liberia OR Madagascar OR Malawi OR Mali OR Mayotte OR Mozambique OR Namibia OR Niger OR Nigeria OR Rwanda OR Sahel OR Senegal OR Sierra Leone OR Somalia OR South Africa OR South Sudan OR Sudan OR Swaziland OR Tanzania OR Togo OR Uganda OR Zambia OR Zimbabwe )<br>AND<br>(**Matern\*** OR Mother*OR **pregnan\*** OR pre natal OR prenatal OR post natal OR **postnatal** OR **preconception** OR pre conception OR reproduc* OR paternal OR **parental** OR **family planning** OR 1000 days )<br>OR<br>(**adolescen\*** OR youth OR young OR teen* OR student* OR girl* OR boy* OR pupil* OR **pubert\*** )<br>OR<br>(offspring OR **infant\*** OR **child** OR children OR **childhood** OR baby OR babies OR **newborn** OR **new born** OR paediatric* ) |
| Intervention | AND<br>((**Nutrition\*** OR Diet* OR food OR eat* OR nutrient OR feed*))<br>Adj3<br>(Attribution Theory OR Health locus of control OR Self determination theor* OR Risk perception OR self affirmation theory OR Self efficacy OR Stage of change model OR health action process approach OR Health belief model OR Theory of planned behav* OR social cognitive theor* |
|  | OR Transtheoretical Model of Change OR Theory of Planned Behav* OR information Motivation Behav* skill* OR IBM OR Social Learning Theor* OR diffusion of innovation OR community mobilisation OR ecological perspective OR COMB OR COM B OR Capability, Opportunity, Motivation Behav* OR behav* change wheel))<br>OR<br>((**Nutrition\*** OR Diet* OR food OR eat* OR nutrient OR feed*))<br>Adj3<br>(social norm* OR attitude OR perception OR belief OR susceptibility OR autonomy OR health promotion OR public health campaign OR empower* OR resilien*OR goal setting OR implementation intention* OR co*production intervention))<br>OR<br>((**Nutrition\*** OR Diet* OR food OR eat* OR nutrient OR feed*))<br>Adj3<br>(behav* change technique* OR **behav\* change theor\*** OR behav* change OR behav* change intervention OR behav* modification OR community behav* change OR prevent behav* OR behav* change communication OR cultur* change* OR theoretical domain framework ))<br>OR<br>((**Nutrition\*** OR Diet* OR food OR eat* OR nutrient OR feed*))<br>Adj3<br>(**Health Psycholog\*** theor* OR Health Psychology OR critical Psycholog* theor* OR clinical psycholog* theor* OR social Psycholog* theor* OR **cross cultural psycholog\*** OR anthropology OR psychosocial OR biopsychosocial OR international psychology OR social science OR social medicine OR behav* medicine)) |
| Outcome | AND<br>((**Nutrition\*** OR Diet* OR food OR eat* OR nutrient OR feed*))<br>adj3<br>(**Health OR health\*care OR wellbeing OR mental health OR psychological function\* OR cognition** ) )<br>OR<br>((**Nutrition\*** OR Diet* OR food OR eat* OR nutrient OR feed*))<br>adj3<br>(**human capital OR economic\* OR social OR employment OR education** ) )<br>OR<br>((**Nutrition\*** OR Diet* OR food OR eat* OR nutrient OR feed*))<br>adj3<br>(choice OR behav* OR habit* OR pattern OR prefer* OR selection OR portion OR size OR tast* OR binge OR comfort OR crav* OR perception OR perceive* OR belief OR attitude OR control OR norm*OR value* OR fruit* OR vegetable*)) |

## Appendix B Behaviour Change systematic review screening document

|  |  |  |
| --- | --- | --- |
| Study Characteristics | Inclusion criteria: | Exclusion criteria: |
| Type of study | ☐ Intervention implementation (RCTs, cohort, longitudinal etc),<br>☐ intervention evaluation<br>☐ systematic reviews of interventions | ☐ Protocols, commentary articles, conference papers, abstracts<br>☐ Qualitative methodology to explore perceptions around nutrition<br>☐ Surveys to investigate nutrition status |
| Participants | Populations living in sub-Saharan Africa:<br>☐ women of child bearing age (preconception)<br>☐ pregnant women<br>☐ fathers<br>☐ adolescents<br>☐ new-borns<br>☐ infants<br>☐ children | ☐ Populations living outside sub-Saharan Africa<br>☐ Adults above child bearing age<br>☐ Expats living in Africa |
| Types of intervention | ☐ Nutrition interventions with behavioural science component such as education, communication, social support etc<br>☐ Nutrition interventions with psychological components such as personality, cognition, social psychology<br>☐ Individual, family or community based | ☐ Non-nutrition interventions<br>☐ Interventions solely focused on HIV/AIDS<br>☐ malaria or other infectious diseases<br>☐ Breastfeeding<br>☐ Interview/Focus group studies<br>☐ Survey studies |
| Types of outcome measures | At least one of the following outcomes for infants, children and adolescents:<br>☐ Nutrition outcomes<br>☐ Behavioural outcomes<br>☐ Short and long term health outcomes |  |
| INCLUDE ☐ EXCLUDE ☐ |  |  |
| Reason for exclusion |  |  |
| Notes: |  |  |

## Appendix C Behaviour Change Wheel (Michie et al, 2011)

| Interventions | Definition | Examples |
| --- | --- | --- |
| Education | Increasing knowledge or understanding | Providing information to promote healthy eating |
| Persuasion | Using communication to induce positive or negative feelings or stimulate action | Using imagery to motivate increases in physical activity |
| Incentivisation | Creating expectation of reward | Using prize draws to induce attempts to stop smoking |
| Coercion | Creating expectation of punishment or cost | Raising the financial cost to reduce excessive alcohol consumption |
| Training | Imparting skills | Advanced driver training to increase safe driving |
| Restriction | Using rules to reduce the opportunity to engage in the target behaviour (or to increase the target behaviour by reducing the opportunity to engage in competing behaviours) | Prohibiting sales of solvents to people under 18 to reduce use for intoxication |
| Environmental restructuring | Changing the physical or social context | Providing on-screen prompts for GPs to ask about smoking behaviour |
| Modelling | Providing an example for people to aspire to or imitate | Using TV drama scenes involving safe-sex practices to increase condom use |
| Enablement | Increasing means/reducing barriers to increase capability or opportunity | Behavioural support for smoking cessation in sub-Saharan Africa, medication for cognitive deficits, surgery to reduce obesity, prostheses to promote physical activity |

## Appendix D Behaviour change systematic review quality assessment scoring rubric

|  | Item | Risk of bias |  |  |
| --- | --- | --- | --- | --- |
|  |  | Low (+1) | Medium (0) | High (1) |
| 1 | Study design | Randomised controlled trial, Quasi-experimental studies that include a control group | Evaluation of Randomised controlled trial such as process review | Experimental studies that do not use a control group. |
| 2 | Randomisation | Participants or communities are randomised in large clusters. |  | Not randomised, or inadequate randomisation methods such as birth date used. OR randomisation methods not described |
| 3 | Blinding | Participants and analysts were blinded to treatment condition. | Participants or communities were not sufficiently blinded, but analysis of results was blinded. | No blinding, or insufficient blinding methods. |
| 4 | Were groups similar at baseline? | Appropriate statistical tests (chi-square and/or t-tests) used to analyse differences between groups at baseline, and found that there were no significant differences. | Groups are similar, but there are some differences that are judged to be acceptable. | Analyst did not test for differences at baseline, or there were significant differences between groups. |
| 5 | Selection | Participants were randomly selected. | Community based interventions that require health professionals. | Volunteers are recruited. |
| 6 | Loss to follow-up | Loss to follow up of less than 10% AND similar loss in all groups. | More dropouts than expected (10%-30%). Similar loss between groups. | High dropout rate (>30%) and/or significant difference in follow up between groups. |
| 7 | Dietary assessment | Very reliable methodology that does not rely on self-report | Measures are reliable, but self-reported | Unreliable methods including questionnaires that have not been piloted or validated, and methods that increase the likelihood of socially acceptable responses |
| 8 | Behaviour change intervention component | Interventions clearly outlined a behaviour change or psychological theory within the intervention design | Components from the Behaviour Change Wheel can be associated to the intervention. | There was no or limited mention of behaviour change intervention functions |
| 9 | Performance bias | The intervention was delivered in a way that was not variable eg. Computer-generated. | The intervention was probably delivered consistently. Eg. The same person delivering a lesson to multiple groups. | The intervention was likely to have been delivered differently across the trial. Eg. Different health workers promoting nutrition education |
| 10 | Intention to treat | Intention to treat analysis was clearly used |  | Intention to treat was not used, or not stated |
| 11 | Analytic methods | Statistical tests used are rigorous and appropriate for the data set. (Regression modelling, ANOVA, etc) OR rigorous qualitative analysis (thematic analysis) | Statistical methods are effective, but not as rigorous as they could be. | Statistical methods are limited, and only report descriptive stats OR tests used are not appropriate for the dataset. |
| 12 | Did the analysis adjust for confounding?<br>• Educational attainment or SES<br>• BMI/weight<br>• Gender<br>• Age<br>• Baseline behaviour | The statistical model used adjusts for all relevant confounding factors. | The model adjusts for 3 or 4 confounders. | The analysis adjusts for fewer than 3 confounders |

## Appendix E Behaviour change systematic review quality assessment form

|  | Quality Assessment (Author, year, country, journal) |  |  |
| --- | --- | --- | --- |
|  | Criteria | Description | Risk of bias score |
| 1 | Study design |  |  |
| 2 | Randomisation |  |  |
| 3 | Blinding |  |  |
| 4 | Were groups similar at baseline? |  |  |
| 5 | Selection (Were the selection criteria for the study specified?) |  |  |
| 6 | Loss to follow-up |  |  |
| 7 | Dietary assessment |  |  |
| 8 | Behaviour change intervention component |  |  |
| 9 | Performance bias -<br>Was the intervention executed in the same way across all participants/clusters? |  |  |
| 10 | Intention to treat |  |  |
| 11 | Analytical methods |  |  |
| 12 | Did the analysis adjust for confounding? |  |  |
|  | Reviewer's comments |  |  |

